# Structural deviations of the posterior fossa and the cerebellum and their cognitive links in a neurodevelopmental deletion syndrome

**DOI:** 10.1101/2022.03.01.22271659

**Authors:** Esra Sefik, Yiheng Li, Brittney Sholar, Lindsey Evans, Jordan Pincus, Zeena Ammar, Melissa M. Murphy, Cheryl Klaiman, Celine A. Saulnier, Stormi P. White, Adam Ezra Goldman-Yassen, Ying Guo, Elaine F. Walker, Longchuan Li, Sarah Shultz, Jennifer G. Mulle

**Author notes:** Correspondence: Jennifer G. Mulle, MHS, PhD Associate Professor Department of Psychiatry Robert Wood Johnson Medical School Rutgers University /. Indicates joint last authorship (Longchuan Li, Sarah Shultz, Jennifer G. Mulle). <u>Supplemental information:</u> One supplemental materials document that includes 13 supplemental figures, 11 supplemental tables, and extended methods / results.

## Abstract

**Background:** High-impact genetic variants associated with neurodevelopmental disorders provide biologically defined entry points for etiological discovery. The 3q29 deletion (3q29Del) is one such variant that confers a ∼40-fold increased risk for schizophrenia, and a ∼30-fold increased risk for autism. However, the specific neural mechanisms underlying this link remain largely unknown.

**Methods:** Here, we report the first *in vivo* quantitative neuroimaging study in 3q29Del individuals (*N*=24) and healthy controls (*N*=1,608) using structural MRI. Given prior reports of posterior fossa abnormalities in 3q29Del, we focus our investigation on the cerebellum and its primary tissue-types. Additionally, we compare the prevalence of cystic/cyst-like malformations of the posterior fossa between 3q29Del participants and controls, and examine the association between neuroanatomical findings and standardized behavioral measures to probe gene-brain-behavior relationships.

**Results:** 3q29Del participants had smaller cerebellar cortex volumes than controls, both before and after correction for intracranial volume (ICV). 3q29Del participants also had larger cerebellar white matter volumes than controls following ICV-correction. The 3q29Del group displayed an elevated rate of posterior fossa arachnoid cysts and mega cisterna magna findings independent of cerebellar volume. Sex played a moderating role in a subset of findings. Cerebellar white matter volume was positively associated with visual-motor integration skills and cognitive ability, while cystic/cyst-like malformations yielded no behavioral link.

**Conclusions:** Abnormal development of posterior fossa structures may represent neuroimaging-based biomarkers in 3q29Del. Results reveal cerebellar associations with sensorimotor and cognitive deficits in 3q29Del and present a novel point of genetic convergence with cerebellar pathology reported in idiopathic forms of neurodevelopmental disease.

## Introduction

Copy number variation (CNV) of DNA sequences (gain/loss of >1-Kb genomic material) has been shown to represent a significant source of genetic diversity (1–5) and a far more important substrate for human evolution and adaptation than previously recognized (6–9). Accumulating findings indicate the existence of multiple rare CNVs that increase susceptibility to neurodevelopmental disorders (10–22). However, many CNVs remain understudied (23). Given their defined genomic boundaries, systematic investigation of recurring pathogenic CNVs can greatly further our understanding of the biology underlying complex neurodevelopmental disorders, such as schizophrenia (SZ), autism spectrum disorder (ASD), and intellectual disability.

Noninvasive neuroimaging technologies, such as high-resolution structural magnetic resonance imaging (MRI), offer an instrumental tool to elucidate the contribution of specific genetic variants to anatomical changes in the living human brain (24, 25). Such intermediate neuroanatomical phenotypes are crucial for bridging the gap between molecular/cellular mechanisms directly downstream of disease loci and behavioral endpoints (26, 27). The integration of genomics and imaging is especially promising for yielding biological insights into high-impact CNVs, as these variants are expected to produce highly disruptive and relatively consistent deviations in brain structure with greater etiological salience, which can help disentangle the extensive heterogeneity observed across studies of brain structure in idiopathic patient populations (28–32).

The 3q29 deletion (3q29Del) is one of eight recurrent CNVs shown to increase risk for SZ at genome-wide significance (10). 3q29Del confers an estimated >40-fold increased risk for SZ, which is one of the most robust and highest known effect sizes in the genetic landscape of this disorder (10, 33–37). It also confers a >30-fold increased risk for ASD, and exhibits pleiotropy for a range of phenotypes, including intellectual disability, developmental delay, attention deficit hyperactivity disorder (ADHD), and graphomotor deficits (38–41). 3q29Del has a prevalence of approximately 1 in 30,000 and usually arises *de novo* during parental meiosis due to the hemizygous deletion of a 1.6-Mb locus, spanning 21 protein-coding genes (39, 42). Although several genes within the interval have been proposed as putative drivers (43, 44), it is not currently known which genes or biological mechanisms contribute to the emergence of abnormal neurodevelopmental phenotypes in 3q29Del.

In a recent study, our group performed deep-phenotyping of the largest sample of 3q29Del participants with gold-standard instruments and structural MRI (39). Neuroradiological inspection revealed abnormal posterior fossa structures (between the tentorium cerebelli and foramen magnum) in over 70% of participants, including cerebellar hypoplasia and cystic/cyst-like malformations, suggesting that this brain region may be particularly relevant to 3q29Del biology. Other neuroimaging studies of 3q29Del have been based on descriptive single-case studies or small case-series with limited generalizability (41, 45–53). To our knowledge, no case-control analysis nor quantitative investigation of brain morphology has been reported in this syndrome.

Here, we report the first *in vivo* quantitative structural MRI study in individuals with 3q29Del and healthy controls. Motivated by previous findings on posterior fossa abnormalities (39), we focus our investigation on volumetric properties of the cerebellum and its primary tissue-types. Additionally, we expand our previous radiological investigation to the whole brain and test whether the prevalence of cystic/cyst-like malformations of the posterior fossa, which are typically considered incidental, asymptomatic findings in the general population (54–56), differ between 3q29Del and controls. Finally, given accumulating evidence implicating the cerebellum in high-order cognitive functions besides sensorimotor control (57–60), we examine the relationship between neuroanatomical findings and cognitive and sensorimotor deficits in 3q29Del to probe gene-brain-behavior relationships.

The findings reported in the present study have important implications for neuroimaging-based biomarker discovery in 3q29Del, and open novel directions for focusing cellular and molecular studies on target brain regions and circuits. These data also contribute to our knowledge of the genetic loci that subserve the normative development of the posterior fossa, particularly the cerebellum, which has been largely understudied in the context of neurodevelopmental and psychiatric disease.

## Methods and Materials

### Participants

#### The 3q29Del sample

3q29Del participants were ascertained from the 3q29 Registry and traveled to Atlanta, GA for evaluation with standardized tools and structural MRI. Data from 24 individuals with 3q29Del, ages 4-39 years (mean ± SD = 14.71 ± 9.21 years, 62.50% male) were included in this study. One participant was excluded from volumetric analyses due to inability to extract reliable volumetric measures but was retained for radiological examination. 3q29Del status was confirmed via the clinical genetics report and/or medical records. Participants provided informed consent/assent; if the participant was a minor, a parent/legal guardian additionally provided consent. All 3q29Del study procedures were approved by the Emory University Institutional Review Board. The project protocol and a summary of findings in the broader 3q29Del study sample, from which the current sample was drawn, have been detailed elsewhere (39, 61).

#### Healthy controls

To achieve reliable estimates for the structural variability observed in typically developing human brains, we used MRI data from the largest available, open-access sample of healthy controls (*N* = 1,765) from the Human Connectome Project (HCP), with cross-sectional continuity over a wide age range (5-37 years) overlapping with the 3q29Del sample (4-39 years). This dataset was selected to establish normative benchmarks around our study metrics with high statistical power, while accounting for linear and non-linear growth trajectories and sex differences. Data access was obtained in accordance with the terms outlined by the HCP. Upon inspection of the family relationships reported among controls, one sibling from each monozygotic twin-pair was randomly removed to minimize bias in standard error estimates (62, 63). Hence, 1,608 healthy controls (mean ± SD = 22.73 ± 8.21 years, 46.46% male) were included in the present study. See (64, 65) for an overview of HCP initiatives and eligibility criteria.

There were no significant differences in the sex, ethnicity, or race compositions of the two diagnostic groups (*p’s* > 0.05). While there was a near complete overlap between their age ranges, the 3q29Del group was relatively younger than controls (*p* ≤ 0.001). Hence, we considered age as a potential confound in downstream analyses. Demographic characteristics are presented in Table 1, Fig. 1, and Table S1.

**Figure 1.**
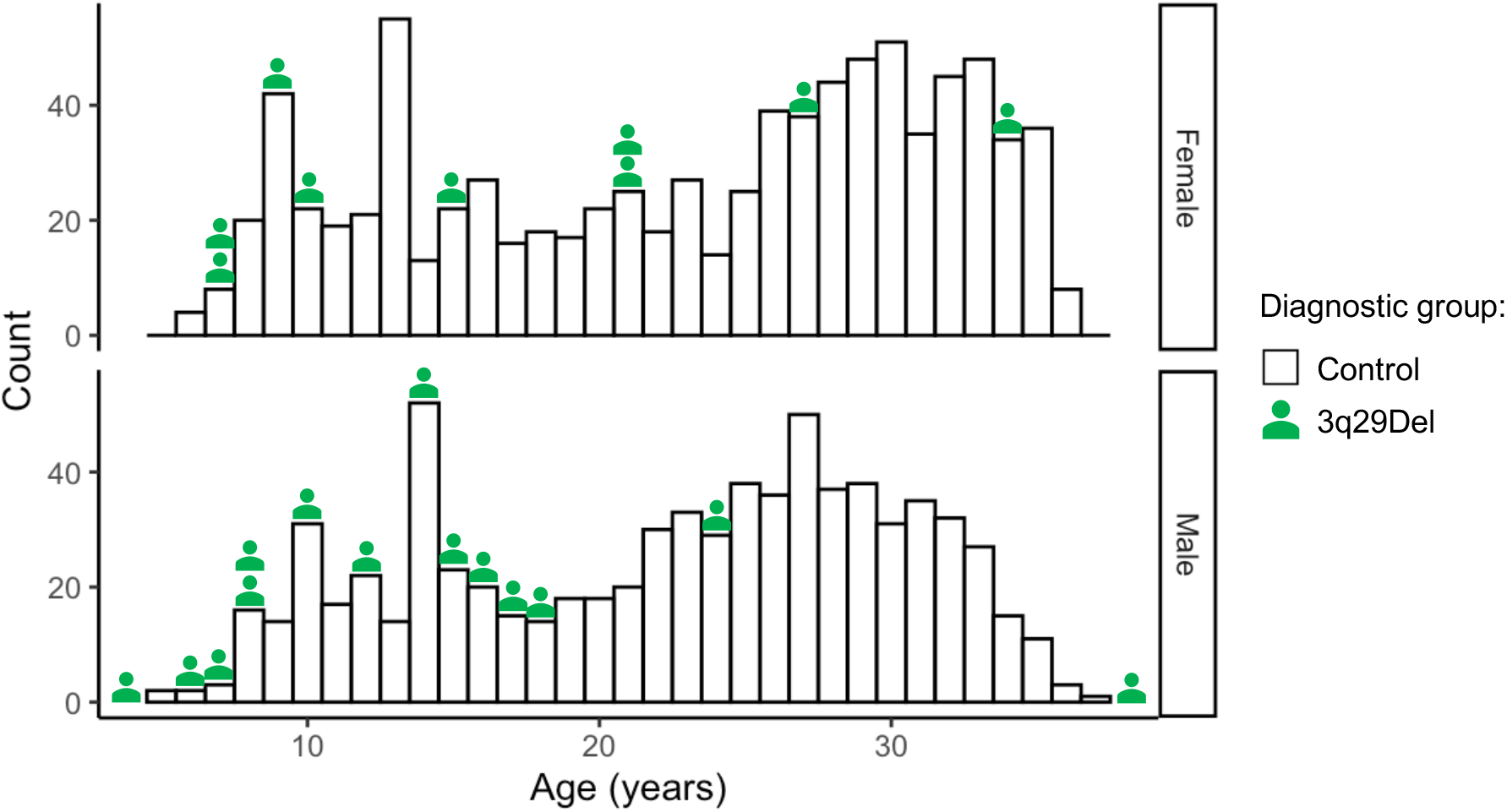
Histogram showing the age distribution of study participants in volumetric analyses, stratified by sex and diagnostic group. To define reliable estimates for the normative trajectory of cerebellar volumetric change across age, we used the largest available open-access sample of healthy controls from the Human Connectome Project, which cross-sectionally covers a wide age range [5 – 37 years] with near complete overlap with the 3q29Del sample [4 – 39 years]. Each histogram bin represents one year. Green icons above each bar symbolize the number of 3q29Del participants included in volumetric analyses for a given age and sex. Control *N* = 1,608 (Female *N* = 861, Male *N* = 747), 3q29Del *N* = 23 (Female *N* = 9, Male *N* = 14). *Abbreviations*: 3q29 deletion syndrome, 3q29Del.

**Table 1.** Demographic characteristics of the study sample in volumetric analyses, stratified by diagnostic group. There were no significant differences in the sex, ethnicity, or race compositions of the two diagnostic groups (p’s > 0.05). While there was a near complete overlap between the age ranges of the two groups, there was a significant age difference between 3q29Del participants and controls on average (p ≤ 0.001). Effect sizes are reported for significant test results only. Non-parametric statistics are reported in cases where the data do not meet parametric assumptions. ^#^Control N = 1,587 for the ethnicity variable due to missing data. ^##^Control N = 1,574 for the race variable due to missing data. Corresponding percentages reflect the fraction of controls with complete data. ^a^ Wilcoxon rank sum test with continuity correction, ^b^ Pearson’s chi-squared test with Yates’ continuity correction. p-value ≤ 0.001 ‘***’, p-value ≤ 0.01 ‘**’, p-value ≤ 0.05 ‘*’, p-value ≤ 0.1 ‘^†^’ Abbreviations: 3q29 deletion syndrome, 3q29Del; standard deviation, SD; degrees of freedom, DF.

| Demographic variables | Control<br>N = 1,608 | 3q29Del<br>N = 23 | Test statistics |
| --- | --- | --- | --- |
| **Age** (in years) |  |  |  |
| Mean ± SD | 22.73 ± 8.21 | 15.09 ± 9.22 | $r = 0.10$ (small effect size),<br>$W = 9429.5$,<br>p-value<sup>a</sup> = 5.26E-05*** |
| Median | 25.00 | 14.00 |  |
| [Range] | [5 – 37] | [4 – 39] |  |
| **Sex**, N (%) |  |  |  |
| Male | 747 (46.46%) | 14 (60.87%) | $\chi^2 = 1.36$, DF = 1,<br>p-value<sup>b</sup> = 0.24 |
| Female | 861 (53.54%) | 9 (39.13%) |  |
| **Ethnicity**, N (%) |  |  |  |
| Non-Hispanic / Latino | 1,397 (88.03%) | 22 (95.65%) | $\chi^2 = 0.64$, DF = 1,<br>p-value<sup>b</sup> = 0.42 |
| Hispanic / Latino | 190 (11.97%) | 1 (4.35%) |  |
| **Race**, N (%) |  |  |  |
| White | 1,112 (70.66%) | 21 (91.30%) | $\chi^2 = 6.25$, DF = 4,<br>p-value<sup>b</sup> = 0.18 |
| Black / African American | 222 (14.10%) | 0 (0%) |  |
| Asian / Native Hawaiian /<br>Other Pacific Islander | 108 (6.86%) | 0 (0%) |  |
| American Indian / Alaskan Native | 4 (0.25%) | 0 (0%) |  |
| More than one race | 128 (8.13%) | 2 (8.70%) |  |

### Structural MRI acquisition and processing

High-resolution structural MRI data were collected from 3q29Del participants on a 3T Siemens Magnetom Prisma scanner, using a Siemens 32-channel head coil and an 80mT/m gradient. T1-weighted 3D images were acquired in the sagittal plane using a single-echo MPRAGE sequence (66) with the following parameters: TE=2.24ms, TR=2400ms, TI=1000ms, bandwidth=210Hz/pixel, FOV=256x256mm, resolution=0.8mm isotropic. T2-weighted 3D images were acquired in the sagittal plane using a SPACE sequence (67) with the following parameters: TE=563ms, TR=3200ms, bandwidth=745Hz/pixel, FOV=256x256mm, resolution=0.8mm isotropic.

Control participants were imaged using a 3T Siemens Prisma or Skyra “Connectom” scanner by the HCP Development and Young Adult consortiums, using a Siemens 32-channel head coil with 80 or 100 mT/m gradients. T1- and T2-weighted 3D images were acquired in the sagittal plane using single- or multi-echo MPRAGE and SPACE sequences, respectively (65, 68, 69). Comparison of the 3q29Del and control protocols indicate that fundamental aspects of the hardware and acquisition parameters are either identical or highly comparable between study samples for combined hypothesis testing (see Table S2 for technical details).

For volumetrics, all 3q29Del scans were processed with the HCP “minimal pre-processing” pipeline to remove spatial artifacts/distortions, align images to MNI space and obtain brain masks/parcellations (70). Image processing for controls was performed by the HCP using this common framework. FreeSurfer software (https://surfer.nmr.mgh.harvard.edu) was used for automated segmentations, based on probabilistic information estimated from a manually labeled training set (71). Using this approach, we extracted volumetric measures for cerebellar cortex and white matter to distinguish the two primary tissue-types of the cerebellum; total cerebellar volume was defined as the sum of these metrics. See Supplemental Materials, Fig. S1 and Fig. S2 for extended methods.

### Normalization of cerebellar volumes

Since skull dimensions and vertebrate brain size are correlated (72–74), correction for inter-subject variation in head size is recommended in analyses of brain volume (75, 76). We used the estimated total intracranial volume (eICV) generated by FreeSurfer as a proxy for head size (77). This method uses an automated atlas-based scaling factor estimated during affine transformation of images from native to standard space, and concords well with manually delineated ICV (78).

An important consideration in deciding whether and how regional brain volumes should be adjusted for head size, is the potential relevance of this variable to intrinsic disease processes (79, 80). Given prior reports of microcephaly in 3q29Del (41), this question is particularly germane to the present study. Hence, we evaluated the relationship between eICV and cerebellar volumes and tested for potential interaction effects between eICV and diagnostic group. Most cerebellar volumes did not scale directly proportionally with eICV, and the relationship between cerebellar white matter volume and eICV changed as a function of diagnostic group (*p* = 0.03) (Fig. S3). Consequently, all volumes were adjusted for eICV using the “residual” method (Fig. S4), based on normative regression slopes derived from the control group, as described in (75, 79, 81). The distributions of absolute and eICV-adjusted volumetric measures of interest (VOI) are visualized in Fig. 2 and Fig. S5.

**Figure 2.**
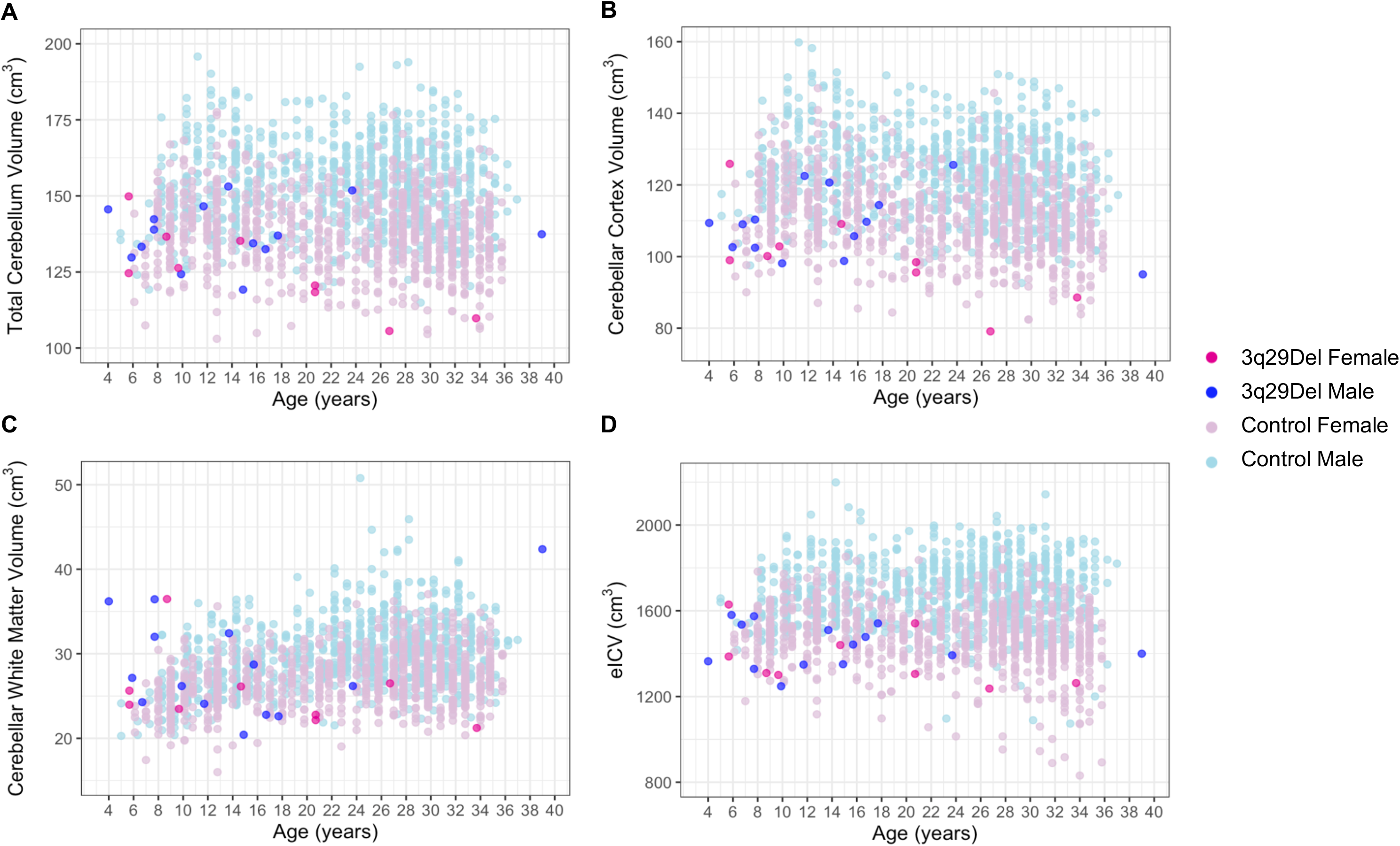
Scatter plots showing the distribution of A) total cerebellum volume, B) cerebellar cortex volume, C) cerebellar white matter volume, and D) eICV as a function of age among male and female participants in each diagnostic group. A slight jitter was added systematically to all panels to minimize overplotting. Data reflect absolute volumes. Control *N* = 1,608 (Female *N* = 861, Male *N* = 747), 3q29Del *N* = 23 (Female *N* = 9, Male *N* = 14). *Abbreviations*: 3q29 deletion syndrome, 3q29Del; estimated total intracranial volume, eICV.

### Radiological evaluation of MRI scans

Prior work by our group using the same 3q29Del scans as in the present study has shown that posterior fossa arachnoid cysts (PFAC) are common among 3q29Del carriers (39). However, our original report did not explicitly evaluate regions outside the posterior fossa. To probe for patterns of relative regional specificity, all 3q29Del scans (*N* = 24) were reviewed qualitatively by a board-certified neuroradiologist (AEGY) at the whole-brain level. T1- and T2-weighted images were reviewed in axial, sagittal, and coronal reformats using Horos (https://horosproject.org). Given well-established challenges in differentiating PFAC from mega cisterna magna (MCM) (shared characteristics in appearance) (82, 83), their prevalence rates were considered jointly.

We also pulled radiographic data on incidental findings in healthy controls (*N* = 1,608) to define normative estimates for the prevalence of cystic/cyst-like malformations of the posterior fossa. Anatomical anomalies were identified by HCP raters and radiologists during standard quality control. Prior to case-control comparisons, the neuroradiologist who evaluated the 3q29Del scans independently confirmed the control findings for consistency. See Supplemental Materials for details.

### Standardized behavioral measures

3q29Del participants also participated in a standardized, norm-referenced battery of cognitive and sensorimotor tests, in the context of a broader deep-phenotyping study described elsewhere by our group (39, 61). Participants were administered the *Beery-Buktenica Developmental Test of Visual-Motor Integration* (VMI, 6^th^ edition) to assess the extent to which they can integrate their visual and fine motor abilities in geometric design-copying tasks (84). Supplemental *Beery-VMI* tests were administered to measure visual perception and fine motor coordination, separately. Cognitive ability (composite, verbal, non-verbal IQ) was assessed using the *Differential Ability Scales* (DAS, 2^nd^ edition) among participants aged 4-17 years (85), and the *Wechsler Abbreviated Scale of Intelligence* (WASI, 2^nd^ edition) among participants aged 18-39 years (86).

### Statistical analyses

#### Case-control comparison of volumetric measures

All statistical analyses were performed using *R* version 4.0.3 (87). Demographic characteristics of the diagnostic groups were compared using Wilcoxon rank sum test and Pearson’s chi-squared test. We used multiple linear regression to model the main effect of diagnostic group on each VOI separately, while correcting for sex and age. Both absolute and eICV-adjusted cerebellar volumes, as well as eICV itself, were tested for group differences, as eICV variation may reflect a biological signal relevant to 3q29Del beyond nuisance variation. Analyses of variance (ANOVA) were performed to identify the best-fitting polynomial function of age for each VOI (see Table S3 for details). Multiple comparisons correction was applied at the VOI level (7 tests) to control the false discovery rate (FDR) using the Benjamini-Hochberg procedure. Since reliance on asymptotic theory can be problematic in small-moderate sample sizes, we additionally calculated exact p-values for model coefficients using marginal permutation tests (10,000 random permutations).

To evaluate whether the effects of diagnostic group on individual VOIs change as a function of sex, we also performed an exploratory analysis of diagnostic group and sex interactions, while correcting for age. In cases where a significant product term was identified (*p* ≤ 0.05), males and females were subsequently tested separately. Given the exploratory nature of these analyses, FDR correction was not applied. Finally, in supplemental analyses, we adopted non-parametric spline modeling to build developmental trajectories for each VOI with increased flexibility and to estimate normative percentile curves for our VOIs akin to growth curves. Since splines can capture a wide range of nonlinear trends, penalized cubic spline and quantile spline methods were incorporated to improve the statistical rigor of our central analyses. See Supplemental Materials for details.

#### Case-control comparison of radiological findings

The prevalence of PFAC/MCM findings was compared between 3q29Del and control groups using Fisher’s exact test. Sex-specific differences in these rates were evaluated within each group using the same procedure. The demographic characteristics of 3q29Del participants with versus without cystic/cyst-like malformations were compared using Student’s two sample t-test and Fisher’s exact test. To rule out conceivable secondary (acquired) etiologies (88–90), differences in the distribution of past head injuries, maternal complications during pregnancy and neonatal complications during delivery were tested between these 3q29Del groups using Fisher’s exact test. Finally, to determine whether the likelihood of these cystic/cyst-like malformations covary with volumetric measures among 3q29Del participants, we modeled the relationship between PFAC/MCM findings and our VOIs using linear regression, with sex, age, and eICV considered as covariates.

#### Tests of brain-behavior relationships in 3q29Del

To probe the functional relevance of our tissue-specific MRI findings, we investigated the association of cerebellar cortex and white matter volumes with composite IQ and VMI scores among 3q29Del participants in separate linear regression models, with adjustment for age and sex. Multiple comparisons correction was applied at the VOI level (4 tests) using Benjamini-Hochberg. Secondary analyses were conducted to investigate which cognitive (verbal / non-verbal IQ) and sensorimotor (visual perception / fine motor coordination) subprocesses may be associated with cerebellar volumes, using the same procedure. Given the exploratory nature of these analyses, FDR correction was not applied. In models where a significant relationship was identified (*p* ≤ 0.05), we subsequently added eICV as an additional covariate to test whether results reflect a link with the cerebellum beyond global variability in head size. To examine the functional relevance of our radiological findings, we compared the standardized test scores of 3q29Del participants with versus without cystic/cyst-like malformations of the posterior fossa using Student’s two sample t-test and Wilcoxon rank sum test.

Standard diagnostics were performed to check all required assumptions. If parametric assumptions were not met, non-parametric alternatives were used. In cases where violations were observed for ordinary least squares regression (see Fig. S6), heteroscedasticity-robust estimates were calculated using the HC1 robust standard error estimator (91). All analyses were two-tailed. See Supplemental Materials for extended methods.

## Results

### Case-control differences in volumetric measures

Using multiple linear regression, we first tested for diagnostic group differences in our VOIs, while correcting for age and sex (Table 2, Table S3A-H, Fig. 3A-G). Participants with 3q29Del had significantly smaller eICVs than controls (*b* = -197.99, *p* ≤ 0.001, *FDR adjusted-p* ≤ 0.001); hence, we report case-control comparisons for both absolute and eICV-adjusted volumes while examining regional changes.

**Figure 3.**
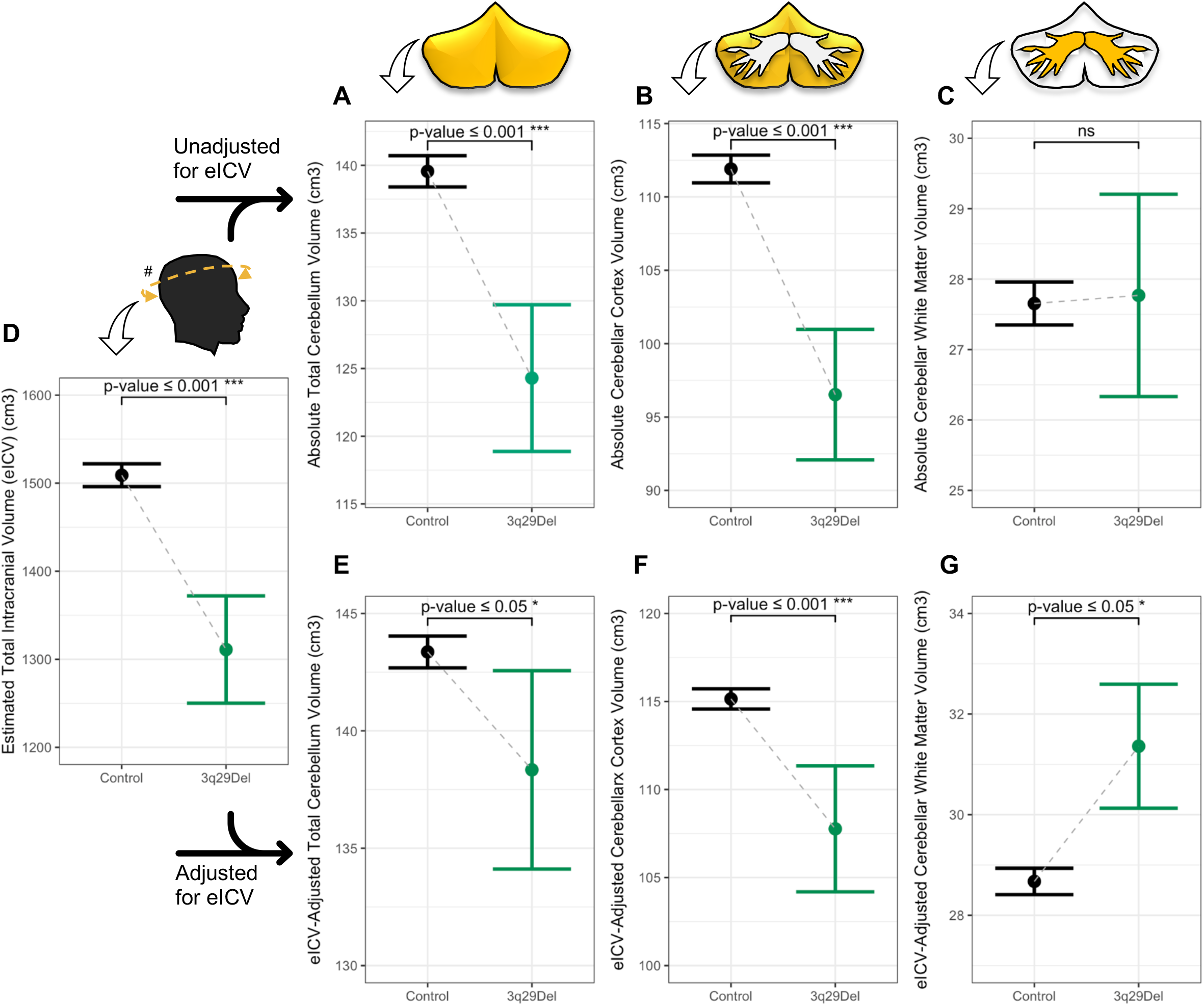
Predictor effect plots showing the effect of diagnostic group on volumetric measures of interest. **A-G)** Predicted values of VOIs across the 3q29Del and control groups were computed from the multiple linear regression models reported in Table 2, while covariates (sex, age, age^2^ (when applicable)) were held fixed. Error bars indicate the 95% confidence interval. P-values for the main effect of diagnostic group are indicated on each plot and reflect heteroskedasticity-robust estimates. A schematic of each VOI (yellow) is presented above the corresponding plot for clarity. Regression results indicate a significant difference between 3q29Del participants and healthy controls in **A)** total cerebellum volume (*p* ≤ 0.001), **B)** cerebellar cortex volume (*p* ≤ 0.001), and **D)** eICV (*p* ≤ 0.001), with smaller volumes observed in 3q29Del participants compared with controls. This effect remained significant after **E)** total cerebellum volume (*p* ≤ 0.05) and **F)** cerebellar cortex volume (*p* ≤ 0.001) were adjusted for eICV. There was no significant effect of diagnostic group on **C)** absolute cerebellar white matter volume (*p* > 0.05), however **G)** after eICV-adjustment, 3q29Del participants had significantly larger cerebellar white matter volumes than controls (*p* ≤ 0.05). Control *N* = 1,608, 3q29Del *N* = 23. ^#^eICV was calculated by FreeSurfer’s atlas-based spatial normalization procedure. p-value ≤ 0.001 ‘***’, p-value ≤ 0.01 ‘**’, p-value ≤ 0.05 ‘*’, p-value ≤ 0.1 *‘^†^’ Abbreviations*: 3q29 deletion syndrome, 3q29Del; volumetric measure of interest, VOI; estimated total intracranial volume, eICV; not significant, ns.

**Table 2.** Summary of multiple linear regression results testing for differences in volumetric measures of interest between 3q29Del and control participants. Sex, age, and age^2^ (when applicable) were included as covariates in each regression equation based on the best-fitting polynomial function of age identified for each VOI (see Table S3 for details). The main effect of diagnostic group is reported in bold for clarity. Regression parameters reflect heteroskedasticity-robust estimates, which were computed as an alternative to ordinary least squares regression to address observed violations of statistical assumptions. Robust Wald test statistics are reported to assess the overall significance of each model. Control N = 1,608, 3q29Del N = 23. Contrast coding: reference levels for the categorical diagnostic group and sex variables are healthy control and female, respectively. p-value ≤ 0.001 ‘***’, p-value ≤ 0.01 ‘**’, p-value ≤ 0.05 ‘*’, p-value ≤ 0.1 ‘^†^’. Abbreviations: 3q29 deletion syndrome, 3q29Del; VOI, volumetric measure of interest; eICV, estimated total intracranial volume; unstandardized coefficient estimate, b; confidence interval, CI; degrees of freedom, DF.

| Outcome variables | Explanatory variables | *b* | CI (95%) | p-value |
| --- | --- | --- | --- | --- |
| **Absolute Total Cerebellum Volume (cm<sup>3</sup>)** | Intercept | 133.50 | 128.97 – 138.03 | < 2.00E-16*** |
|  | Age (years) | 0.68 | 0.22 – 1.14 | 3.72E-03** |
|  | Age<sup>2</sup> | -0.02 | -0.03 – -0.01 | 6.22E-04*** |
|  | Sex [Male] | 16.22 | 14.97 – 17.48 | < 2.00E-16*** |
|  | **Diagnostic Group [3q29Del]** | **-15.26** | **-20.01 – -10.51** | **3.76E-10\*\*\*** |
|  | R<sup>2</sup> / R<sup>2</sup> adjusted<br>Robust Wald test | 0.31 / 0.31<br>184.0 on 4 and 1626 DF, p-value < 2.20E-16*** |  |  |
| **Absolute Cerebellar Cortex Volume (cm<sup>3</sup>)** | Intercept | 113.52 | 109.73 – 117.31 | < 2.00E-16*** |
|  | Age (years) | 0.13 | -0.26 – 0.51 | 0.52 |
|  | Age<sup>2</sup> | -0.01 | -0.02 – -0.0001 | 0.04* |
|  | Sex [Male] | 13.13 | 12.10 – 14.17 | < 2.00E-16*** |
|  | **Diagnostic Group [3q29Del]** | **-15.38** | **-19.53 – -11.22** | **5.86E-13\*\*\*** |
|  | R<sup>2</sup> / R<sup>2</sup> adjusted<br>Robust Wald test | 0.32 / 0.32<br>193.2 on 4 and 1626 DF, p-value < 2.20E-16*** |  |  |
| **Absolute Cerebellar White Matter Volume (cm<sup>3</sup>)** | Intercept | 19.98 | 18.82 – 21.15 | < 2.00E-16*** |
|  | Age (years) | 0.55 | 0.43 – 0.67 | < 2.00E-16*** |
|  | Age<sup>2</sup> | -0.01 | -0.01 – -0.01 | 8.88E-11*** |
|  | Sex [Male] | 3.09 | 2.76 – 3.43 | < 2.00E-16*** |
|  | **Diagnostic Group [3q29Del]** | **0.11** | **-2.42 – 2.65** | **0.93** |
|  | R<sup>2</sup> / R<sup>2</sup> adjusted<br>Robust Wald test | 0.28 / 0.27<br>155.1 on 4 and 1626 DF, p-value < 2.20E-16*** |  |  |
| **Estimated Total Intracranial Volume (eICV) (cm<sup>3</sup>)** | Intercept | 1415.02 | 1367.87 – 1462.17 | < 2.00E-16*** |
|  | Age (years) | 9.89 | 4.84 – 14.94 | 1.26E-04*** |
|  | Age<sup>2</sup> | -0.25 | -0.37 – -0.13 | 3.66E-05*** |
|  | Sex [Male] | 206.00 | 192.06 – 219.93 | < 2.00E-16*** |
|  | **Diagnostic Group [3q29Del]** | **-197.99** | **-253.23 – -142.74** | **3.05E-12\*\*\*** |
|  | R<sup>2</sup> / R<sup>2</sup> adjusted<br>Robust Wald test | 0.37 / 0.37<br>232.5 on 4 and 1626 DF, p-value < 2.20E-16*** |  |  |
| **eICV-Adjusted Total Cerebellum Volume (cm<sup>3</sup>)** | Intercept | 144.44 | 142.97 – 145.92 | < 2.00E-16*** |
|  | Age (years) | -0.05 | -0.11 – 0.01 | 0.12 |
|  | Sex [Male] | 5.38 | 4.38 – 6.38 | < 2.00E-16*** |
|  | **Diagnostic Group [3q29Del]** | **-5.02** | **-9.25 – -0.80** | **0.02\*** |
|  | R<sup>2</sup> / R<sup>2</sup> adjusted<br>Robust Wald test | 0.07 / 0.07<br>39.0 on 3 and 1627 DF, p-value < 2.20E-16*** |  |  |
| **eICV-Adjusted Cerebellar Cortex Volume (cm<sup>3</sup>)** | Intercept | 119.88 | 118.60 – 121.17 | < 2.00E-16*** |
|  | Age (years) | -0.21 | -0.26 – -0.16 | 1.37E-15*** |
|  | Sex [Male] | 4.87 | 4.03 – 5.72 | < 2.00E-16*** |
|  | **Diagnostic Group [3q29Del]** | **-7.38** | **-10.98 – -3.78** | **6.03E-05\*\*\*** |
|  | R<sup>2</sup> / R<sup>2</sup> adjusted<br>Robust Wald test | 0.11 / 0.11<br>67.5 on 3 and 1627 DF, p-value < 2.20E-16*** |  |  |
| **eICV-Adjusted Cerebellar White Matter Volume (cm<sup>3</sup>)** | Intercept | 22.22 | 21.22 – 23.23 | < 2.00E-16*** |
|  | Age (years) | 0.42 | 0.31 – 0.53 | 6.39E-14*** |
|  | Age<sup>2</sup> | -0.01 | -0.01 – -0.004 | 3.94E-06*** |
|  | Sex [Male] | 0.41 | 0.13 – 0.70 | 4.51E-03** |
|  | **Diagnostic Group [3q29Del]** | **2.69** | **0.10 – 5.28** | **0.04\*** |
|  | R<sup>2</sup> / R<sup>2</sup> adjusted<br>Robust Wald test | 0.19 / 0.19<br>106.7 on 4 and 1626 DF, p-value < 2.20E-16*** |  |  |

The 3q29Del group had significantly smaller total cerebellum volumes than controls (*b* = -15.26, *p* ≤ 0.001, *FDR adjusted-p* ≤ 0.001), and this finding persisted after eICV-adjustment (*b* = -5.02, *p* = 0.02, *FDR adjusted-p* = 0.03). When the cerebellum was broken down to its two primary tissue-types, cerebellar cortex volumes were significantly smaller in the 3q29Del group (*b* = -15.38, *p* ≤ 0.001, *FDR adjusted-p* ≤ 0.001), while cerebellar white matter volumes did not differ between the groups (*p* = 0.93). Case-control differences in cerebellar cortex remained significant after eICV-adjustment (*b* = -7.38, *p* ≤ 0.001, *FDR adjusted-p* ≤ 0.001). Unexpectedly, 3q29Del participants also had significantly larger cerebellar white matter volumes than controls after eICV-adjustment (*b* = 2.69, *p* = 0.04, *FDR adjusted-p* = 0.05).

Considering these findings, we tested participants’ cerebellar cortex to white matter volume ratios for group differences (Table S3D); this revealed significantly smaller ratios in 3q29Del (*b* = -0.49, *p* ≤ 0.01). As an ancillary method, we fit generalized additive models (GAM) with a cubic spline basis to our data to identify group differences in VOIs with greater flexibly for modeling age. GAM results support the case-control differences identified using linear regression (Table S4, Fig. S7).

In exploratory analyses, we evaluated whether the effects of diagnostic group change as a function of sex, while correcting for age (Table S5A-H). There was a significant interaction between diagnostic group and sex on eICV (*b* = -149.64, *p* ≤ 0.01). Both male and female 3q29Del participants had smaller eICVs than controls, however this reduction was greater among male (*b* = -237.09, *p* ≤ 0.001) than female 3q29Del participants (*b* = -119.89, *p* ≤ 0.01) (Fig. 4, Table 3). Both OLS-based asymptotic p-values and exact-p-values by permutation testing were concordant with heteroscedasticity-robust estimates across these analyses.

**Figure 4.**
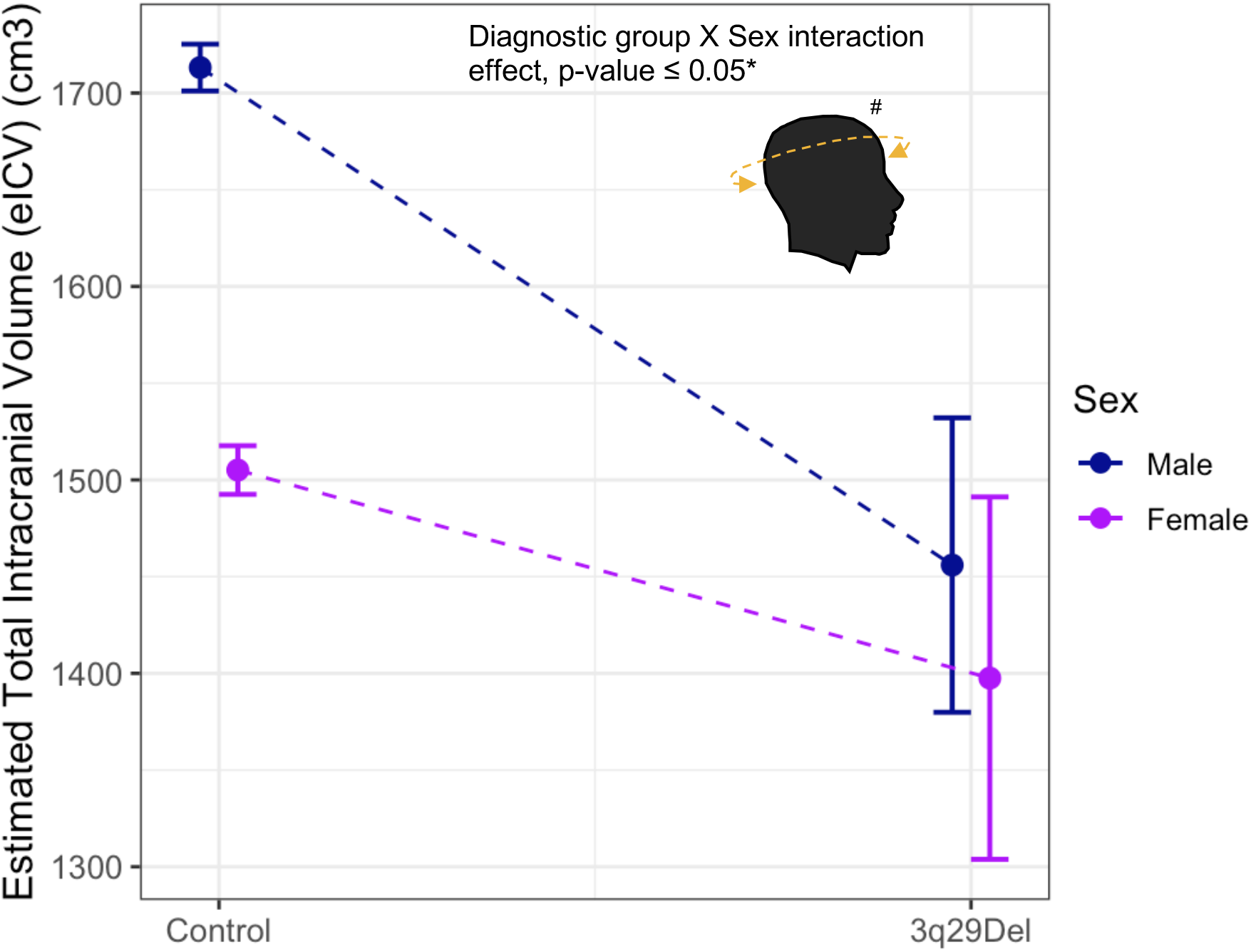
Predictor effect plot showing the moderating effect of sex on the relationship between diagnostic group and eICV. Predicted values of eICV across male versus female 3q29Del and control groups were computed from the exploratory interaction model reported in Table S4E, while covariates (age, age^2^) were held fixed. Error bars indicate the 95% confidence interval. Heteroskedasticity-robust regression results indicate a significant diagnostic group by sex interaction effect on eICV (*p* ≤ 0.05). Control *N* = 1,608 (Female *N* = 861, Male *N* = 747), 3q29Del *N* = 23 (Female *N* = 9, Male *N* = 14). ^#^ eICV was calculated by FreeSurfer’s atlas-based spatial normalization procedure. p-value ≤ 0.001 ‘***’, p-value ≤ 0.01 ‘**’, p-value ≤ 0.05 ‘*’, p-value ≤ 0.1 *‘^†^’ Abbreviations*: 3q29 deletion syndrome, 3q29Del; estimated total intracranial volume, eICV.

**Table 3.**
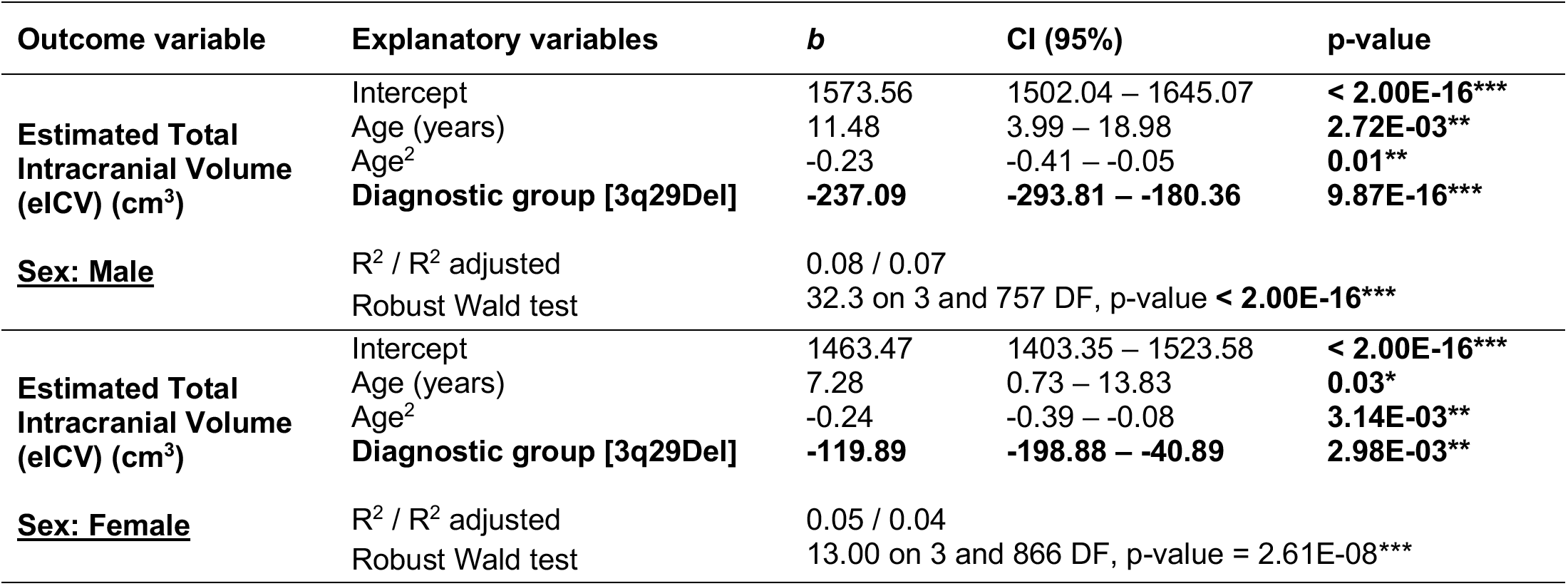
*Post hoc* analysis of the sex-specific effects of diagnostic group on eICV. Sex-stratified linear regression models include age and age^2^ as covariates based on the best-fitting model reported for this VOI in Table 2. The main effect of diagnostic group is reported in bold for clarity. Heteroskedasticity-robust regression results indicated that both male and female 3q29Del participants had smaller eICVs than controls (*p’s* ≤ 0.01), however this reduction was greater among male 3q29Del participants than female 3q29Del participants. Robust Wald test statistics are reported to assess the overall significance of each model. Contrast coding: reference level for the diagnostic group variable is healthy control. Male *N =* 761 (Control *N* = 747, 3q29Del *N* = 14), Female *N =* 870 (Control *N* = 861, 3q29Del *N* = 9). p-value ≤ 0.001 ‘***’, p-value ≤ 0.01 ‘**’, p-value ≤ 0.05 ‘*’, p-value ≤ 0.1 ‘^†^’. *Abbreviations*: 3q29 deletion syndrome, 3q29Del; estimated total intracranial volume, eICV; unstandardized coefficient estimate, *b*; confidence interval, CI; degrees of freedom, DF.

Additionally, OLS regression and permutation testing indicated a significant interaction between diagnostic group and sex on eICV-adjusted cerebellar white matter volumes (*b* = 2.73, *p* = 0.03); however, this effect failed to reach significance when heteroscedasticity-robust estimates were calculated (*p* = 0.29) (see Fig. S8 and Table S6 for sex-stratified results). Finally, using quantile splines as a supplemental method, we estimated sex-specific normative percentile curves for our VOIs. Fig. S9 visualizes the cerebellar volumes and eICV of each 3q29Del participant relative to developmental trajectories in controls, showing that many 3q29Del participants fall into extreme percentiles of growth, although within-group variability was observed in percentile ranks. A summary of case-control differences and descriptive statistics for each VOI are presented in Table S7.

### Case-control differences in radiological findings

Upon radiological evaluation, 11 controls (0.68%, out of 1,608) and 13 3q29Del participants (54.17%, out of 24) were found to have a PFAC/MCM finding. Comparison of these rates indicated a significantly higher prevalence in 3q29Del than controls (OR = 165.87, *p* ≤ 0.001) (Fig. 5A-C). In sex-stratified analyses, among controls, 1 female (0.12%, out of 861) and 10 males (1.34%, out of 747) had a PFAC/MCM, while 5 females (55.56%, out of 9) and 8 males (53.33%, out of 15) had a PFAC/MCM in 3q29Del (Fig. 5D). Comparison of these sex-specific rates indicated an increase among male compared with female controls (OR = 11.66, *p* ≤ 0.01), but no significant difference between male and female 3q29Del participants (OR = 0.92, *p* > 0.05).

**Figure 5.**
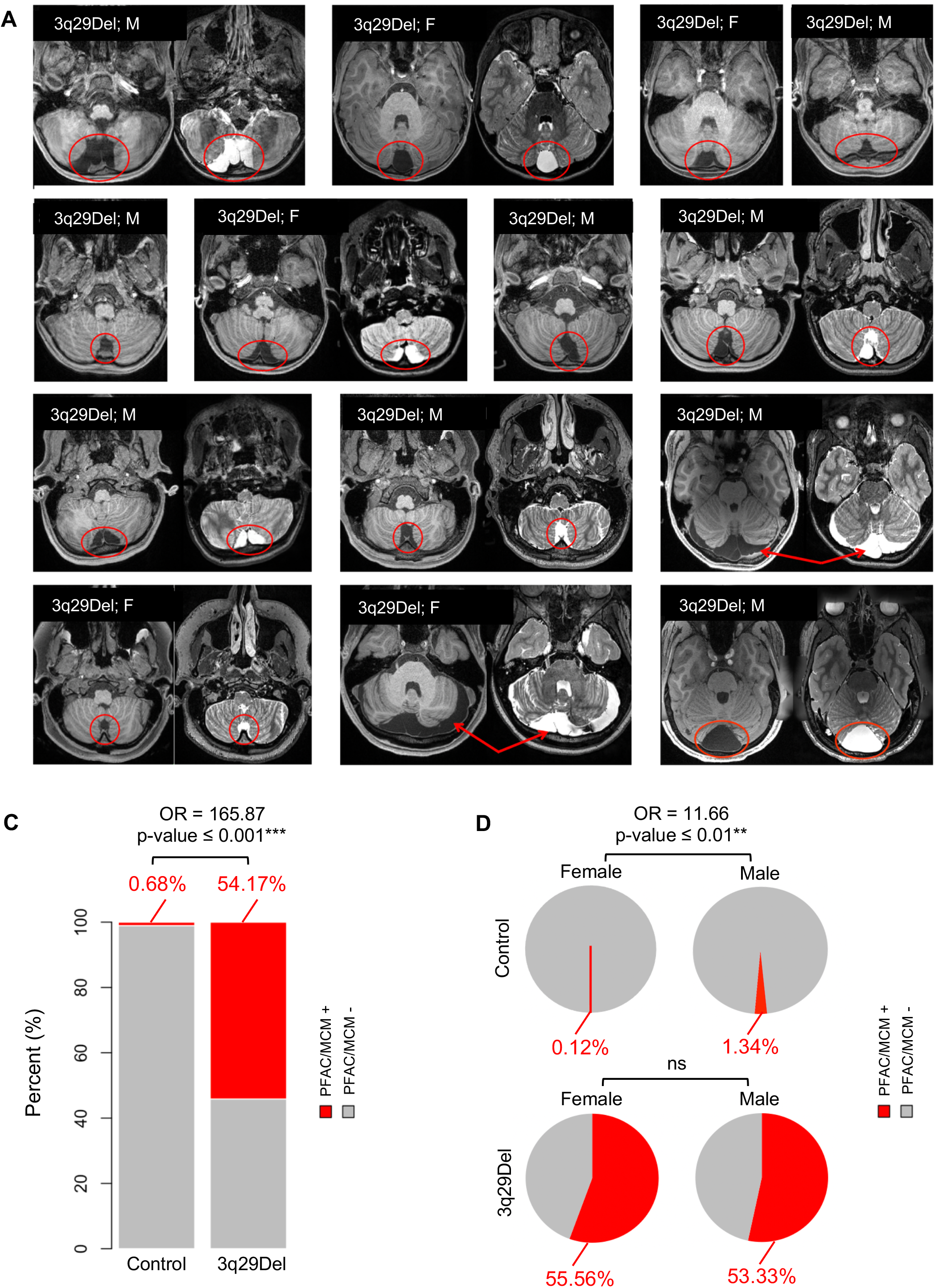
Prevalence of posterior fossa arachnoid cyst and mega cisterna magna findings in structural MRI scans of 3q29Del and control participants. A) 13 3q29Del participants had a PFAC/MCM finding (marked in red) upon radiological evaluation of structural MRI scans. Representative T1- and/or T2-weighted MR images showing these radiological findings are provided in axial view for each 3q29Del participant. B) T1- and T2-weighted MR images of a representative control participant with a PFAC/MCM finding (marked in red). Sex information for each participant is indicated on individual images. C) Bar graphs represent the frequency of PFAC/MCM findings in the 3q29Del and control groups, separately. 3q29Del participants had a significantly elevated rate of PFAC/MCM findings compared with controls (*p* ≤ 0.001) D) Pie charts represent the sex-stratified frequency of PFAC/MCM findings in the 3q29Del and control groups, separately. Male controls had a significantly elevated rate of PFAC/MCM findings compared with female controls (*p* ≤ 0.01), while there were no sex differences in these rates within the 3q29Del group (*p* > 0.05). Control *N* = 1,608 (Female *N =* 861, Male *N =* 747), 3q29Del *N* = 24 (Female *N* = 9, Male *N* = 15). p-value ≤ 0.001 ‘***’, p-value ≤ 0.01 ‘**’, p-value ≤ 0.05 ‘*’, p-value ≤ 0.1 ‘^†^’. *Abbreviations*: 3q29 deletion syndrome, 3q29Del; posterior fossa arachnoid cyst, PFAC; mega cisterna magna, MCM; magnetic resonance imaging, MRI; years-old, y/o; male, M; female, F; odds ratio, OR; not significant, ns.

Furthermore, there were no significant differences between 3q29Del participants with versus without PFAC/MCM findings in age, ethnicity, or race, nor in the distribution of past head injuries, maternal complications during pregnancy and/or neonatal complications during delivery (*p’s* > 0.05) (Table S8), suggesting that these findings are most likely of primary (congenital) origin and cannot be readily explained by conceivable secondary causes. Notably, in our whole-brain examination, only one 3q29Del participant (4.17%, out of 24) had an arachnoid cyst outside the posterior fossa (parietal lobe), highlighting increased vulnerability of the posterior fossa in this syndrome.

Lastly, regression results indicated no significant association between the likelihood of PFAC/MCM and volumetric variations in the cerebellum or eICV among 3q29Del participants, while controlling for age, sex, and/or eICV (*p’s* > 0.05) (Table 4, Fig. S10), implying that VOI changes identified in the present study are not simply a byproduct of the cystic/cyst-like malformations enriched in this region.

**Table 4.**
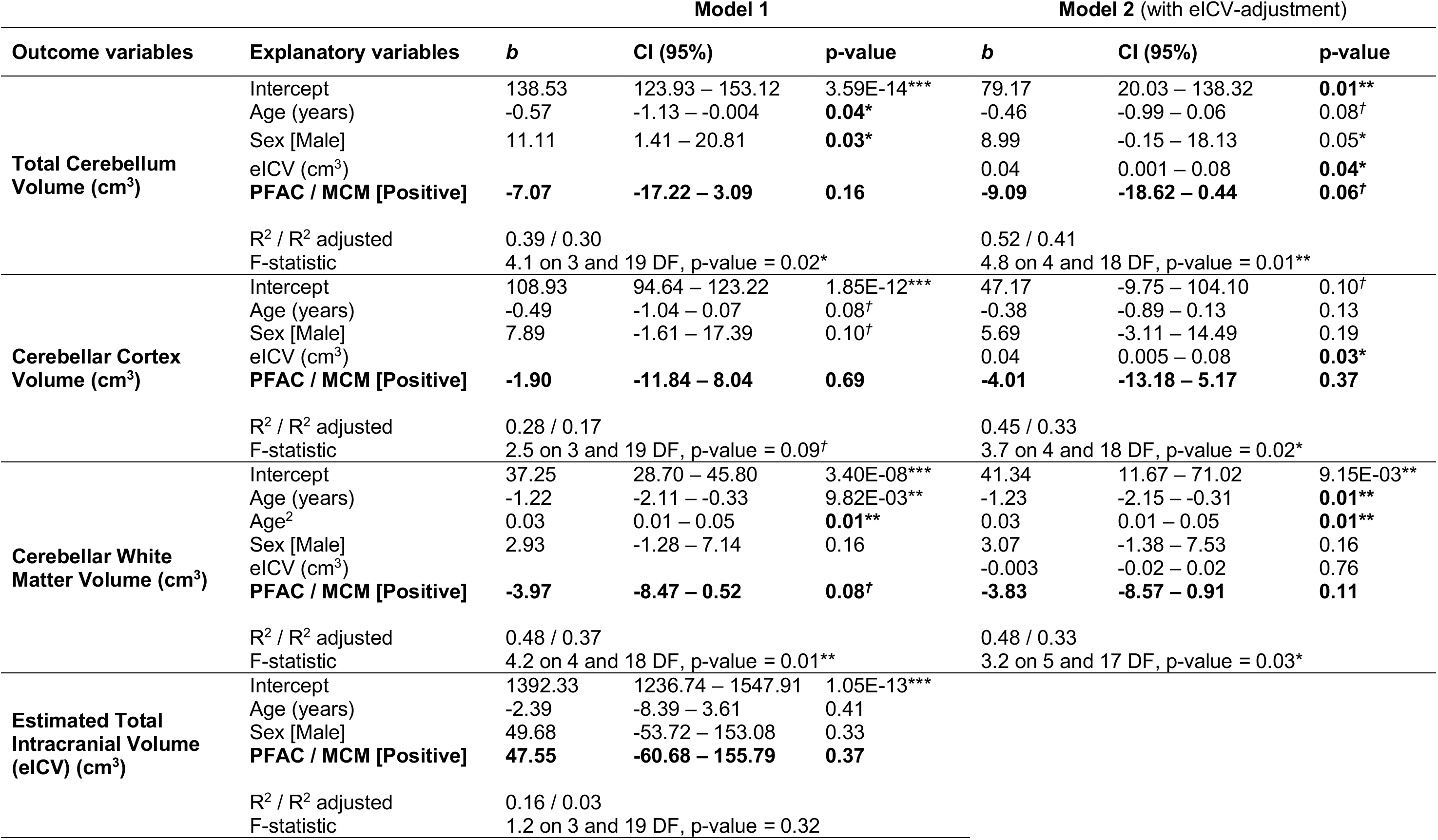
Exploratory analysis of the relationship between volumetric measures of interest and posterior fossa arachnoid cyst and mega cisterna magna findings among 3q29Del participants. Multiple linear regression results for total cerebellum volume, cerebellar cortex volume, cerebellar white matter volume and eICV indicate no significant relationship between the likelihood of PFAC/MCM findings and interrogated VOIs among 3q29Del participants (p’s > 0.05), while correcting for sex, age, age^2^ (if appropriate) and eICV (in model 2 only). The main effect of the binary PFAC/MCM variable is reported in bold for clarity. The age term/s included in each model reflect the best-fitting polynomial expansion of age for a given VOI selected by ANOVA. Major assumptions for ordinary least squares regression were met in the current analysis, hence additional heteroskedasticity-robust estimates were not calculated. F-statistics are reported to assess the overall significance of each model. 3q29Del N = 23. Contrast coding: reference levels for the PFAC/MCM and sex variables are negative and female, respectively. p-value ≤ 0.001 ‘***’, p-value ≤ 0.01 ‘**’, p-value ≤ 0.05 ‘*’, p-value ≤ 0.1 ‘^†^’. Abbreviations: 3q29 deletion syndrome, 3q29Del; volumetric measure of interest, VOI; posterior fossa arachnoid cyst, PFAC; mega cisterna magna, MCM; estimated total intracranial volume, eICV; analysis of variance, ANOVA; unstandardized coefficient estimate, b; confidence interval, CI; degrees of freedom, DF.

### Brain-behavior relationships in 3q29Del

We next investigated the functional relevance of these neuroimaging findings to sensorimotor and cognitive deficits in 3q29Del. The mean standardized scores for both VMI and composite IQ were approximately two standard deviations below normative means, indicating clinical impairment (Fig. S11, Table S9). Both VMI (*b* = 1.38, *p* ≤ 0.01, *FDR adjusted-p* = 0.02) and composite IQ (*b* = 1.33, *p* ≤ 0.01, *FDR adjusted-p* = 0.02) were positively associated with cerebellar white matter volume, while controlling for sex and age, but not with cerebellar cortex volume (*p’s* > 0.05) (Table S10).

In exploratory analyses, we aimed to differentiate the sensorimotor and cognitive subprocesses that may contribute to these findings. Cerebellar white matter volume was positively associated with both verbal (*b* = 1.51, *p* = 0.03) and non-verbal IQ (*b* = 1.28, *p* = 0.03) cognitive subtest scores, but not with visual perception or fine motor coordination skills (*p’s* > 0.05) (Table S10). In models where a significant brain-behavior relationship was identified (*p’s* ≤ 0.05), eICV was subsequently added as an additional covariate to consider the effect of head size. Cerebellar white matter volume remained significantly associated with VMI (*b* = 1.51, *p* ≤ 0.01), composite (*b* = 1.43, *p* ≤ 0.01), verbal (*b* = 1.59, *p* = 0.03), and non-verbal IQ (*b* = 1.36, *p* ≤ 0.01) after eICV-adjustment (Table 5, Fig. 6).

**Figure 6.**
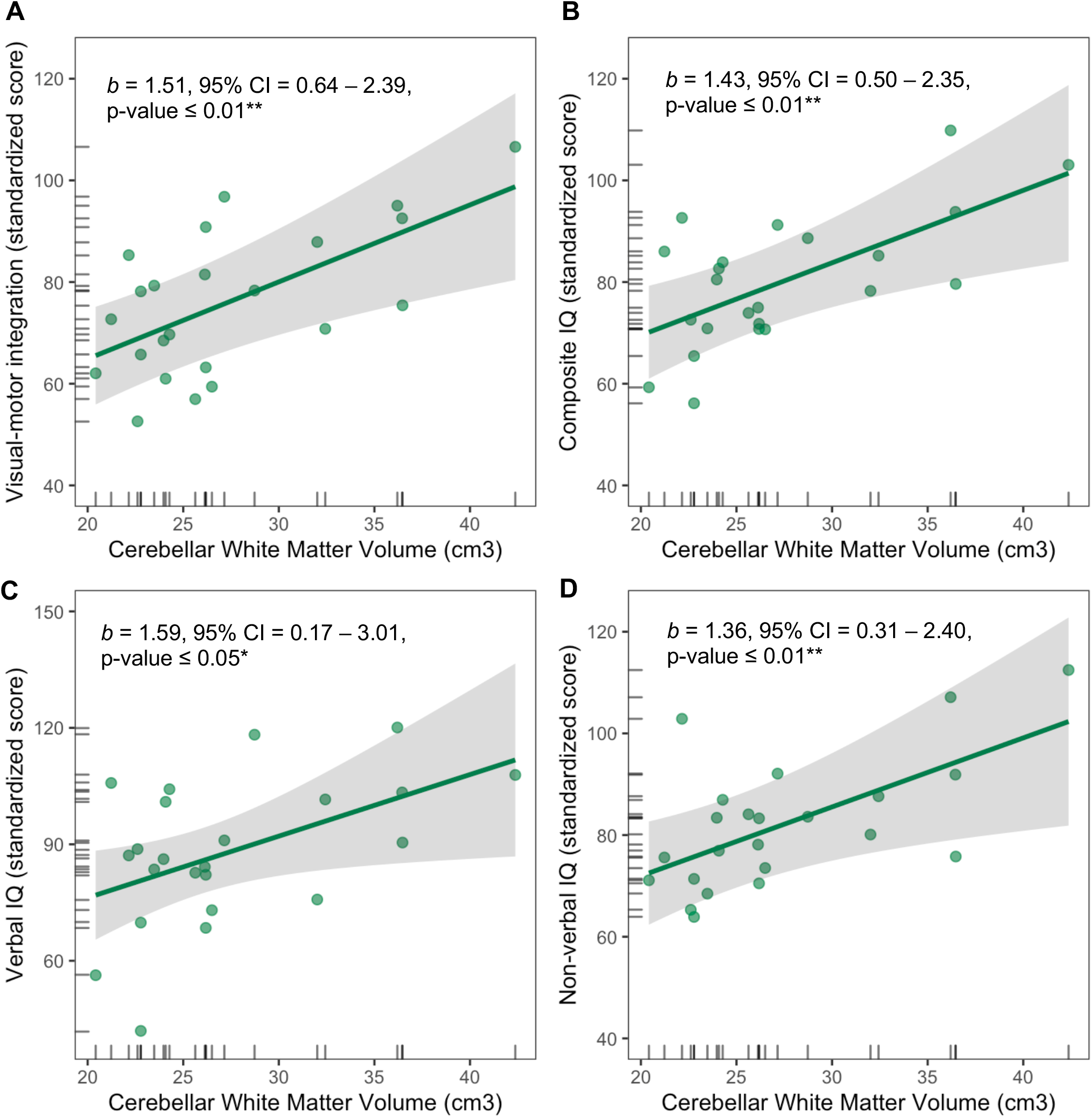
Predictor effect plots showing the relationships between cerebellar white matter volume and visual-motor integration skills, composite IQ, verbal IQ and non-verbal IQ among 3q29Del participants. **A-D)** Predicted values for visual-motor integration, composite IQ, verbal IQ, and non-verbal IQ scores were computed from the multiple linear regression models reported in Table 5, while covariates (sex, age, eICV) were held fixed. Error bands indicate the 95% confidence interval, data points represent partial residuals, and rug plots show the distribution of the variables. Parameter estimates for the main effect of cerebellar white matter volume are indicated on each plot and reflect heteroskedasticity-robust estimates. Regression results indicate significant relationships between cerebellar white matter volume and standardized test scores for **A**) visual-motor integration skills (*p* ≤ 0.01), **B**) composite IQ (*p* ≤ 0.01), **C**) verbal IQ (*p* ≤ 0.05), and **D**) non-verbal IQ (*p* ≤ 0.01), with larger volumes predicting higher scores among 3q29Del participants. 3q29Del *N* = 23. p-value ≤ 0.001 ‘***’, p-value ≤ 0.01 ‘**’, p-value ≤ 0.05 ‘*’, p-value ≤ 0.1 *‘^†^’ Abbreviations*: 3q29 deletion syndrome, 3q29Del; intelligence quotient, IQ; estimated total intracranial volume, eICV; unstandardized coefficient estimate, *b*; confidence interval, CI.

**Table 5.**
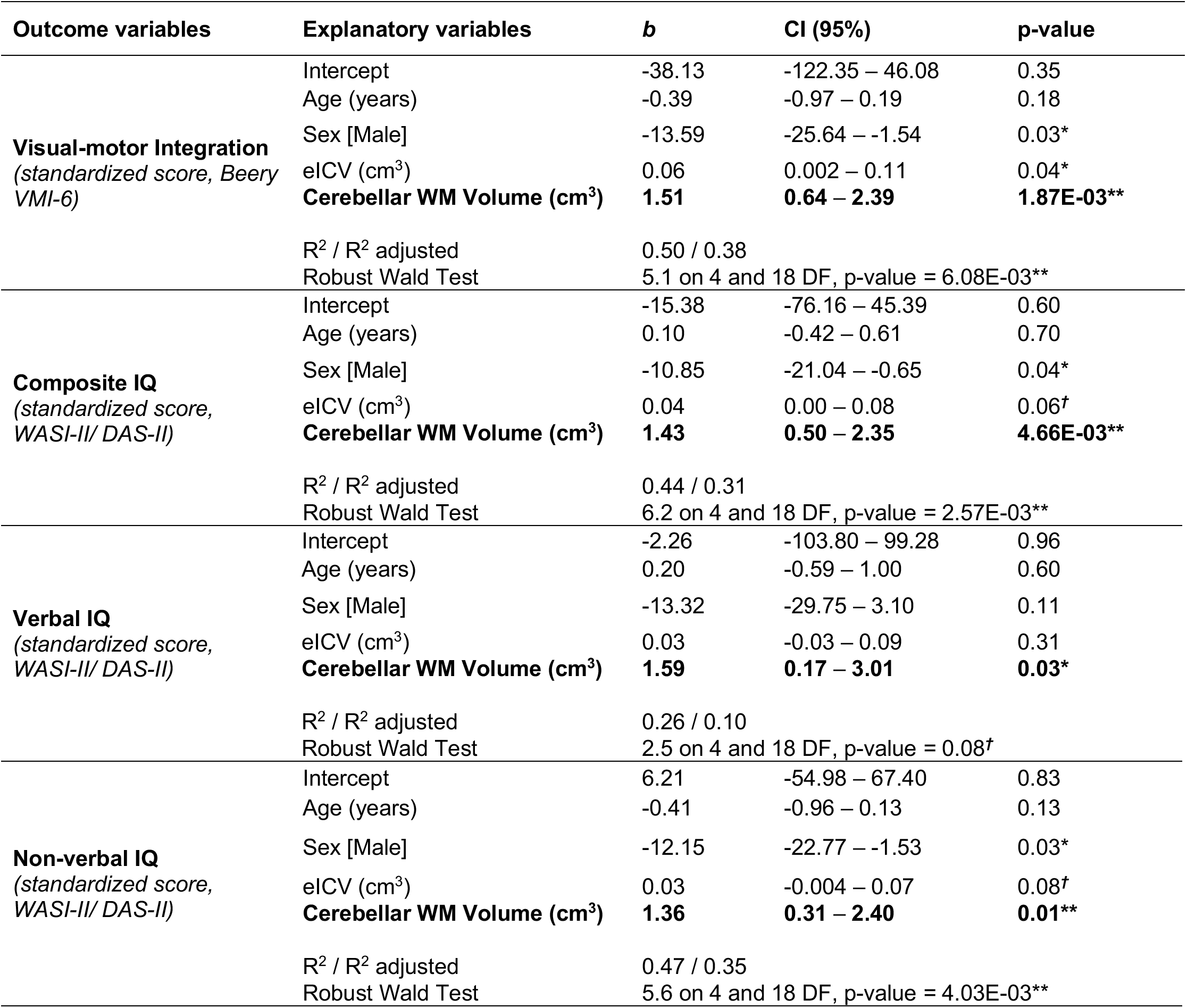
Summary of multiple linear regression results showing the relationships between cerebellar white matter volume and standardized test scores for sensorimotor and cognitive abilities among 3q29Del participants. Only the significant test results from our investigation of brain-behavior relationships are reported in this table for brevity (*p’s* ≤ 0.05) (see Table S10 for a complete summary of behavioral findings). Sex, age and eICV were included as covariates in each regression model. The main effect of cerebellar white matter volume is reported in bold for clarity. Regression parameters reflect heteroskedasticity-robust estimates. Robust Wald test statistics are reported to assess the overall significance of each model. Results indicate a significant relationship between cerebellar white matter volume and standardized test scores for visual-motor integration skills (*p* ≤ 0.01), composite IQ (*p* ≤ 0.01), verbal IQ (*p* ≤ 0.05), and non-verbal IQ (*p* ≤ 0.01) among 3q29Del participants. 3q29Del *N* = 23. Contrast coding: reference level for the categorical sex variable is female. p-value ≤ 0.001 ‘***’, p-value ≤ 0.01 ‘**’, p-value ≤ 0.05 ‘*’, p-value ≤ 0.1 ‘^†^’. *Abbreviations*: 3q29 deletion syndrome, 3q29Del; estimated total intracranial volume, eICV; intelligence quotient, IQ; WASI, Wechsler Abbreviated Scale of Intelligence; DAS, Differential Ability Scales; VMI, visual-motor integration; unstandardized coefficient estimate, *b*; confidence interval, CI; degrees of freedom, DF.

| Outcome variables | Explanatory variables | *b* | CI (95%) | p-value |
| --- | --- | --- | --- | --- |
| **Visual-motor Integration**<br>(standardized score, Beery VMI-6) | Intercept | -38.13 | -122.35 – 46.08 | 0.35 |
|  | Age (years) | -0.39 | -0.97 – 0.19 | 0.18 |
|  | Sex [Male] | -13.59 | -25.64 – -1.54 | 0.03* |
|  | eICV (cm<sup>3</sup>) | 0.06 | 0.002 – 0.11 | 0.04* |
|  | **Cerebellar WM Volume (cm<sup>3</sup>)** | **1.51** | **0.64 – 2.39** | **1.87E-03\*\*** |
|  | R<sup>2</sup> / R<sup>2</sup> adjusted<br>Robust Wald Test | 0.50 / 0.38<br>5.1 on 4 and 18 DF, p-value = 6.08E-03** |  |  |
| **Composite IQ**<br>(standardized score, WASI-II/ DAS-II) | Intercept | -15.38 | -76.16 – 45.39 | 0.60 |
|  | Age (years) | 0.10 | -0.42 – 0.61 | 0.70 |
|  | Sex [Male] | -10.85 | -21.04 – -0.65 | 0.04* |
|  | eICV (cm<sup>3</sup>) | 0.04 | 0.00 – 0.08 | 0.06<sup>†</sup> |
|  | **Cerebellar WM Volume (cm<sup>3</sup>)** | **1.43** | **0.50 – 2.35** | **4.66E-03\*\*** |
|  | R<sup>2</sup> / R<sup>2</sup> adjusted<br>Robust Wald Test | 0.44 / 0.31<br>6.2 on 4 and 18 DF, p-value = 2.57E-03** |  |  |
| **Verbal IQ**<br>(standardized score, WASI-II/ DAS-II) | Intercept | -2.26 | -103.80 – 99.28 | 0.96 |
|  | Age (years) | 0.20 | -0.59 – 1.00 | 0.60 |
|  | Sex [Male] | -13.32 | -29.75 – 3.10 | 0.11 |
|  | eICV (cm<sup>3</sup>) | 0.03 | -0.03 – 0.09 | 0.31 |
|  | **Cerebellar WM Volume (cm<sup>3</sup>)** | **1.59** | **0.17 – 3.01** | **0.03\*** |
|  | R<sup>2</sup> / R<sup>2</sup> adjusted<br>Robust Wald Test | 0.26 / 0.10<br>2.5 on 4 and 18 DF, p-value = 0.08<sup>†</sup> |  |  |
| **Non-verbal IQ**<br>(standardized score, WASI-II/ DAS-II) | Intercept | 6.21 | -54.98 – 67.40 | 0.83 |
|  | Age (years) | -0.41 | -0.96 – 0.13 | 0.13 |
|  | Sex [Male] | -12.15 | -22.77 – -1.53 | 0.03* |
|  | eICV (cm<sup>3</sup>) | 0.03 | -0.004 – 0.07 | 0.08<sup>†</sup> |
|  | **Cerebellar WM Volume (cm<sup>3</sup>)** | **1.36** | **0.31 – 2.40** | **0.01\*\*** |
|  | R<sup>2</sup> / R<sup>2</sup> adjusted<br>Robust Wald Test | 0.47 / 0.35<br>5.6 on 4 and 18 DF, p-value = 4.03E-03** |  |  |

We also assessed whether these behavioral relationships may have been partially confounded by shared variance (Fig. S12). VMI had a significant positive correlation with composite IQ (*r* = 0.62, *p* ≤ 0.01), and this relationship was explained by a significant correlation between VMI and non-verbal IQ (overlapping constructs) (*r* = 0.75, *p* ≤ 0.001). VMI did not correlate significantly with verbal IQ (*p* > 0.05), implying that VMI deficits do not explain the relationship between cerebellar white matter volume and verbal IQ in a significant way, and vice versa. Lastly, we found no significant differences between the standardized test scores of 3q29Del participants with versus without cystic/cyst-like malformations of the posterior fossa (*p’s* > 0.05) (Table S11), which supports the specificity of our behavioral findings to cerebellar changes.

## Discussion

It is now clear that the genetic component of many neurodevelopmental and psychiatric disorders includes CNVs arising from large structural rearrangements of the genome (10–18). However, there is a big gap in our understanding of the neurobiology underpinning many of these associations. To gain insights into pathogenesis, it is crucial to determine how these CNVs impact the structure and function of the human brain.

To this end, here we report the first *in vivo* quantitative neuroimaging study in individuals with 3q29Del: a recurrent CNV that confers exceptionally high genetic risk for neurodevelopmental and psychiatric disorders, including intellectual disability, ASD and the largest known effect size for SZ (10, 33–41). Using high-resolution MRI, standardized behavioral assessment tools, and a hypothesis-driven approach, our study revealed key findings on local alterations of brain structure and brain-behavior relationships in individuals with 3q29Del.

First, by volumetric analysis, we showed that the average size of the cerebellum is significantly smaller among 3q29Del participants compared with healthy controls. This difference remained significant after adjustment for eICV, which was itself smaller among 3q29Del participants, indicating that the magnitude of reduction observed in the cerebellar volumes of individuals with this CNV shows regional specificity. This result corroborates radiological findings from several case reports (39, 50, 92). In our previous work on the same 3q29Del sample, we identified cerebellar hypoplasia in 14 males and females with 3q29Del between the ages of six to 34 years (39). Citta et al. (2013) identified cerebellar “atrophy” in a female adolescent with 3q29Del, who had a history of intellectual disability and psychosis (50). Sargent et al. (1985) identified “absence” of the cerebellar vermis and “small” cerebellar hemispheres in a male infant with a 3q terminal deletion (proximal to 3q29) (92). Hence, our volumetric findings converge with qualitative findings from case reports and for the first time quantitatively highlight the cerebellum as a region of marked pathology in 3q92Del.

For a more nuanced characterization, we next separated the cerebellum into its two primary tissue-types and found that the 3q29Del group had smaller cerebellar cortex volumes than controls, both with and without adjustment for eICV. In contrast, absolute volumes of cerebellar white matter did not differ between the groups, indicating that volumetric reduction is localized to the cerebellar cortex, which contains almost all neuronal cell bodies in the cerebellum and more than half of the neurons in the entire adult brain (93, 94). Surprisingly, 3q29Del participants also had larger cerebellar white matter volumes than controls after eICV-adjustment, implying that cerebellar white matter, which mostly contains myelinated axons, exhibits volumetric expansion in 3q29Del relative to what would be expected from controls with the same head size. Furthermore, the volumetric ratio of cerebellar cortex to white matter was smaller among 3q29Del participants, confirming that changes in these two cerebellar structures are nonuniform.

Unlike cerebral structures, the normative development of the cerebellum has been investigated in only a small number of studies. Based on this evidence, the human cerebellum starts differentiation during the first trimester (95) and displays a prolonged developmental course, with cerebellar cortex and white matter following distinct trajectories (96–101). In our large sample of healthy controls, cross-sectional modeling of cerebellar volumes revealed patterns consistent with earlier findings. From childhood to adulthood, the typical volume of cerebellar cortex roughly made an inverted U-shape, which is thought to reflect a period of transient exuberance followed by synaptic pruning (102). In contrast, the typical volume of cerebellar white matter showed a concomitant expansion continuing into adulthood, which is thought to reflect an increase in myelination and axonal diameter (102). It is conceivable that idiosyncrasies in the rate and type of these neuromaturational processes confer different susceptibilities to cerebellar cortex versus white matter development, which may partially explain the opposite direction of volumetric changes observed in these tissue-types in our 3q29Del sample.

It is also plausible that the expression of 3q29 genes themselves differ between cerebellar cortex and white matter, which if true, would lead to tissue-specific consequences upon their hemizygosity. Accordingly, we queried the Human Protein Atlas (https://www.proteinatlas.org) (103, 104) for the 21 genes located in the 3q29 interval and found that 11 were detected in the cerebellar cortex at the protein level (Fig. S13). Notably, *BDH1*, *DLG1* and *PCYT1A* showed high protein expression profiles in the granular or Purkinje layers of the cerebellar cortex. *DLG1* encodes a synaptic scaffolding protein that interacts with AMPA and NMDA receptors (105), which are key components of glutamatergic synapses and mediators of synaptic plasticity. *BDH1 and PCYT1A* are involved in ketone body metabolism (106) and phosphatidylcholine biosynthesis (107), which are important regulators of bioenergetic homeostasis. Coincidently, both *DLG1* and *BDH1* were previously proposed as drivers of psychiatric and neurodevelopmental phenotypes in this CNV (43, 44). We conjecture that dosage alterations in these genes may be especially detrimental for cerebellar cortex development and function.

At the same time, *DLG1* was also detected in cerebellar white matter at the protein level (Fig. S13). Interestingly, *DLG1* was shown to regulate myelin thickness in the peripheral nervous system (108). Assuming a similar mechanism of action, larger cerebellar white matter volumes in our 3q29Del sample may reflect a failure in *DLG1*’s ability to put a break on myelination, although we highlight that haploinsufficiency of *Dlg1* alone was not sufficient to recapitulate the mouse-specific phenotypes of 3q29Del in an earlier study (109). Overall, these data lend molecular support to our volumetric findings implicating the 3q29 locus in cerebellar development.

It is also worth noting that larger cerebellar white matter volumes in 3q29Del may reflect a compensatory mechanism (as opposed to a primary disturbance) originating either internally within the white matter network of the cerebellum or externally through afferent pathways. Oligodendrocyte precursor cells continuously survey their environment and have the capacity to differentiate into oligodendrocytes well into adulthood, which can regenerate myelin following injury or disease (110–114). Hence, structural white matter alterations can serve compensatory functions by modifying conduction velocity. Altogether, mechanisms that may underly our structural findings include changes in number of neurons, neuronal size or dendritic arborization for cerebellar cortex, and changes in axon growth, guidance, or myelination for white matter. Future work will aim to elucidate the relative contribution of these potential mechanisms to altered cerebellar development in 3q29Del.

Another key finding was the relationships identified between tissue-specific cerebellar volumes and sensorimotor and cognitive abilities among 3q29Del participants. Smaller cerebellar white matter volumes were associated with worse VMI, and composite, verbal, and non-verbal IQ scores. Interestingly, neither visual perception nor fine motor coordination skills separately had a significant relationship with cerebellar white matter, suggesting that the identified sensorimotor link is specific to the ability to integrate information from multiple modalities, as opposed to performing either one alone. Importantly, VMI and verbal IQ were not significantly correlated with each other in this sample, indicating that cerebellar associations identified with these motor and non-motor abilities are not secondary consequences of one another. Cumulatively, these findings suggest that structural cerebellar changes have functional significance for specific behavioral phenotypes in 3q29Del.

The role of the cerebellum in sensorimotor integration is long established (115). However, cerebellar contributions to cognitive processing have traditionally received less attention, in part due to a cortico-centric view of “higher” functions (116). A major shift has begun towards re-conceptualizing the cerebellum as a calibrator (i.e., “feed-forward controller”) of the output and possibly the development of not only motor but also non-motor systems (57-59, 117-125), although skepticism about cerebellar roles beyond motor control remains. Recent anatomical studies have identified strong connections between the cerebellum and non-motor brain regions (117, 123, 126–128). Consistent with this, cerebellar activation has been reported in various cognitive tasks in functional neuroimaging (129–135), and cerebellar damage has been shown to produce impairments in linguistic processing, spatial cognition and executive functions (136–139).

In recent years, the cerebellum has also emerged as a site of renewed interest for a broad range of neurodevelopmental and psychiatric illnesses (58, 140, 141). Two independent studies have shown that structural alterations within the cerebellum are associated with general liability for common psychopathology (142, 143). Supporting these findings, perinatal injury to the cerebellum is now considered the largest known nongenetic risk factor for ASD (58, 144–147). Cerebellar abnormalities have also been reported in idiopathic SZ, ASD and ADHD (148–160), although findings in this area show strong heterogeneity likely due to technical and physiological causes. Despite these advances, the precise molecular and neurobiological basis of cerebellar contributions to non-motor behaviors and mental illnesses remains largely unknown.

Our findings indicate that 3q29Del sits at the intersection of abnormal cerebellar development and increased risk for neurodevelopmental and psychiatric disease, which converges with growing evidence implicating the cerebellum in the pathophysiology of several other genomic variants, including well-known ASD or SZ-related genes such as *TSC1*, *FMR1*, *SHANK3*, *MECP2*, and *PTEN*, and CNVs like 22q11.2 deletion (161–167). The explanatory gap between these loci and disease mechanism has not yet been fully bridged, however altered cerebellar development may be a shared neuroanatomical endophenotype that contributes to heightened risk for a variety of neurodevelopmental and psychiatric disorders across these variants. Future work should investigate whether the biological mechanisms disrupted in 3q29Del converge onto pathways that are vulnerable to other genetic variants associated with cerebellar abnormalities.

Finally, our findings also indicate that cystic/cyst-like malformations of the posterior fossa show an elevated prevalence among 3q29Del participants compared with controls. The prevalence of PFAC/MCM findings was approximately 1% among controls, and more common among males than females, consistent with prior estimates in the general population (55, 56). In contrast, their prevalence was approximately 54% among 3q29Del participants, with no sex differences. Consequently, besides cerebellar abnormalities, 3q29Del also confers a greater risk for cystic/cyst-like malformations in this region, suggesting a general vulnerability of the posterior fossa to this CNV. However, the likelihood of PFAC/MCM was not associated with cognitive or sensorimotor phenotypes in 3q29Del, supporting the specificity of behavioral associations to the cerebellum.

Notably, we found that 3q29Del confers greater influence on risk for these cystic/cyst-like malformations among females than males. In prior work, we also identified a reduction in the male:female bias among individuals with 3q29Del in ASD rates (38). In the general population the ratio of males:females with ASD is approximately 4:1 (168–170); in 3q29Del this ratio is 2:1 (38). According to the liability threshold model, females require a larger genetic burden than males before reaching affection status (171, 172), and the sex ratio in disease prevalence tends to approach 1:1 as the severity of a mutation increases (173). 3q29Del is on the more severe end of this genetic spectrum, which may explain why the rate of PFAC/MCM is similar among males and females with this CNV. That said, we found a greater reduction in eICV among male than female 3q29Del participants, but no robust sex by diagnostic group interaction effect on cerebellar volumes. These results suggest regional variability in patterns of sexual dimorphism in 3q29Del and highlight that sex must be considered an important variable in studies of this CNV.

Several limitations should be addressed in future research. First, due to its rare prevalence, the 3q29Del sample size was limited, which prevented an in-depth analysis of complex interactions involving age and sex. Replication is needed in a larger sample of cases imaged on the same scanner as controls. Demands of study participation may have barred individuals with more severe disease manifestations from participating; this may have led to underestimation of effect sizes. For MRI, the complex architecture of the cerebellum introduced methodological challenges; future work with improved segmentation techniques will explore finer parcellations. Analyses will also be expanded to clinical phenotypes, and other measures of cognition for a more comprehensive investigation. Importantly, cerebellar connections with cerebral regions warrant investigation to expand questions to the circuitry-level. Finally, given our cross-sectional approach, reported findings do not necessarily reflect causation. Leveraging the mouse model of 3q29Del (109) may help elucidate mechanistic impacts on cerebellar development.

## Data Availability

The 3q29Del data collected in this study are deposited in the NIMH Data Archive (nda.nih.gov) (behavioral data: collection 2614, embargoed until September 2022; neuroimaging data: collection 3126, embargoed until November 2023). Prior to these dates, the 3q29Del data are available from the corresponding author upon reasonable request. Access to the Human Connectome Project data used in the present study is subject to Open Access and Restricted Data Use Terms for the HCP-Young Adult dataset (1200 subjects data release; minimally preprocessed structural data available from the Connectome Coordination Facility via: https://humanconnectome.org/study/hcp-young-adult/document/1200-subjects-data-release) and the NIMH Data Archive Data Use Certification Agreement for the HCP-Development dataset (HCP-Development Lifespan 2.0 release; minimally preprocessed structural data available from the NIMH Data Archive via: https://nda.nih.gov/general-query.html?q=query=featured-datasets:HCP%20Aging%20and%20Development).

## Acknowledgements

Acknowledgements and disclosures

We acknowledge the 3q29Del study population, their families and the 3q29 Project members. We thank the Marcus Autism Center for providing clinical assessment resources. Funding for 3q29Del work was provided by National Institutes of Health (NIH) grants R01 MH110701, R01 MH118534, and National Institutes of Mental Health (NIMH) grant U01MH081988. REDCap is supported by grant UL1 TR000424. Healthy control data for the young-adult age range of the present study were provided in part by the Human Connectome Project, WU-Minn Consortium (Principal Investigators: David Van Essen and Kamil Ugurbil; 1U54MH091657) funded by the 16 NIH Institutes and Centers that support the NIH Blueprint for Neuroscience Research; and by the McDonnell Center for Systems Neuroscience at Washington University. Healthy control data for the developmental age range of the present study were provided by the Lifespan Human Connectome Project, supported by the NIMH under Award Number U01MH109589. The content is solely the responsibility of the authors and does not necessarily represent the official views of the NIH.

The authors have no competing financial interests related to this study to declare.

## Data availability

The 3q29Del data collected in this study are deposited in the NIMH Data Archive (nda.nih.gov) (behavioral data: collection 2614, embargoed until September 2022; neuroimaging data: collection 3126, embargoed until November 2023). Prior to these dates, the 3q29Del data are available from the corresponding author on reasonable request. Access to the Human Connectome Project data used in the present study is subject to the WU-Minn HCP Consortium’s Open Access and Restricted Data Use Terms for the HCP-Young Adult dataset (1200 subjects data release; minimally preprocessed structural data available from the Connectome Coordination Facility via: https://humanconnectome.org/study/hcp-young-adult/document/1200-subjects-data-release) and the NIMH Data Archive’s Data Use Certification Agreement for the HCP-Development dataset (HCP-Development Lifespan 2.0 release; minimally preprocessed structural data available from the NIMH Data Archive via: https://nda.nih.gov/general-query.html?q=query=featured-datasets:HCP%20Aging%20and%20Development).

