## Supplemental Materials for "Structural deviations of the posterior fossa and the cerebellum and their cognitive links in a neurodevelopmental deletion syndrome"

*Table of contents*

**Supplemental figures and tables**……………………………………………………………………………3-41

Table S1. Demographic characteristics of the study sample in volumetric analyses, stratified by diagnostic group and sex……………………………………………………………………………………………3

Table S2. Comparison of structural magnetic resonance imaging (MRI) protocols……………………4-5

Fig. S1. Example cerebellar segmentation masks for representative age- and sex-matched 3q29Del and healthy control pairs………………………………………………………………………………………6-7

Table S3. Extended linear regression results testing the effect of diagnostic group on volumetric measures of interest and polynomial modeling of age………………………………………………………12-19

Fig. S6. Regression diagnostics: testing the assumptions of ordinary least squares regression for best-fitting models from Table S3……………………………………………………………………20-21

Table S4. Summary of supplemental results from penalized cubic spline models testing the effect of diagnostic group on volumetric measures of interest……………………………………………………22

Fig. S7. Developmental trajectories for volumetric measures of interest estimated by the penalized cubic spline approach……………………………………………………………………………………23

Table S5. Exploratory modeling of diagnostic group by sex interaction effects on volumetric measures of interest…………………………………………………………………………………………24-26

Fig. S8. Predictor effect plot showing a suggestive interaction effect between diagnostic group and sex on eICV-adjusted cerebellar white matter volumes…………………………………………………………26

Table S6. *Post hoc* analysis of the suggestive sex by diagnostic group interaction effect on eICV-adjusted cerebellar white matter volumes………………………………………………………………………27

Table S7. Summary of multiple linear regression findings and descriptive statistics for volumetric measures of interest in 3q29Del and control groups…………………………………………………………28-29

Fig. S9. Estimated normative percentile curves for cerebellar volumes and eICV, stratified by sex…..30-32

Table S10. Extended multiple linear regression results testing the relationships between tissue-specific cerebellar volumes and sensorimotor and cognitive abilities among 3q29Del participants ……………36-37

Fig. S12. Heatmap visualization of pairwise Pearson's correlations between standardized test scores for sensorimotor and cognitive abilities among 3q29Del participants ……………………………………38

Table S11. Standardized test scores for sensorimotor and cognitive abilities in 3q29Del participants with versus without posterior fossa arachnoid cyst or mega cisterna magna findings …………….………...…39
Fig. S13. Tissue and cell-type specific protein expression profiles of 3q29 interval genes annotated by the Human Protein Atlas.………………..……………………..………………………………..………...…….40

**Supplemental methods**……………………………..……………………………………………………………41-47

Extended methods for processing and quality control of structural MRI data……………………………..…..41
Extended methods for radiological evaluation of structural MRI data……………………………...…..41-42
Details on standardized behavioral measures…………………………………………………………...........42
Extended statistical methods for penalized cubic spline and quantile spline models……...………..…42-46
List of R packages used for statistical analyses and diagnostics……………………………………….........46
References for supplemental methods……………………………………………………………….……...47-48

### Supplemental figures and tables

| Demographic variables | Males | | | Females | | |
|---|---|---|---|---|---|---|
| | **Control** *N* = 747 | **3q29Del** *N* = 14 | **Test statistics** | **Control** *N* = 861 | **3q29Del** *N* = 9 | **Test statistics** |
| **Age** (in years) | | | | | | |
| Mean ± SD | 22.38 ± 7.74 | 14.14 ± 9.03 | *r* = 0.13 (small effect size), *W* = 8146.5, p-value [a] = 3.40E-04*** | 23.03 ± 8.60 | 16.56 ± 9.86 | *r* = 0.07 (small effect size), *W* = 5422, p-value [a] = 0.04* |
| Median | 24.00 | 13.00 | | 25.00 | 15.00 | |
| [Range] | [5 – 37] | [4 – 39] | | [6 – 36] | [6 – 34] | |
| **Ethnicity**[#], *N (%)* | | | | | | |
| Non-Hispanic / Latino | 64 (86.60%) | 13 (92.86%) | $X^2$ = 0.08, DF = 1, p-value [b] = 0.78 | 757 (98.31%) | 9 (100%) | $X^2$ = 0.25, DF = 1, p-value [b] = 0.62 |
| Hispanic / Latino | 99 (13.40%) | 1 (7.14%) | | 91 (11.82%) | 0 (0%) | |
| **Race**[##], *N (%)* | | | | | | |
| White | 52 (71.53%) | 13 (92.86%) | $X^2$ = 3.83, DF = 4, p-value [b] = 0.43 | 587 (69.88%) | 8 (88.89%) | $X^2$ = 2.43, DF = 4, p-value [b] = 0.66 |
| Black / African American | 97 (13.21%) | 0 (0%) | | 125 (14.88%) | 0 (0%) | |
| Asian / Native Hawaiian / Other Pacific Islander | 55 (7.49%) | 0 (0%) | | 53 (6.31%) | 0 (0%) | |
| American Indian / Alaskan Native | 3 (0.41%) | 0 (0%) | | 1 (0.12%) | 0 (0%) | |
| More than one race | 54 (7.36%) | 1 (7.14%) | | 74 (8.81%) | 1 (11.11%) | |

**Table S1. Demographic characteristics of the study sample in volumetric analyses, stratified by diagnostic group and sex.** To supplement the models that explore the sex-specific effects of 3q29Del on VOIs, we report here the demographic characteristics of the study sample stratified by diagnostic group and sex. While there was a near complete overlap between the age ranges of the two diagnostic groups in each sex, there was a significant age difference between male 3q29Del participants and male controls ($p \leq 0.001$), and female 3q29Del participants and female controls on average ($p \leq 0.05$). There were no significant differences in the ethnicity or race compositions of the two diagnostic groups in either sex (*p's* > 0.05). Effect sizes are reported for significant test results only. Non-parametric statistics are reported in cases where the data do not meet parametric assumptions. [#]Male control *N* = 739, Female control *N* = 848 for the ethnicity variable due to missing data. [##]Male control *N* = 734, Female control *N* = 840 for the race variable due to missing data. Corresponding percentages reflect the fraction of male or female controls with complete data. [a]Wilcoxon rank sum test with continuity correction, [b]Pearson's chi-squared test with Yates' continuity correction. p-value ≤ 0.001 '***', p-value ≤ 0.01 '**', p-value ≤ 0.05 '*', p-value ≤ 0.1 *'†'* *Abbreviations*: 3q29 deletion syndrome, 3q29Del; volumetric measure of interest, VOI; standard deviation, SD; degrees of freedom, DF.

| | **3q29Del**<br>*N* = 24 | **HCP Young Adult**<br>*N* = 1,113 | **HCP Development**<br>*N* = 652 |
|---|---|---|---|
| **MRI Hardware** | | | |
| Scanner | Siemens Magnetom Prisma 3T | Siemens Magnetom Skyra 3T (Customized 3T "Conectom") | Siemens Prisma 3T |
| Max gradient strength | 80 mT/m gradient coil | 100 mT/m gradient coil | 80 mT/m gradient coil |
| Head coil | Siemens 32-channel Prisma head coil | Siemens 32-channel standard head coil | Siemens 32-channel Prisma head coil |
| **Acquisition parameters \| T1- & T2-weighted structurals** | | | |
| Slice thickness (mm) | 0.8 | 0.7 | 0.8 |
| vNavs for prospective motion correction | NO | NO | YES |
| FOV read (mm) | 256 | 224 | 256 |
| FOV phase (%) | 93.8 | 100 | 93.8 |
| Base resolution | 320 | 320 | 320 |
| Slabs | 1 | 1 | 1 |
| Slices per slab | 208 | 256 | 208 |
| Slice orientation | Sagittal | Sagittal | Sagittal |
| PAT mode | GRAPPA | GRAPPA | GRAPPA |
| Acceleration Factor PE | 2 | 2 | 2 |
| Reference lines PE | 32 | 32 | 32 |
| **Acquisition parameters \| T1-weighted structural** | | | |
| Pulse sequence | 3D single-echo T1-weighted MPRAGE | 3D single-echo T1-weighted MPRAGE | 3D multi-echo T1-weighted MPRAGE |
| TR (ms) | 2400 | 2400 | 2500 |
| TE (ms) | 2.2 | 2.1 | 1.8 / 3.6 / 5.4 / 7.2 |
| TI (ms) | 1000 | 1000 | 1000 |
| Flip angle (degree) | 8 | 8 | 8 |
| Coil elements | HEA; HEP | HEA; HEP | HEA; HEP |
| Bandwidth (Hz/Px) | 220 | 210 | 744 / 744 / 744 / 744 |
| Fat suppression | Water excitation fast | Water excitation fast | Water excitation fast |
| Scan time (min:sec) | 6:38 | 7:40 | 8:22 |
| **Acquisition parameters \| T2-weighted structural** | | | |
| Pulse sequence | 3D T2-weighted SPACE | 3D T2-weighted SPACE | 3D T2-weighted SPACE |
| TR (ms) | 3200 | 3200 | 3200 |
| TE (ms) | 563 | 565 | 564 |
| Flip angle mode | variable | variable | variable |
| Coil elements | HC1-7; NC1,2 | HEA; HEP | HEA; HEP |
| Bandwidth (Hz/Px) | 744 | 744 | 744 |
| Turbo factor | 314 | 314 | 314 |
| Fat suppression | None | None | None |
| Scan time (min:sec) | 6:38 | 7:40 | 8:22 |
| **Other** | | | |
| Data release version | N/A | "1200 subjects data release" - minimally processed format | "HCP-Development Lifespan 2.0 release" - minimally processed format |
| Structural preprocessing pipeline | HCP "minimal pre-processing" pipeline (v4.1.3 with FreeSurfer v6.0) (Glasser et al., 2013, Harms et al., 2018) | HCP "minimal pre-processing" pipeline (v3.21 with FreeSurfer v5.3.0-HCP) (Glasser et al., 2013, Harms et al., 2018) | HCP "minimal pre-processing" pipeline (v4.3.0 with FreeSurfer v6.0) (Glasser et al., 2013, Harms et al., 2018) |
| QC on structural preprocessing outputs | YES | YES | YES |
| Manual editing after automated segmentation | NO | NO | NO |

**Table S2. Comparison of structural magnetic resonance imaging (MRI) protocols.** A detailed report of the imaging protocols and relevant parameters used in the 3q29Del, HCP Young Adult and HCP Development datasets are provided in this table for increased transparency. HCP Young Adult and Development datasets were pooled to derive the control dataset; their corresponding imaging protocols were previously shown to be largely congruent, with most differences rooted in challenges related to scanning developmental populations (Harms et al., 2018). Note that the original "1200 Subjects Release" by the HCP Young Adult project includes structural MRI scans for *N* = 1,113 participants. A subset of these data comes from monozygotic twins, who have been previously reported to exhibit high to moderate correlations in brain morphology (e.g., White et al., 2002). To minimize bias in our standard error estimates, one sibling from each known monozygotic twin-pair that was scanned by the HCP Young Adult project was removed from the original dataset in the present study (*N* = 154 monozygotic twin-pairs based on available genotyping and/or self-report data**;** in cases of discrepancy between genotyping and self-report data, we relied on genetically verified data for final filtering). We additionally removed all participants with unknown zygosity information *(N* = 3) from the HCP Young Adult dataset for stringency. Hence, the final sample size of the HCP Young Adult data included in the present study was *N* = 956. Note that there were no monozygotic twin-pairs in the 3q29Del dataset. Zygosity information was not publicly available for the HCP Development dataset. Importable imaging protocols for all HCP datasets are available at https://www.humanconnectome.org/hcp-protocols. *Abbreviations*: 3q29 deletion syndrome, 3q29Del; Human Connectome Project, HCP; magnetization-prepared rapid gradient-echo, MPRAGE; Sampling perfection with application optimized contrast using different angle evolutions, SPACE; repetition time, TR; echo time, TE; inversion time, TI; volumetric navigators, vNavs; Field-of-view, FOV; parallel acquisition technique, PAT; phase-encoding, PE; generalized auto-calibrating partial parallel acquisition, GRAPPA; quality control, QC.

Harms MP, Somerville LH, Ances BM, Andersson J, Barch DM, Bastiani M, et al. (2018): Extending the Human Connectome Project across ages: Imaging protocols for the Lifespan Development and Aging projects. *Neuroimage*. 183:972-984.

Glasser MF, Sotiropoulos SN, Wilson JA, Coalson TS, Fischl B, Andersson JL, et al. (2013): The minimal preprocessing pipelines for the Human Connectome Project. *Neuroimage*. 80:105-124.

White T, Andreasen NC, Nopoulos P (2002): Brain volumes and surface morphology in monozygotic twins. *Cereb Cortex*. 12:486-493.

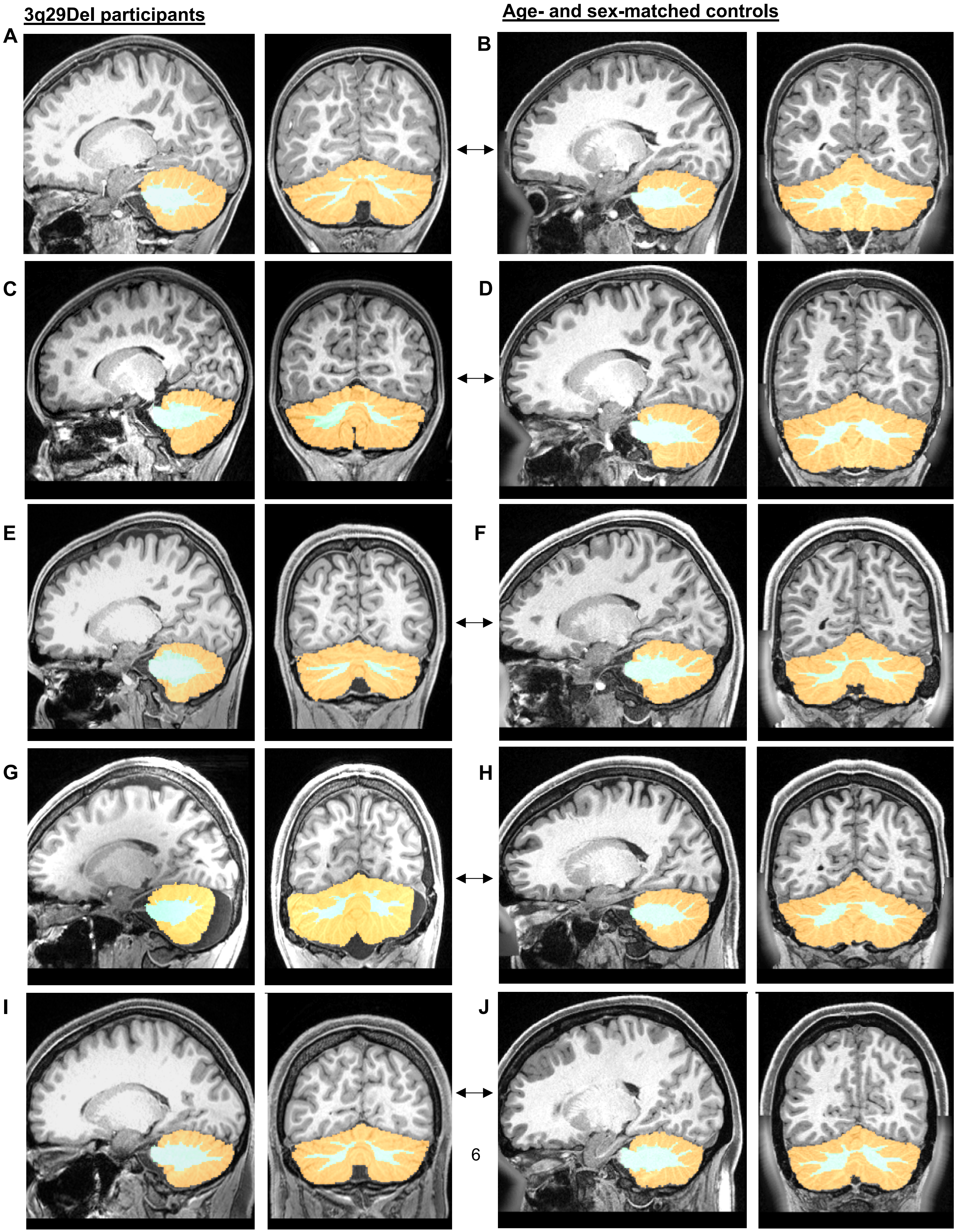
3q29Del participants
Age- and sex-matched controls
A
B
C
D
E
F
G
H
I
J

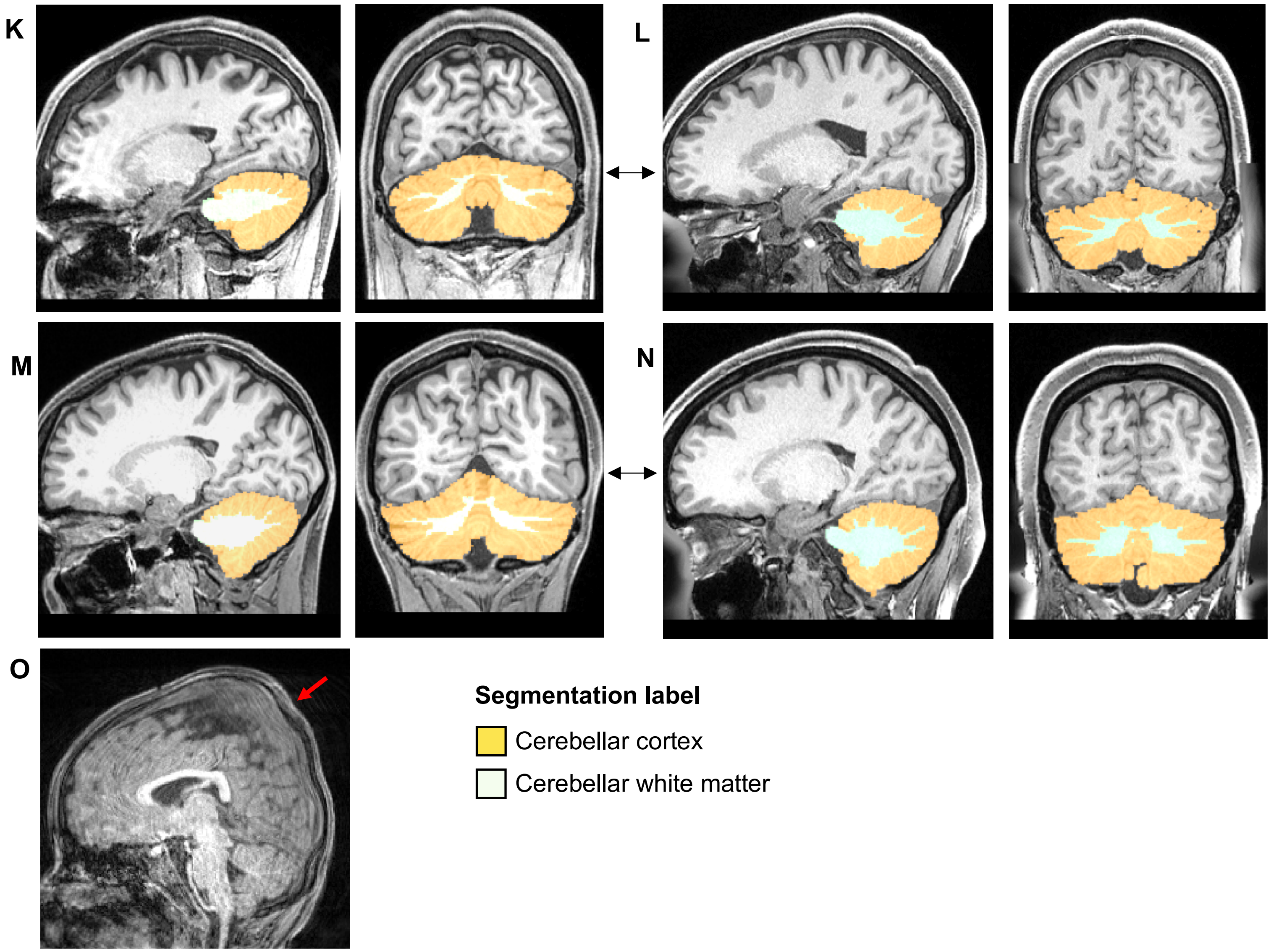

**Fig. S1. Example cerebellar segmentation masks for representative age- and sex-matched 3q29Del and healthy control pairs. A-N)** For quality control, cerebellar cortex and white matter segmentation masks were overlaid on T1-weighted MR images acquired from $N$ = 23 age- and sex-matched case-control pairs randomly selected from the dataset. Representative images for seven randomly selected case-control pairs are provided in this figure for illustration in sagittal and coronal planes. 3D volumes were segmented using FreeSurfer's automated subcortical segmentation algorithm in both diagnostic groups, as implemented in the HCP "minimal pre-processing" pipeline (Glasser et al., 2013; Harms et al., 2018). Image quality, tissue contrast and the boundaries of delineation for cerebellar voxel classifications were examined by two independent raters using the Connectome Workbench tool (https://www.humanconnectome.org/software/get-connectome-workbench). These outputs were found to be highly consistent within and between diagnostic groups, independent of age, sex, and presence or absence of radiologically observable posterior fossa abnormalities. **O)** A representative T1-weighted sagittal MR image of male 3q29Del participant who was excluded from volumetric analyses due to motion artifact and a skull deformity interfering with reliable volume estimations. Upon radiological examination, this 3q29Del participant was found to have plagiocephaly (red arrow). An example MR image is provided to illustrate our basis for exclusion. *Abbreviations*: 3q29 deletion syndrome, 3q29Del; years old, y/o.

Harms MP, Somerville LH, Ances BM, Andersson J, Barch DM, Bastiani M, et al. (2018): Extending the Human Connectome Project across ages: Imaging protocols for the Lifespan Development and Aging projects. *Neuroimage*. 183:972-984.

Glasser MF, Sotiropoulos SN, Wilson JA, Coalson TS, Fischl B, Andersson JL, et al. (2013): The minimal preprocessing pipelines for the Human Connectome Project. *Neuroimage*. 80:105-124.

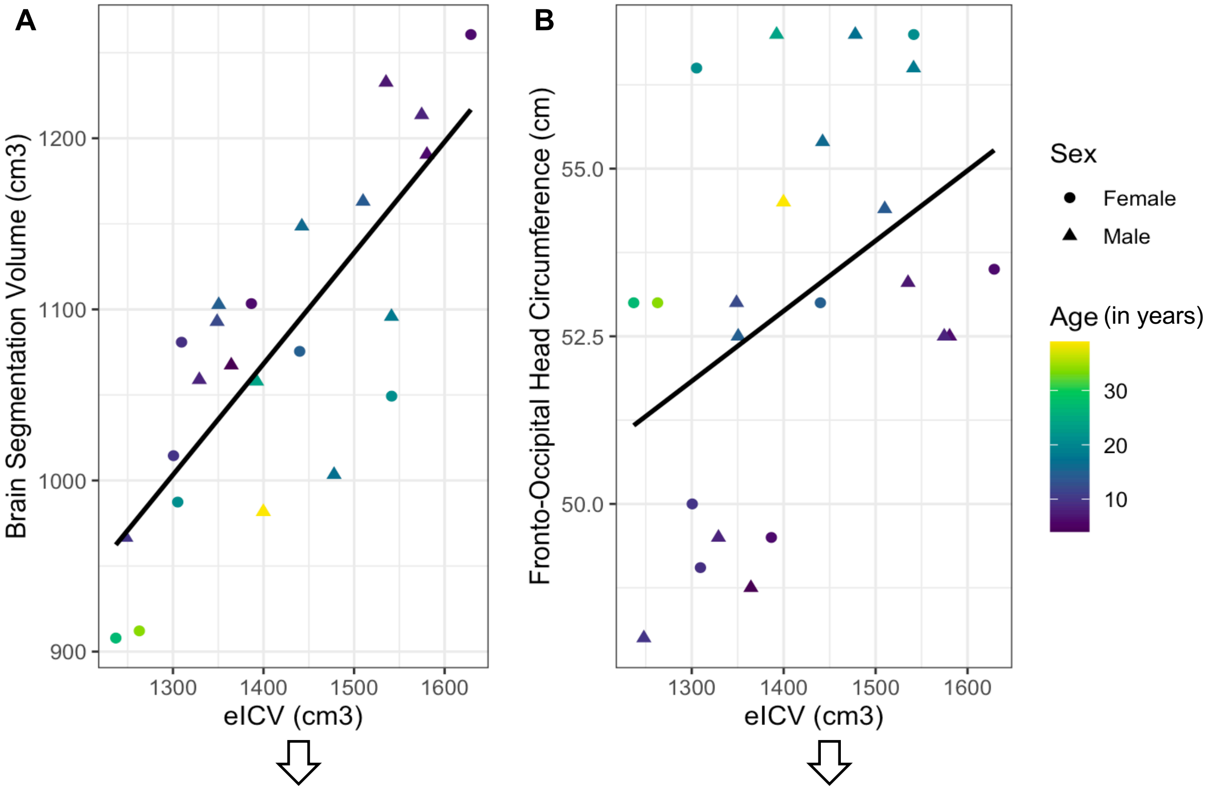

Pearson's *r* = 0.79
p-value = 6.50E-06***

Partial correlation after controlling for age and sex:

Pearson's *r* = 0.82
p-value = 6.55E-06***

Pearson's *r* = 0.43
p-value = 0.04*

Partial correlation after controlling for age and sex:

Pearson's *r* = 0.71
p-value = 2.92E-04***

**Fig. S2. Relationships between estimated total intracranial volume, total brain volume and head circumference among 3q29Del participants.** Morphometric analysis of regional brain structure confronts the challenge of accounting for head size variation. A widely used automated procedure for head size correction, which has been validated against manually delineated measurements of total intracranial volume (ICV), is the atlas-based head size normalization technique developed by FreeSurfer. This method exploits the relationship between ICV and the linear transform to MNI space to calculate an estimated total intracranial volume (eICV) for each participant, as described in Buckner et al. (2004). Since direct quality control of ICV segmentations is not attainable in this framework (due to lack of available segmentation masks), we assessed the quality of our eICV data by testing the Pearson's correlations between **A)** eICV and total brain volume (i.e., the "brain segmentation volume" label in FreeSurfer: volume of all voxels that are not background / brain stem) and **B)** eICV and fronto-occipital head circumference, which was determined by our team using standardized tape measurement in *N* = 23 3q29Del participants. Consistent with previous literature (Hshieh et al., 2016; Wolf et al., 2003 and others), these variables showed a significant positive correlation with eICV in our 3q29Del dataset, with moderate to strong correlation coefficients (p-values ≤ 0.05), providing an indirect metric for quality control. Note that visual inspection of the alignments from which eICV values were calculated did not reveal any errors. *Abbreviations*: 3q29 deletion syndrome, 3q29Del.

Buckner RL, Head D, Parker J, Fotenos AF, Marcus D, Morris JC, et al. (2004): A unified approach for morphometric and functional data analysis in young, old, and demented adults using automated atlas-based head size normalization: reliability and validation against manual measurement of total intracranial volume. *Neuroimage*. 23:724-738.

Hshieh TT, Fox ML, Kosar CM, Cavallari M, Guttmann CR, Alsop D, et al. (2016): Head circumference as a useful surrogate for intracranial volume in older adults. *Int Psychogeriatr*. 28:157-162.

Wolf H, Kruggel F, Hensel A, Wahlund LO, Arendt T, Gertz HJ (2003): The relationship between head size and intracranial volume in elderly subjects. *Brain Res*. 973:74-80.

**A**

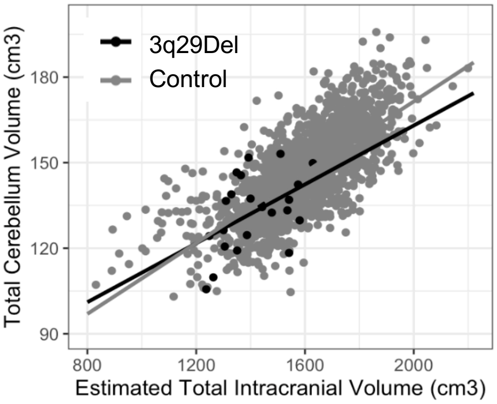

$$\mathbf{3q29Del} \mid \widehat{Total\ Cerebellum\ Volume} = 82.77^{***} - 0.31\,(Age) + 9.68\,(Sex_M)^{\dagger} + 0.03\,(eICV)$$

$$\mathbf{Control} \mid \widehat{Total\ Cerebellum\ Volume} = 60.15^{***} - 0.04\,(Age) + 5.31\,(Sex_M)^{***} + 0.05\,(eICV)^{***}$$

**Pooled analysis in 3q29Del & control participants:**
**eICV X Diagnostic group interaction effect:** *b* = -0.01, 95% CI = -0.04 – 0.03, p-value = 0.67

**B**

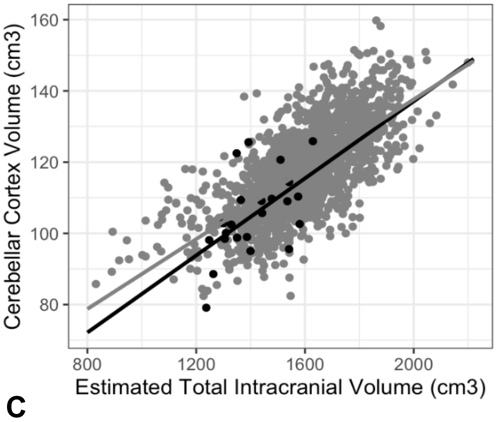

$$\mathbf{3q29Del} \mid \widehat{Cerebellar\ Cortex\ Volume} = 48.77 - 0.31\,(Age) + 5.99\,(Sex_M) + 0.04\,(eICV)^{*}$$

$$\mathbf{Control} \mid \widehat{Cerebellar\ Cortex\ Volume} = 55.61^{***} - 0.21\,(Age)^{***} + 4.75\,(Sex_M)^{***} + 0.04\,(eICV)^{***}$$

**Pooled analysis in 3q29Del & control participants:**
**eICV X Diagnostic group interaction effect:** *b* = 0.004, 95% CI = -0.03 – 0.03, p-value = 0.80

**C**

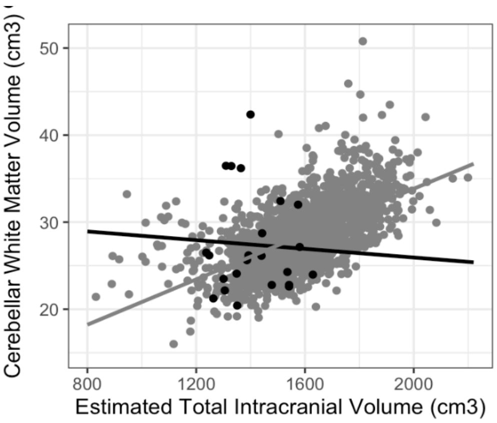

$$\mathbf{3q29Del} \mid \widehat{Cerebellar\ WM\ Volume} = 34.01 + 0.001\,(Age) + 3.69\,(Sex_M) - 0.01\,(eICV)$$

$$\mathbf{Control} \mid \widehat{Cerebellar\ WM\ Volume} = 4.53^{***} + 0.16\,(Age)^{***} + 0.56\,(Sex_M)^{**} + 0.01\,(eICV)^{***}$$

**Pooled analysis in 3q29Del & control participants:**
**eICV X Diagnostic group interaction effect:** *b* = -0.01, 95% CI = -0.02 – -0.001, p-value = 0.03*

**Fig. S3. Relationships between eICV and A) total cerebellum, B) cerebellar cortex, and C) cerebellar white matter volumes among 3q29Del participants versus controls.** The grey and black lines represent the estimated linear regression lines for control and 3q29Del groups, respectively. Corresponding multiple linear regression equations for each outcome, with age and sex added as covariates, are provided on the right. Four out of six group-specific intercepts were significantly different from zero (*p's* ≤ 0.05), indicating that the data do not meet the assumptions for using the "proportion method" for head size adjustment. When 3q29Del and control data were pooled, a significant interaction effect was identified between eICV and diagnostic group on cerebellar white matter volumes, while correcting for age and sex (*p* ≤ 0.05), with a significant positive slope observed among controls (*b* = 0.01, *p* ≤ 0.05) and a negative but not significant slope observed among 3q29Del participants (*b* = -0.01, *p* > 0.05). These inhomogeneous regression slopes, along with non-zero intercept tests support the implementation of the "residual method" as a more statistically appropriate approach to the eICV-adjustment of cerebellar volumes in downstream analyses. Refer to Mathalon et al. (1993), O'Brien et al. (2011) and Voevodskaya et al. (2014) for details on statistical considerations for head size adjustment. 3q29Del *N* = 23, Control *N* = 1,608. Contrast coding: reference level for the sex variable in regression models is female. p-value ≤ 0.001 '***', p-value ≤ 0.01 '**', p-value ≤ 0.05 '*', p-value ≤ 0.1 *'†'*. *Abbreviations*: 3q29 deletion syndrome, 3q29Del; unstandardized coefficient estimate, *b*; confidence interval, CI; white matter, WM; estimated total intracranial volume, eICV; male, M.

**A**

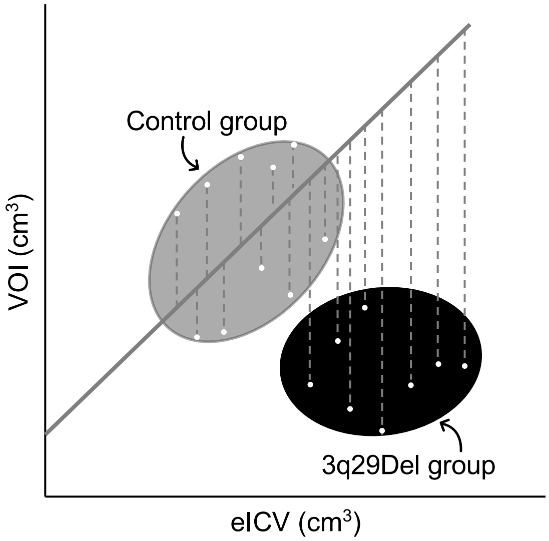

**B**

Using $i$ to indicate a given participant, obtain parameter estimates using the control data only:

$$Estimated(VOI_i)_{control} = b_{0\,control} + b_{1_{control}}(eICV_i) + b_{2_{control}}(Sex_i) + b_{3_{control}}(Age_i).$$

Generate residuals using the estimates from this model for all participants and calculate eICV-adjusted VOIs:

$$Adjusted(VOI_i) = VOI_i - b_{control} \times (eICV_i - \overline{eICV}_{control})$$

; where $\overline{eICV}_{control}$ is the mean $eICV$ of the control group.

$$Adjusted(VOI_i) = \underbrace{VOI_i - (b_{1_{control}} \times eICV_i)}_{residual_i} + \underbrace{(b_{control} \times \overline{eICV}_{control})}_{\overline{VOI}_{control}},$$

$$Adjusted(VOI_i) = residual_i + \overline{VOI}_{control}.$$

**Fig. S4. Correction of cerebellar volumes for head size variation in volumetric case-control analyses.** Absolute cerebellar volumes were adjusted for eICV using the residual method for head size correction (statistical justification for methodology is outlined in Fig. S3). **A)** A schematic illustration of the residual method for calculating eICV-adjusted VOIs, modified from O'Brien et al. (2011). The gray and black ellipses denote illustrative scatterplots for control and 3q29Del groups, respectively. The solid gray line represents a least-square derived linear regression between eICV and a given VOI, computed based on data from the control group only. Dashed vertical lines illustrate a sample of residuals calculated from this regression line for both control and 3q29Del participants. **B)** Formulas used for calculating eICV-adjusted VOIs based on the residual approach (disregarding error). We assume that the regression slope, *b1* represents the normative relationship between eICV and cerebellar volumes and that this relationship is not necessarily sustained in the 3q29Del group. Using the residual approach, we regress the VOI of healthy controls on the eICV of healthy controls, including age and sex as covariates, and use the estimates obtained from this linear regression model to calculate residuals for all participants from both diagnostic groups. Hence, each residual represents the deviation of a given participant's observed VOI from what would be expected of a control participant with the same eICV. Previous work has shown that the residual method is generally robust to systematic and random errors in MRI datasets (Sanfilipo et al., 2004), which provides advantages for detecting true group differences in combined datasets. Refer to Mathalon et al. (1993), O'Brien et al. (2011) and Voevodskaya et al. (2014) for details on statistical considerations for head size adjustment. *Abbreviations*: 3q29 deletion syndrome, 3q29Del; estimated total intracranial volume, eICV; volumetric measure of interest, VOI.

Mathalon DH, Sullivan EV, Rawles JM, Pfefferbaum A (1993): Correction for head size in brain-imaging measurements. *Psychiatry Res*. 50:121-139.

O'Brien LM, Ziegler DA, Deutsch CK, Frazier JA, Herbert MR, Locascio JJ (2011): Statistical adjustments for brain size in volumetric neuroimaging studies: some practical implications in methods. *Psychiatry Res*. 193:113-122.

Sanfilipo MP, Benedict RH, Zivadinov R, Bakshi R (2004): Correction for intracranial volume in analysis of whole brain atrophy in multiple sclerosis: the proportion vs. residual method. *Neuroimage*. 22:1732-1743.

Voevodskaya O, Simmons A, Nordenskjold R, Kullberg J, Ahlstrom H, Lind L, et al. (2014): The effects of intracranial volume adjustment approaches on multiple regional MRI volumes in healthy aging and Alzheimer's disease. *Front Aging Neurosci*. 6:264.

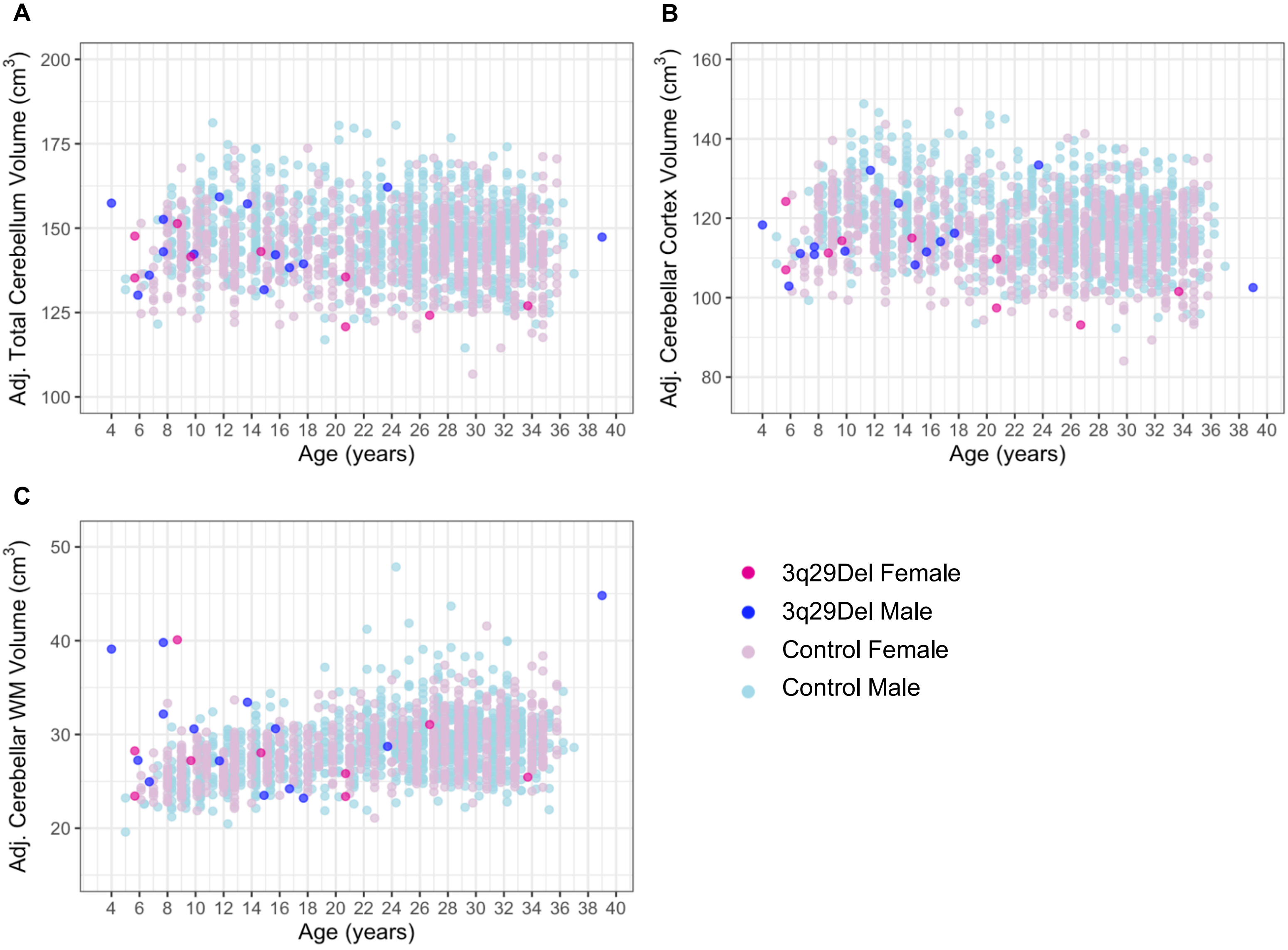

**Fig. S5. Scatter plots showing the distribution of eICV-adjusted A) total cerebellum volume, B) cerebellar cortex volume, and C) cerebellar white matter volume as a function of age among male and female participants in each diagnostic group.** A slight jitter was added systematically to all panels to minimize overplotting. Control *N* = 1,608 (Female *N* = 861, Male *N* = 747), 3q29Del *N* = 23 (Female *N* = 9, Male *N* = 14). *Abbreviations*: 3q29 deletion syndrome, 3q29Del; adjusted for estimated total intracranial volume, adj.; white matter, WM.

**Table S3. Extended linear regression results testing the effect of diagnostic group on volumetric measures of interest and polynomial modeling of age. A-H)** Multiple linear regression models include sex, age, age$^2$ and/or age$^3$ as covariates. The main effect of diagnostic group is reported in bold for clarity. ANOVAs were performed to sequentially compare simpler models to more complex models to identify the best-fitting polynomial function of age for each VOI (highlighted in blue). The relationship between the best-fitting polynomial function of age and VOIs is plotted in the right bottom corner of each panel using data pooled across diagnostic groups. Final inferences are based on heteroskedasticity-robust estimates, which are provided above non-robust OLS estimates (in grey brackets), along with robust Wald statistics, for the best-fitting models only. We also report exact p-values calculated by non-asymptotic permutation marginal tests. Contrast coding: reference levels for the diagnostic group and sex variables are healthy control and female, respectively.

#### A. VOI: Absolute Total Cerebellum Volume (cm$^3$)

| Degree of polynomial | Explanatory variables | *b* | CI (95%) | p-value | perm. p-value |
|---|---|---|---|---|---|
| **Linear (Model 1)** | Intercept | 140.44 | 138.52 – 142.36 | < 2.00E-16*** | |
| | Age (years) | -0.10 | -0.17 – -0.02 | 0.01** | 0.01** |
| | Sex [Male] | 16.50 | 15.27 – 17.74 | < 2.00E-16*** | 1.00E-04*** |
| | **Diagnostic Group [3q29Del]** | **-16.25** | **-21.50 – -11.00** | **1.57E-09*** ** | **1.00E-04*** ** |
| | $R^2$ / $R^2$ adjusted | 0.31 / 0.31 | | | |
| | F-statistic (OLS) | 241.7 on 3 and 1627 DF, p-value < 2.20E-16*** | | | |
| **Quadratic (Model 2)**<br>*Best-fit* | Intercept | 133.50 | 128.97 – 138.03<br>[129.05 – 137.96] | < 2.00E-16***<br>[< 2.00E-16***] | |
| | Age (years) | 0.68 | 0.22 – 1.14<br>[0.22 – 1.13] | 3.72E-03**<br>[3.63E-03**] | 3.30E-03*** |
| | Age$^2$ | -0.02 | -0.03 – -0.01<br>[-0.03 – -0.01] | 6.22E-04***<br>[7.31E-04***] | 1.00E-03*** |
| | Sex [Male] | 16.22 | 14.97 – 17.48<br>[14.98 – 17.47] | < 2.00E-16***<br>[< 2.00E-16***] | 1.00E-04*** |
| | **Diagnostic Group [3q29Del]** | **-15.26** | **-20.01 – -10.51**<br>**[-20.53 – -10.00]** | **3.76E-10*** **<br>**[1.53E-08***]** | **1.00E-04*** ** |
| | $R^2$ / $R^2$ adjusted | 0.31 / 0.31 | | | |
| | Robust Wald test | 184.0 on 4 and 1626 DF, p-value < 2.20E-16*** | | | |
| | [F-statistic (OLS) | 185.3 on 4 and 1626 DF, p-value < 2.20E-16***] | | | |
| **Cubic (Model 3)** | Intercept | 127.70 | 117.23 – 138.16 | < 2.00E-16*** | |
| | Age (years) | 1.69 | -0.02 – 3.39 | 0.06† | 0.06† |
| | Age$^2$ | -0.07 | -0.15 – 0.01 | 0.11 | 0.11 |
| | Age$^3$ | 0.001 | -0.001 – 0.002 | 0.23 | 0.23 |
| | Sex [Male] | 16.25 | 15.01 – 17.49 | < 2.00E-16*** | 1.00E-04*** |
| | **Diagnostic Group [3q29Del]** | **-15.10** | **-20.37 – -9.83** | **2.25E-08*** ** | **1.00E-04*** ** |
| | $R^2$ / $R^2$ adjusted | 0.31 / 0.31 | | | |
| | F-statistic (OLS) | 148.6 on 5 and 1625 DF, p-value < 2.20E-16*** | | | |

Model 1 vs Model 2 – ANOVA Table:

| | Resid. DF | Resid. SS | DF | SS | F-value | Pr(>F) |
|---|---|---|---|---|---|---|
| **1** | 1627 | 260917 | | | | |
| **2** | 1626 | 259092 | 1 | 1824.90 | 11.45 | 7.31E-04*** |

Model 2 vs Model 3 – ANOVA Table:

| | Resid. DF | Resid. SS | DF | SS | F-value | Pr(>F) |
|---|---|---|---|---|---|---|
| **2** | 1626 | 259092 | | | | |
| **3** | 1625 | 258862 | 1 | 230.30 | 1.45 | 0.23 |

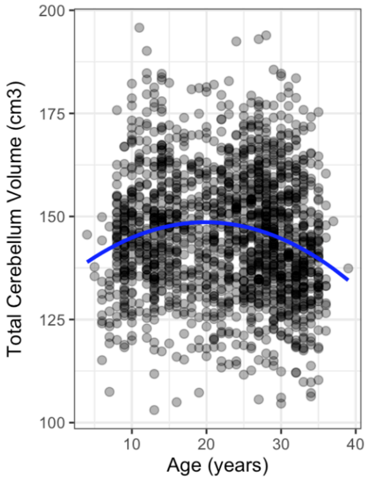

#### B. VOI: Absolute Cerebellar Cortex Volume ($cm^3$)

| Degree of polynomial | Explanatory variables | *b* | CI (95%) | p-value | perm. p-value |
|---|---|---|---|---|---|
| **Linear (Model 1)** | Intercept | 116.86 | 115.29 – 118.43 | < 2.00E-16*** | |
| | Age (years) | -0.25 | -0.31 – -0.19 | 3.87E-15*** | 1.00E-04*** |
| | Sex [Male] | 13.27 | 12.25 – 14.28 | < 2.00E-16*** | 1.00E-04*** |
| | **Diagnostic Group [3q29Del]** | **-15.85** | **-20.15 – -11.55** | **7.27E-13***** | **1.00E-04***** |
| | $R^2$ / $R^2$ adjusted | 0.32 / 0.32 | | | |
| | F-statistic (OLS) | 257.9 on 3 and 1627 DF, p-value < 2.20E-16*** | | | |
| **Quadratic (Model 2)** *Best-fit* | Intercept | 113.52 | 109.73 – 117.31 [109.86 – 117.18] | < 2.00E-16*** [< 2.00E-16***] | |
| | Age (years) | 0.13 | -0.26 – 0.51 [-0.25 – 0.50] | 0.52 [0.51] | 0.50 |
| | $Age^2$ | -0.01 | -0.02 – -0.0001 [-0.02 – -0.0001] | 0.04* [0.04*] | 0.04* |
| | Sex [Male] | 13.13 | 12.10 – 14.17 [12.11 – 14.15] | < 2.00E-16*** [< 2.00E-16***] | 1.00E-04*** |
| | **Diagnostic Group [3q29Del]** | **-15.38** | **-19.53 – -11.22 [-19.70 – -11.06]** | **5.86E-13*** [4.28E-12***]** | **1.00E-04***** |
| | $R^2$ / $R^2$ adjusted | 0.32 / 0.32 | | | |
| | Robust Wald test | 193.2 on 4 and 1626 DF, p-value < 2.20E-16*** | | | |
| | [F-statistic (OLS) | 194.8 on 4 and 1626 DF, p-value < 2.20E-16***] | | | |
| **Cubic (Model 3)** | Intercept | 107.48 | 98.89 – 116.07 | < 2.00E-16 *** | |
| | Age (years) | 1.18 | -0.23 – 2.58 | 0.10† | 0.10† |
| | $Age^2$ | -0.06 | -0.13 – 0.01 | 0.08† | 0.08† |
| | $Age^3$ | 0.001 | -0.0002 – 0.002 | 0.13 | 0.13 |
| | Sex [Male] | 13.16 | 12.14 – 14.17 | < 2.00E-16 *** | 1.00E-04 *** |
| | **Diagnostic Group [3q29Del]** | **-15.21** | **-19.53 – -10.88** | **7.57E-12 ***** | **1.00E-04 ***** |
| | $R^2$ / $R^2$ adjusted | 0.32 / 0.32 | | | |
| | F-statistic (OLS) | 156.4 on 5 and 1625 DF, p-value < 2.20E-16 *** | | | |

Model 1 vs Model 2 – ANOVA Table:

| | Resid. DF | Resid. SS | DF | SS | F-value | Pr(>F) |
|---|---|---|---|---|---|---|
| **1** | 1627 | 174977 | | | | |
| **2** | 1626 | 174554 | 1 | 422.96 | 3.94 | 0.04 * |

Model 2 vs Model 3 – ANOVA Table:

| | Resid. DF | Resid. SS | DF | SS | F-value | Pr(>F) |
|---|---|---|---|---|---|---|
| **2** | 1626 | 174554 | | | | |
| **3** | 1625 | 174305 | 1 | 249.44 | 2.33 | 0.13 |

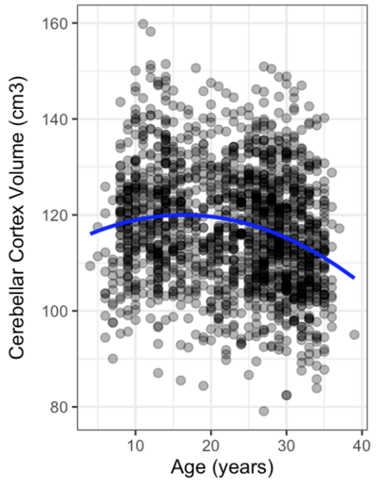

#### C. VOI: Absolute Cerebellar White Matter Volume ($cm^3$)

| Degree of polynomial | Explanatory variables | *b* | CI (95%) | p-value | perm. p-value |
|---|---|---|---|---|---|
| **Linear (Model 1)** | Intercept | 23.58 | 23.07 – 24.10 | < 2.00E-16*** | |
| | Age (years) | 0.15 | 0.13 – 0.17 | < 2.00E-16*** | 1.00E-04*** |
| | Sex [Male] | 3.24 | 2.90 – 3.57 | < 2.00E-16*** | 1.00E-04*** |
| | **Diagnostic Group [3q29Del]** | **-0.40** | **-1.80 – 1.01** | **0.58** | **0.57** |
| | $R^2$ / $R^2$ adjusted | 0.26 / 0.26 | | | |
| | F-statistic (OLS) | 187.3 on 3 and 1627 DF, p-value < 2.20E-16 *** | | | |
| **Quadratic (Model 2)** *Best-fit* | Intercept | 19.98 | 18.82 – 21.15 [18.80 – 21.16] | < 2.00E-16*** [< 2.00E-16***] | |
| | Age (years) | 0.55 | 0.43 – 0.67 [0.43 – 0.67] | < 2.00E-16*** [< 2.00E-16***] | 1.00E-04*** |
| | $Age^2$ | -0.01 | -0.01 – -0.01 [-0.01 – -0.01] | 8.88E-11*** [4.94E-11***] | 1.00E-04*** |
| | Sex [Male] | 3.09 | 2.76 – 3.43 [2.76 – 3.42] | < 2.00E-16*** [< 2.00E-16***] | 1.00E-04*** |
| | **Diagnostic Group [3q29Del]** | **0.11** | **-2.42 – 2.65** **[-1.28 – 1.51]** | **0.93** **[0.87]** | **0.87** |
| | $R^2$ / $R^2$ adjusted | 0.28 / 0.27 | | | |
| | Robust Wald test | 155.1 on 4 and 1626 DF, p-value < 2.20E-16*** | | | |
| | [F-statistic (OLS) | 155.1 on 4 and 1626 DF, p-value < 2.20E-16***] | | | |
| **Cubic (Model 3)** | Intercept | 20.22 | 17.45 – 23.00 | < 2.00E-16*** | |
| | Age (years) | 0.51 | 0.06 – 0.96 | 0.03* | 0.03* |
| | $Age^2$ | -0.01 | -0.03 – 0.02 | 0.52 | 0.53 |
| | $Age^3$ | 0.00 | -0.0004 – 0.0003 | 0.85 | 0.85 |
| | Sex [Male] | 3.09 | 2.76 – 3.42 | < 2.00E-16*** | 1.00E-04*** |
| | **Diagnostic Group [3q29Del]** | **0.11** | **-1.29 – 1.51** | **0.88** | **0.87** |
| | $R^2$ / $R^2$ adjusted | 0.28 / 0.27 | | | |
| | F-statistic (OLS) | 124.0 on 5 and 1625 DF, p-value < 2.20E-16 *** | | | |

Model 1 vs Model 2 – ANOVA Table:

| | Resid. DF | Resid. SS | DF | SS | F-value | Pr(>F) |
|---|---|---|---|---|---|---|
| **1** | 1627 | 18695 | | | | |
| **2** | 1626 | 18205 | 1 | 490.30 | 43.79 | 4.94E-11*** |

Model 2 vs Model 3 – ANOVA Table:

| | Resid. DF | Resid. SS | DF | SS | F-value | Pr(>F) |
|---|---|---|---|---|---|---|
| **2** | 1626 | 18205 | | | | |
| **3** | 1625 | 18204 | 1 | 0.39 | 0.04 | 0.85 |

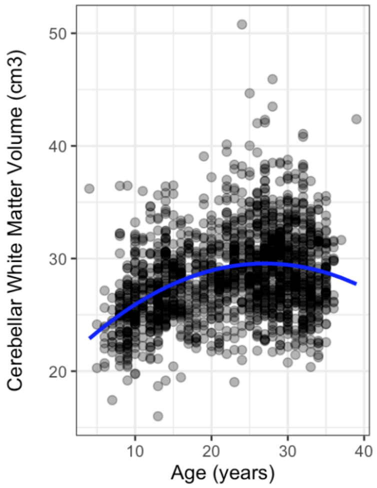

#### D. VOI: Cerebellar Cortex to Cerebellar White Matter Volume Ratio

| Degree of polynomial | Explanatory variables | *b* | CI (95%) | p-value | perm. p-value |
|---|---|---|---|---|---|
| **Linear (Model 1)** | Intercept | 4.88 | 4.82 – 4.94 | < 2.00E-16*** | |
| | Age (years) | -0.03 | -0.03 – -0.03 | < 2.00E-16*** | 1.00E-04*** |
| | Sex [Male] | 0.01 | -0.03 – 0.05 | 0.56 | 0.56 |
| | **Diagnostic Group [3q29Del]** | **-0.42** | **-0.58 – -0.25** | **5.35E-07*** ** | **1.00E-04*** ** |
| | $R^2$ / $R^2$ adjusted | 0.31 / 0.31 | | | |
| | F-statistic (OLS) | 240.4 on 3 and 1627 DF, p-value < 2.20E-16*** | | | |
| **Quadratic (Model 2)** *Best-fit* | Intercept | 5.38 | 5.22 – 5.53 [5.22 – 5.53] | < 2.00E-16*** [< 2.00E-16***] | |
| | Age (years) | -0.09 | -0.10 – -0.07 [-0.10 – -0.07] | < 2.00E-16*** [< 2.00E-16***] | 1.00E-04*** |
| | $Age^2$ | 0.001 | 0.001 – 0.002 [0.001 – 0.002] | 9.59E-13*** [2.46E-15***] | 1.00E-04*** |
| | Sex [Male] | 0.03 | -0.01 – 0.07 [-0.01 – 0.07] | 0.11 [0.11] | 0.11 |
| | **Diagnostic Group [3q29Del]** | **-0.49** | **-0.86 – -0.12 [-0.65 – -0.33]** | **9.82E-03** [2.92E-09***]** | **1.00E-04*** ** |
| | $R^2$ / $R^2$ adjusted | 0.33 / 0.33 | | | |
| | Robust Wald test | 179.4 on 4 and 1626 DF, p-value < 2.20E-16*** | | | |
| | [F-statistic (OLS) | 203.2 on 4 and 1626 DF, p-value < 2.20E-16***] | | | |
| **Cubic (Model 3)** | Intercept | 5.30 | 4.98 – 5.62 | < 2.00E-16*** | |
| | Age (years) | -0.07 | -0.13 – -0.02 | 0.01** | 0.01** |
| | $Age^2$ | 0.001 | -0.002 – 0.003 | 0.63 | 0.63 |
| | $Age^3$ | 0.00 | 0.00 – 0.0001 | 0.60 | 0.59 |
| | Sex [Male] | 0.03 | -0.01 – 0.07 | 0.10† | 0.11 |
| | **Diagnostic Group [3q29Del]** | **-0.49** | **-0.65 – -0.33** | **3.61E-09*** ** | **1.00E-04*** ** |
| | $R^2$ / $R^2$ adjusted | 0.33 / 0.33 | | | |
| | F-statistic (OLS) | 162.6 on 5 and 1625 DF, p-value < 2.2E-16 *** | | | |

Model 1 vs Model 2 – ANOVA Table:

| | Resid. DF | Resid. SS | DF | SS | F-value | Pr(>F) |
|---|---|---|---|---|---|---|
| **1** | 1627 | 250.16 | | | | |
| **2** | 1626 | 240.70 | 1 | 9.46 | 63.90 | 2.46E-15*** |

Model 2 vs Model 3 – ANOVA Table:

| | Resid. DF | Resid. SS | DF | SS | F-value | Pr(>F) |
|---|---|---|---|---|---|---|
| **2** | 1626 | 240.70 | | | | |
| **3** | 1625 | 240.66 | 1 | 0.04 | 0.27 | 0.60 |

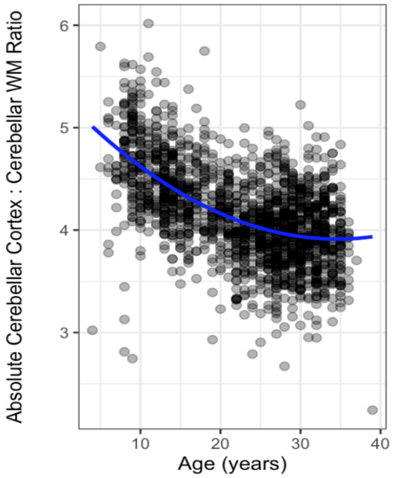

#### E. VOI: Estimated Total Intracranial Volume (eICV) (cm3)

| Degree of polynomial | Explanatory variables | *b* | CI (95%) | p-value | perm. p-value |
|---|---|---|---|---|---|
| **Linear (Model 1)** | Intercept | 1511.99 | 1490.34 – 1533.63 | < 2.00E-16*** | |
| | Age (years) | -0.97 | -1.81 – -0.12 | 0.03* | 0.02* |
| | Sex [Male] | 209.85 | 195.94 – 223.77 | < 2.00E-16*** | 1.00E-04*** |
| | **Diagnostic Group [3q29Del]** | **-211.79** | **-270.97 – -152.61** | **3.26E-12*** ** | **1.00E-04*** ** |
| | $R^2$ / $R^2$ adjusted | 0.36 / 0.36 | | | |
| | F-statistic (OLS) | 306.8 on 3 and 1627 DF, p-value < 2.20E-16*** | | | |
| **Quadratic (Model 2)** *Best-fit* | Intercept | 1415.02 | 1367.87 – 1462.17 [1364.89 – 1465.14] | < 2.00E-16*** [< 2.00E-16***] | |
| | Age (years) | 9.89 | 4.84 – 14.94 [4.75 – 15.03] | 1.26E-04*** [1.65E-04***] | 2.00E-04*** |
| | $Age^2$ | -0.25 | -0.37 – -0.13 [-0.37 – -0.14] | 3.66E-05*** [2.79E-05***] | 1.00E-04*** |
| | Sex [Male] | 206.00 | 192.06 – 219.93 [192.03 – 219.96] | < 2.00E-16*** [< 2.00E-16***] | 1.00E-04*** |
| | **Diagnostic Group [3q29Del]** | **-197.99** | **-253.23 – -142.74 [-257.22 – -138.75]** | **3.05E-12*** [7.39E-11***]** | **1.00E-04*** ** |
| | $R^2$ / $R^2$ adjusted | 0.37 / 0.37 | | | |
| | Robust Wald test | 232.5 on 4 and 1626 DF, p-value < 2.20E-16*** | | | |
| | [F-statistic (OLS) | 236.9 on 4 and 1626 DF, p-value < 2.20E-16***] | | | |
| **Cubic (Model 3)** | Intercept | 1419.77 | 1301.95 – 1537.60 | < 2.00E-16*** | |
| | Age (years) | 9.06 | -10.14 – 28.27 | 0.36 | 0.36 |
| | $Age^2$ | -0.21 | -1.16 – 0.74 | 0.66 | 0.66 |
| | $Age^3$ | -0.001 | -0.02 – 0.01 | 0.93 | 0.93 |
| | Sex [Male] | 205.98 | 192.01 – 219.95 | < 2.00E-16*** | 1.00E-04*** |
| | **Diagnostic Group [3q29Del]** | **-198.12** | **-257.44 – -138.79** | **7.69E-11*** ** | **1.00E-04*** ** |
| | $R^2$ / $R^2$ adjusted | 0.37 / 0.37 | | | |
| | F-statistic (OLS) | 189.4 on 5 and 1625 DF, p-value < 2.20E-16*** | | | |

Model 1 vs Model 2 – ANOVA Table:

| | Resid. DF | Resid. SS | DF | SS | F-value | Pr(>F) |
|---|---|---|---|---|---|---|
| **1** | 1627 | 33159001 | | | | |
| **2** | 1626 | 32802768 | 1 | 356233 | 17.66 | 2.79E-05*** |

Model 2 vs Model 3 – ANOVA Table:

| | Resid. DF | Resid. SS | DF | SS | F-value | Pr(>F) |
|---|---|---|---|---|---|---|
| **2** | 1626 | 32802768 | | | | |
| **3** | 1625 | 32802613 | 1 | 154.45 | 0.01 | 0.93 |

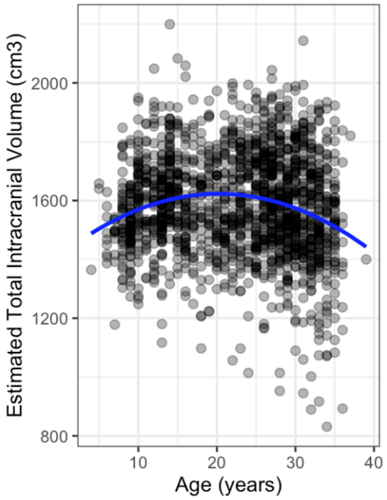

#### F. VOI: eICV-Adjusted Total Cerebellum Volume (cm$^3$)

| Degree of polynomial | Explanatory variables | *b* | CI (95%) | p-value | perm. p-value |
|---|---|---|---|---|---|
| **Linear (Model 1)** *Best-fit* | Intercept | 144.44 | 142.97 – 145.92<br>[142.90 – 145.99] | < 2.00E-16***<br>[< 2.00E-16***] | |
| | Age (years) | -0.05 | -0.11 – 0.01<br>[-0.11 – 0.01] | 0.12<br>[0.12] | 0.12 |
| | Sex [Male] | 5.38 | 4.38 – 6.38<br>[4.39 – 6.37] | < 2.00E-16***<br>[< 2.00E-16***] | 1.00E-04*** |
| | **Diagnostic Group [3q29Del]** | **-5.02** | **-9.25 – -0.80**<br>**[-9.24 – -0.81]** | **0.02***<br>**[0.02*]** | **0.02*** |
| | $R^2$ / $R^2$ adjusted | 0.07 / 0.07 | | | |
| | Robust Wald test | 39.0 on 3 and 1627 DF, p-value < 2.20E-16*** | | | |
| | [F-statistic (OLS) | 40.1 on 3 and 1627 DF, p-value < 2.20E-16***] | | | |
| **Quadratic (Model 2)** | Intercept | 142.64 | 139.06 – 146.23 | < 2.00E-16*** | |
| | Age (years) | 0.15 | -0.21 – 0.52 | 0.41 | 0.41 |
| | $Age^2$ | -0.005 | -0.01 – 0.004 | 0.28 | 0.27 |
| | Sex [Male] | 5.31 | 4.31 – 6.31 | < 2.00E-16*** | 1.00E-04*** |
| | **Diagnostic Group [3q29Del]** | **-4.77** | **-9.01 – -0.53** | **0.03*** | **0.03*** |
| | $R^2$ / $R^2$ adjusted | 0.07 / 0.07 | | | |
| | F-statistic (OLS) | 30.4 on 4 and 1626 DF, p-value < 2.20E-16*** | | | |
| **Cubic (Model 3)** | Intercept | 136.59 | 128.16 – 145.02 | < 2.00E-16*** | |
| | Age (years) | 1.21 | -0.17 – 2.58 | 0.09† | 0.08† |
| | $Age^2$ | -0.06 | -0.13 – 0.01 | 0.09† | 0.09† |
| | $Age^3$ | 0.001 | -0.0002 – 0.002 | 0.12 | 0.12 |
| | Sex [Male] | 5.33 | 4.33 – 6.33 | < 2.00E-16*** | 1.00E-04*** |
| | **Diagnostic Group [3q29Del]** | **-4.60** | **-8.84 – -0.35** | **0.03*** | **0.03*** |
| | $R^2$ / $R^2$ adjusted | 0.07 / 0.07 | | | |
| | F-statistic (OLS) | 24.8 on 5 and 1625 DF, p-value < 2.20E-16*** | | | |

Model 1 vs Model 2 – ANOVA Table:

| | Resid. DF | Resid. SS | DF | SS | F-value | Pr(>F) |
|---|---|---|---|---|---|---|
| **1** | 1627 | 168327 | | | | |
| **2** | 1626 | 168204 | 1 | 122.66 | 1.19 | 0.28 |

Model 2 vs Model 3 – ANOVA Table:

| | Resid. DF | Resid. SS | DF | SS | F-value | Pr(>F) |
|---|---|---|---|---|---|---|
| **2** | 1626 | 168204 | | | | |
| **3** | 1625 | 167953 | 1 | 250.74 | 2.43 | 0.12 |

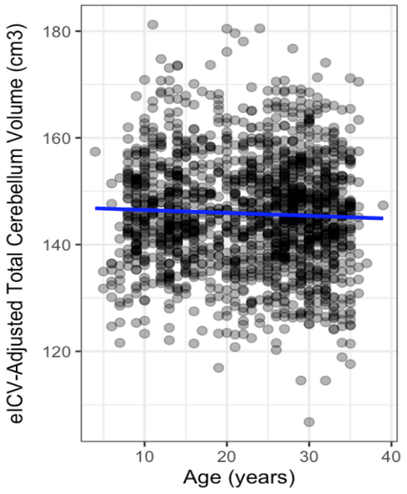

### G. VOI: eICV-Adjusted Cerebellar Cortex Volume ($cm^3$)

| Degree of polynomial | Explanatory variables | *b* | CI (95%) | p-value | perm. p-value |
|---|---|---|---|---|---|
| **Linear (Model 1)** *Best-fit* | Intercept | 119.88 | 118.60 – 121.17 [118.58 – 121.19] | < 2.00E-16*** [< 2.00E-16***] | |
| | Age (years) | -0.21 | -0.26 – -0.16 [-0.26 – -0.16] | 1.37E-15*** [1.42E-15***] | 1.00E-04*** |
| | Sex [Male] | 4.87 | 4.03 – 5.72 [4.03 – 5.71] | < 2.00E-16*** [< 2.00E-16***] | 1.00E-04*** |
| | **Diagnostic Group [3q29Del]** | **-7.38** | **-10.98 – -3.78** **[-10.95 – -3.81]** | **6.03E-05*** [5.28E-05***]** | **1.00E-04*** |
| | $R^2$ / $R^2$ adjusted | 0.11 / 0.11 | | | |
| | Robust Wald test | 67.5 on 3 and 1627 DF, p-value < 2.20E-16*** | | | |
| | [F-statistic (OLS) | 70.1 on 3 and 1627 DF, p-value < 2.20E-16***] | | | |
| **Quadratic (Model 2)** | Intercept | 120.42 | 117.38 – 123.46 | < 2.00E-16*** | |
| | Age (years) | -0.27 | -0.58 – 0.04 | 0.09† | 0.09† |
| | $Age^2$ | 0.001 | -0.01 – 0.01 | 0.70 | 0.70 |
| | Sex [Male] | 4.89 | 4.05 – 5.74 | < 2.00E-16*** | 1.00E-04*** |
| | **Diagnostic Group [3q29Del]** | **-7.46** | **-11.05 – -3.86** | **4.92E-05*** | **2.00E-04*** |
| | $R^2$ / $R^2$ adjusted | 0.11 / 0.11 | | | |
| | F-statistic (OLS) | 52.6 on 4 and 1626 DF, p-value < 2.20E-16 *** | | | |
| **Cubic (Model 3)** | Intercept | 114.19 | 107.05 – 121.33 | < 2.00E-16*** | |
| | Age (years) | 0.81 | -0.35 – 1.98 | 0.17 | 0.17 |
| | $Age^2$ | -0.05 | -0.11 – 0.00 | 0.07† | 0.07† |
| | $Age^3$ | 0.001 | 0.00 – 0.002 | 0.06† | 0.06† |
| | Sex [Male] | 4.92 | 4.07 – 5.76 | < 2.00E-16*** | 1.00E-04*** |
| | **Diagnostic Group [3q29Del]** | **-7.28** | **-10.88 – -3.69** | **7.40E-05*** | **3.00E-04*** |
| | $R^2$ / $R^2$ adjusted | 0.12 / 0.11 | | | |
| | F-statistic (OLS) | 2442.8 on 5 and 1625 DF, p-value < 2.20E-16*** | | | |

Model 1 vs Model 2 – ANOVA Table:

| | Resid. DF | Resid. SS | DF | SS | F-value | Pr(>F) |
|---|---|---|---|---|---|---|
| **1** | 1627 | 120694 | | | | |
| **2** | 1626 | 120683 | 1 | 10.95 | 0.15 | 0.70 |

Model 2 vs Model 3 – ANOVA Table:

| | Resid. DF | Resid. SS | DF | SS | F-value | Pr(>F) |
|---|---|---|---|---|---|---|
| **2** | 1626 | 120683 | | | | |
| **3** | 1625 | 120418 | 1 | 265.34 | 3.58 | 0.06† |

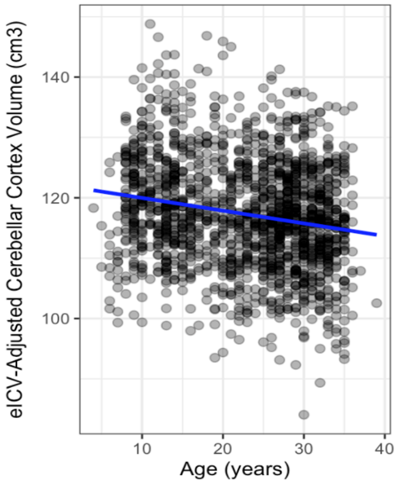

#### H. VOI: eICV-Adjusted Cerebellar White Matter Volume ($cm^3$)

| Degree of polynomial | Explanatory variables | *b* | CI (95%) | p-value | perm. p-value |
|---|---|---|---|---|---|
| **Linear (Model 1)** | Intercept | 24.56 | 24.12 – 25.00 | < 2.00E-16*** | |
| | Age (years) | 0.16 | 0.14 – 0.18 | < 2.00E-16*** | 1.00E-04*** |
| | Sex [Male] | 0.51 | 0.23 – 0.79 | 4.33E-04*** | 8.00E-04*** |
| | **Diagnostic Group [3q29Del]** | **2.36** | **1.16 – 3.55** | **1.22E-04*** ** | **3.00E-04*** ** |
| | $R^2$ / $R^2$ adjusted | 0.18 / 0.18 | | | |
| | F-statistic (OLS) | 118.2 on 3 and 1627 DF, p-value < 2.20E-16*** | | | |
| **Quadratic (Model 2)** *Best-fit* | Intercept | 22.22 | 21.22 – 23.23 [21.21 – 23.24] | < 2.00E-16*** [< 2.00E-16***] | |
| | Age (years) | 0.42 | 0.31 – 0.53 [0.32 – 0.53] | 6.39E-14*** [2.53E-15***] | 1.00E-04*** |
| | $Age^2$ | -0.01 | -0.01 – -0.004 [-0.01 – -0.004] | 3.94E-06*** [6.12E-07***] | 1.00E-04*** |
| | Sex [Male] | 0.41 | 0.13 – 0.70 [0.13 – 0.70] | 4.51E-03** [4.07E-03**] | 4.50E-03** |
| | **Diagnostic Group [3q29Del]** | **2.69** | **0.10 – 5.28 [1.49 – 3.89]** | **0.04* [1.15E-05***]** | **1.00E-04*** ** |
| | $R^2$ / $R^2$ adjusted | 0.19 / 0.19 | | | |
| | Robust Wald test | 106.7 on 4 and 1626 DF, p-value < 2.20E-16*** | | | |
| | [F-statistic (OLS) | 96.2 on 4 and 1626 DF, p-value < 2.20E-16***] | | | |
| **Cubic (Model 3)** | Intercept | 22.40 | 20.02 – 24.78 | < 2.00E-16 *** | |
| | Age (years) | 0.39 | 0.004 – 0.78 | 0.04* | 0.04* |
| | $Age^2$ | -0.005 | -0.02 – 0.01 | 0.64 | 0.63 |
| | $Age^3$ | 0.00 | -0.003 – 0.003 | 0.87 | 0.87 |
| | Sex [Male] | 0.41 | 0.13 – 0.70 | 4.16E-03** | 4.50E-03** |
| | **Diagnostic Group [3q29Del]** | **2.68** | **1.48 – 3.88** | **1.23E-05*** ** | **1.00E-04*** ** |
| | $R^2$ / $R^2$ adjusted | 0.19 / 0.19 | | | |
| | F-statistic (OLS) | 77.0 on 5 and 1625 DF, p-value < 2.20E-16*** | | | |

Model 1 vs Model 2 – ANOVA Table:

| | Resid. DF | Resid. SS | DF | SS | F-value | Pr(>F) |
|---|---|---|---|---|---|---|
| **1** | 1627 | 13626 | | | | |
| **2** | 1626 | 13420 | 1 | 206.93 | 25.07 | 6.12E-07*** |

Model 2 vs Model 3 – ANOVA Table:

| | Resid. DF | Resid. SS | DF | SS | F-value | Pr(>F) |
|---|---|---|---|---|---|---|
| **2** | 1626 | 13420 | | | | |
| **3** | 1625 | 13419 | 1 | 0.21 | 0.03 | 0.87 |

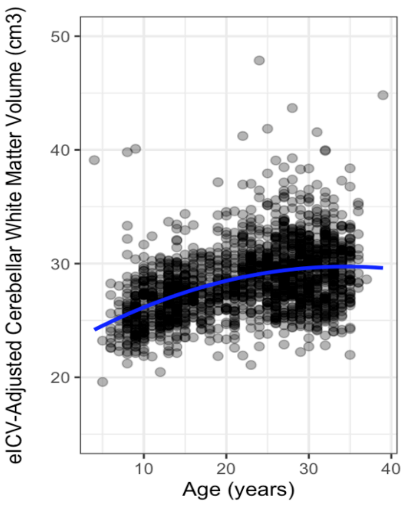

Control *N* = 1,608, 3q29Del *N* = 23. p-value ≤ 0.001 '***', p-value ≤ 0.01 '**', p-value ≤ 0.05 '*', p-value ≤ 0.1 '†'. *Abbreviations*: 3q29 deletion syndrome, 3q29Del; VOI, volumetric measure of interest; eICV, estimated total intracranial volume; unstandardized coefficient estimate, *b*; confidence interval, CI; degrees of freedom, DF; sum of squares, SS; analysis of variance, ANOVA; permutation, perm; ordinary least squares, OLS; residual, resid.

**Fig. S6. Regression diagnostics: testing the assumptions of ordinary least squares regression for best-fitting models from Table S3. A-H)** Plots of residuals versus predicted values for checking the assumptions of linearity and homoscedasticity in multiple linear regression. The residuals should appear approximately linear and be evenly spread around the red *y = 0* line (i.e., slope & y-intercept = 0) to assume a linear relationship between the predictors and the outcome variables and to assume homoscedasticity. **I-P)** QQ-plots of residuals for checking the normality assumption. The normal probability plot of residuals should approximately follow a 45-degree straight line to indicate that all errors are normally distributed around zero. Shapiro-Wilk normality tests were performed to additionally check that the residuals are normally distributed. **Q-X)** Scale-location plots for checking the homoscedasticity assumption. Standardized residuals reflect residuals divided by estimated standard error. Variance of the errors should be approximately the same for any combination of values of the independent variables. Studentized Breusch-Pagan tests were performed to additionally check for homoscedasticity. Given several observed violations of necessary linear regression assumptions, we calculated heteroscedasticity-robust estimates for final inferences. p-value ≤ 0.001 '***', p-value ≤ 0.01 '**', p-value ≤ 0.05 '*', p-value ≤ 0.1 '†'. *Abbreviations*: volumetric measure of interest, VOI; estimated total intracranial volume, eICV.

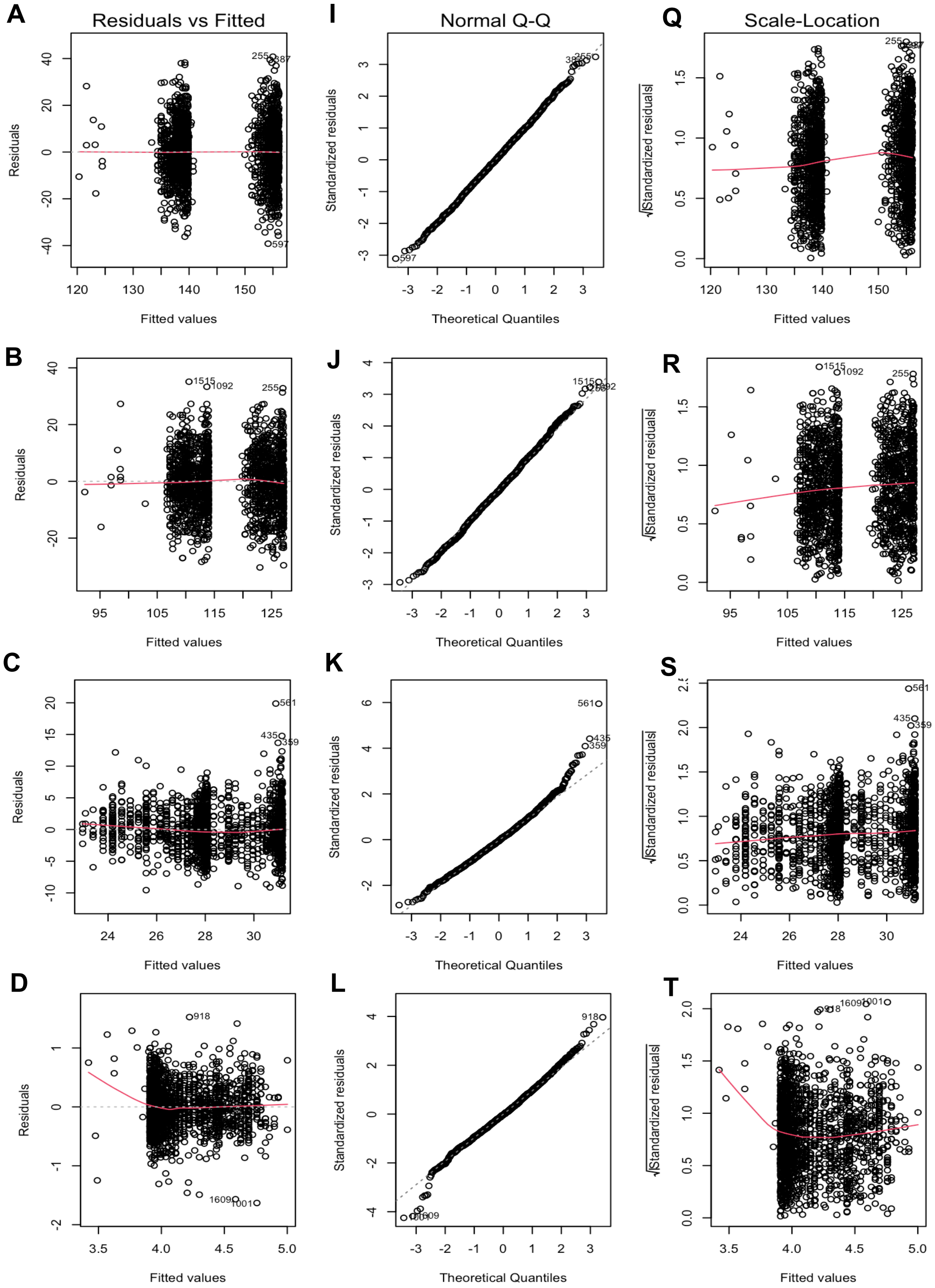

**VOI: Absolute Total Cerebellum Volume**
Best fit: Model 2 (refer to Table S3A)

Shapiro-Wilk test: W = 1.00, p-value = 0.80
Studentized Breusch-Pagan test: BP = 13.19, p-value = 0.01**.

**VOI: Absolute Cerebellar Cortex Volume**
Best fit: Model 2 (refer to Table S3B)

Shapiro-Wilk test: W = 1.00, p-value = 0.30
Studentized Breusch-Pagan test: BP = 11.05, p-value = 0.03*.

**VOI: Absolute Cerebellar White Matter Volume**
Best fit: Model 2 (refer to Table S3C)

Shapiro-Wilk test: W = 0.99, p-value = 2.15E-11***
Studentized Breusch-Pagan test: BP = 71.94, p-value = 8.85E-15***.

**VOI: Cerebellar Cortex to Cerebellar White Matter Volume Ratio**
Best fit: Model 2 (refer to Table S3D)

Shapiro-Wilk test: W = 0.99, p-value = 1.09E-06***
Studentized Breusch-Pagan test: BP = 180.9, p-value < 2.2E-16***.

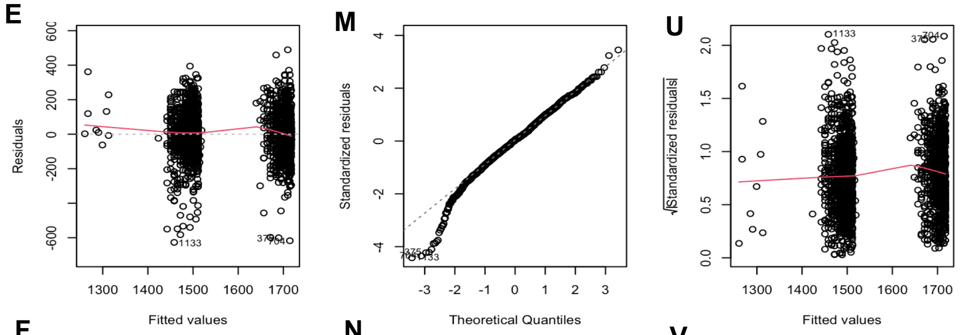

**VOI: eICV**
Best fit: Model 2 (refer to Table S3E)

Shapiro-Wilk test: W = 0.98, p-value = 1.66E-12***
Studentized Breusch-Pagan test: BP = 16.01, p-value = 3.00E-03**.

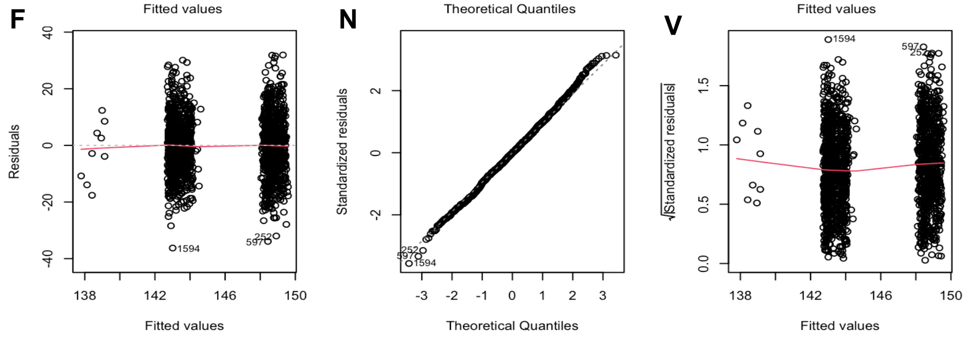

**VOI: eICV-Adjusted Total Cerebellum Volume**
Best fit: Model 1 (refer to Table S3F)

Shapiro-Wilk test: W = 1.00, p-value = 0.26
Studentized Breusch-Pagan test: BP = 12.86, p-value = 4.95E-03**.

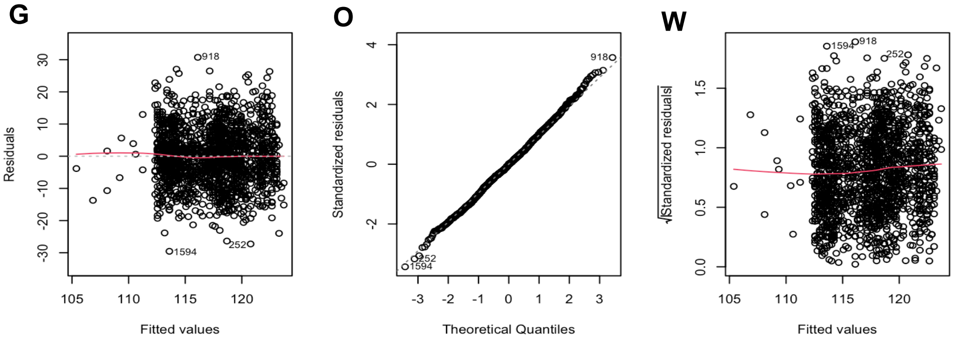

**VOI: eICV-Adjusted Cerebellar Cortex Volume**
Best fit: Model 1 (refer to Table S3G)

Shapiro-Wilk test: W = 1.00, p-value = 0.29
Studentized Breusch-Pagan test: BP = 6.67, p-value = 0.08$^{\dagger}$.

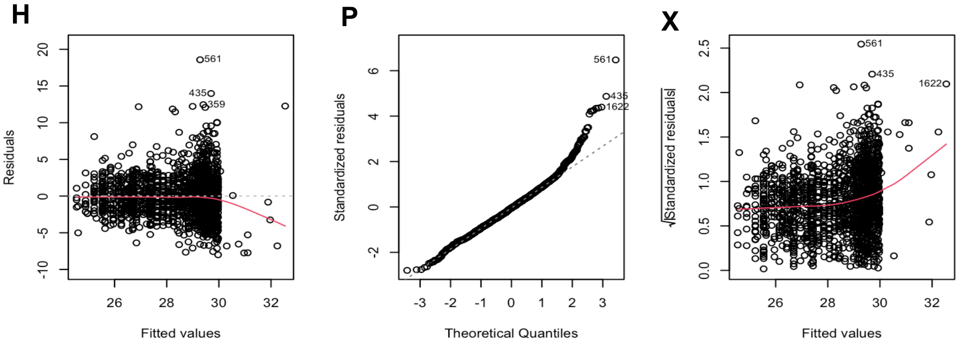

**VOI: eICV-Adjusted Cerebellar White Matter Volume**
Best fit: Model 2 (refer to Table S3H)

Shapiro-Wilk test: W = 0.97, p-value < 2.2E-16 ***
Studentized Breusch-Pagan test: BP = 121.71, p-value < 2.2E-16 ***.

| VOI | EDF | p-value | | |
|---|---|---|---|---|
| | | s(Age) (years) | Sex [Male] | Diagnostic Group [3q29Del] |
| **Total Cerebellum Volume ($cm^3$)** | | | | |
| **Absolute Volume** | 5.73 | 1.47E-06*** | < 2.00E-16*** (male > female) | **3.47E-07*** (3q29Del < control)** |
| **eICV-Adjusted Volume** | 4.35 | 0.09† | < 2.00E-16*** (male > female) | **0.05*** |
| **Cerebellar Cortex Volume ($cm^3$)** | | | | |
| **Absolute Volume** | 5.98 | < 2.00E-16*** | < 2.00E-16*** (male > female) | **5.32E-10*** (3q29Del < control)** |
| **eICV-Adjusted Volume** | 5.36 | < 2.00E-16*** | < 2.00E-16*** (male > female) | **4.67E-04*** (3q29Del < control)** |
| **Cerebellar White Matter Volume ($cm^3$)** | | | | |
| **Absolute Volume** | 5.26 | < 2.00E-16*** | < 2.00E-16*** (male > female) | **0.89** |
| **eICV-Adjusted Volume** | 3.99 | < 2.00E-16*** | 4.87E-03** (male > female) | **2.52E-05*** (3q29Del > control)** |
| **eICV ($cm^3$)** | 5.00 | 5.05E-06*** | < 2.00E-16*** (male > female) | **5.01E-10*** (3q29Del < control)** |

**Table S4. Summary of supplemental results from penalized cubic spline models testing the effect of diagnostic group on volumetric measures of interest.** To more flexibly account for linear and non-linear trajectories of volumetric change across age without requiring *a priori* selection of candidate models, we fit generalized additive models (GAM) with a cubic spline basis to our data as a supplemental method. Smoothing parameters were selected by the restricted maximum likelihood (REML) approach. EDF represents how complex the developmental pattern of the estimated volumetric trajectory is across age. EDF = 1 is equivalent to a straight line, EDF = 2 is equivalent to a quadratic curve, etc., with higher EDFs describing increased wiggliness. s(Age) represents the spline term of age. The main effect of diagnostic group is reported in bold for clarity. The p-values for sex and diagnostic group are derived from standard t-tests of whether the indicator of sex/diagnostic group are significant in determining each VOI. The p-values for s(Age) are derived from an approximate F-test of whether the smooth term of age is significant in the penalized cubic spline models for each VOI. Contrast coding: reference levels for the diagnostic group and sex variables are healthy control and female, respectively. For categorical variables, the direction of the corresponding effect in significant tests (*p's* ≤ 0.05) is specified by the ">" (greater than) or "<" (less than) symbols. Control *N* = 1,608**,** 3q29Del *N* = 23. p-value ≤ 0.001 '***', p-value ≤ 0.01 '**', p-value ≤ 0.05 '*', p-value ≤ 0.1 '†'. *Abbreviations*: 3q29 deletion syndrome, 3q29Del; estimated total intracranial volume, eICV; volumetric measure of interest, VOI; effective degrees of freedom, EDF.

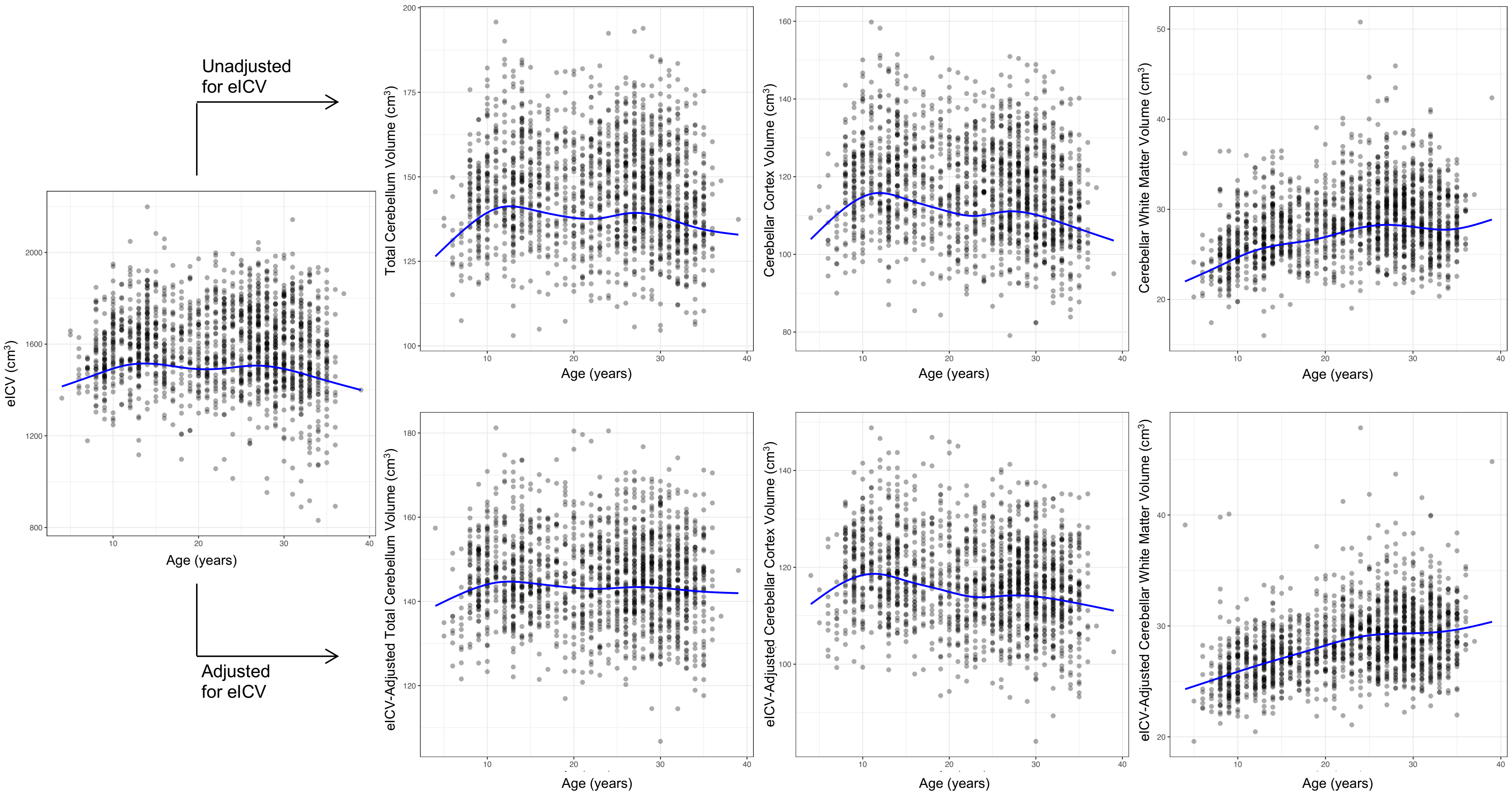

**Fig. S7. Developmental trajectories for volumetric measures of interest estimated by the penalized cubic spline approach.** Each dot represents volumetric data for a single study participant. Blue solid lines represent the estimated mean volumes. An indicator of sex (male / female), an indicator of diagnostic group (3q29Del / healthy control) and a spline term of age were included in each generalized additive model (GAM). Control *N* = 1,608**,** 3q29Del *N* = 23. *Abbreviations*: 3q29 deletion syndrome, 3q29Del; estimated total intracranial volume, eICV.

**Table S5. Exploratory modeling of diagnostic group by sex interaction effects on volumetric measures of interest. A-H)** Interrogated interaction effects are reported in bold for clarity. Covariates included in each linear regression model reflect the best-fitting models from Table S3. Final inferences are based on heteroskedasticity-robust estimates, which are provided above non-robust OLS estimates (in grey brackets), along with robust Wald statistics for each model. We also report exact p-values calculated by non-asymptotic permutation marginal tests. Contrast coding: reference levels for the diagnostic group and sex variables are healthy control and female, respectively.

| VOI | Explanatory variables | *b* | CI (95%) | p-value | perm. p-value |
|---|---|---|---|---|---|
| **A. Absolute Total Cerebellum Volume (cm$^3$)** | Intercept | 133.53 | 128.99 – 138.08<br>[129.08 – 137.99] | < 2.00E-16***<br>[< 2.00E-16***] | |
| | Age (years) | 0.67 | 0.21 – 1.13<br>[0.22 – 1.13] | 4.18E-03**<br>[3.96E-03**] | 3.10E-03** |
| | Age$^2$ | -0.02 | -0.03 – -0.01<br>[-0.03 – -0.01] | 7.13E-04***<br>[8.03E-04***] | 1.10E-03** |
| | Sex [Male] | 16.28 | 15.01 – 17.55<br>[15.03 – 17.53] | < 2.00E-16***<br>[< 2.00E-16***] | 1.00E-04*** |
| | Diagnostic Group [3q29Del] | -12.95 | -21.59 – -4.32<br>[-21.27 – -4.63] | 3.30E-03**<br>[2.29E-03**] | 2.20E-03** |
| | **Diagnostic Group x Sex** | **-3.82** | **-13.91 – 6.27**<br>**[-14.48 – 6.84]** | **0.46**<br>**[0.48]** | **0.48** |
| | $R^2$ / $R^2$ adjusted | 0.31 / 0.31 | | | |
| | Robust Wald test | 148.1 on 5 and 1625 DF, p-value < 2.20E-16*** | | | |
| | [F-statistic (OLS) | 148.3 on 5 and 1625 DF, p-value < 2.20E-16***] | | | |
| **B. Absolute Cerebellar Cortex Volume (cm$^3$)** | Intercept | 113.56 | 109.77 – 117.35<br>[109.90 – 117.22] | < 2.00E-16***<br>[< 2.00E-16***] | |
| | Age (years) | 0.12 | -0.26 – 0.50<br>[-0.26 – 0.49] | 0.54<br>[0.53] | 0.52 |
| | Age$^2$ | -0.01 | -0.02 – 0.0001<br>[-0.02 – 0.0001] | 0.06†<br>[0.06†] | 0.06† |
| | Sex [Male] | 13.20 | 12.16 – 14.24<br>[12.17 – 14.22] | < 2.00E-16***<br>[< 2.00E-16***] | 1.00E-04*** |
| | Diagnostic Group [3q29Del] | -12.59 | -19.84 – -5.34<br>[-19.42 – -5.76] | 6.71E-04***<br>[3.06E-04***] | 2.00E-04*** |
| | **Diagnostic Group x Sex** | **-4.61** | **-13.25 – 4.03**<br>**[-13.36 – 4.14]** | **0.30**<br>**[0.30]** | **0.30** |
| | $R^2$ / $R^2$ adjusted | 0.32 / 0.32 | | | |
| | Robust Wald test | 155.6 on 5 and 1625 DF, p-value < 2.20E-16 *** | | | |
| | [F-statistic (OLS) | 156.0 on 5 and 1625 DF, p-value < 2.20E-16 ***] | | | |
| **C. Absolute Cerebellar White Matter Volume (cm$^3$)** | Intercept | 19.98 | 18.82 – 21.14<br>[18.80 – 21.16] | < 2.00E-16***<br>[< 2.00E-16***] | |
| | Age (years) | 0.55 | 0.43 –0.67<br>[0.43 – 0.67] | < 2.00E-16***<br>[< 2.00E-16***] | 1.00E-04*** |
| | Age$^2$ | -0.01 | -0.01 – -0.01<br>[-0.01 – -0.01] | 5.63E-11***<br>[4.53E-11***] | 1.00E-04*** |
| | Sex [Male] | 3.08 | 2.75 – 3.41<br>[2.75 – 3.41] | < 2.00E-16***<br>[< 2.00E-16***] | 1.00E-04*** |
| | Diagnostic Group [3q29Del] | -0.36 | -3.85 – 3.13<br>[-2.57 – 1.84] | 0.84<br>[0.75] | 0.75 |
| | **Diagnostic Group x Sex** | **0.79** | **-4.17 – 5.74**<br>**[-2.04 – 3.61]** | **0.76**<br>**[0.58]** | **0.58** |
| | $R^2$ / $R^2$ adjusted | 0.28 / 0.27 | | | |
| | Robust Wald test | 121.5 on 5 and 1625 DF, p-value < 2.20E-16*** | | | |
| | [F-statistic (OLS) | 124.1 on 5 and 1625 DF, p-value < 2.20E-16***] | | | |

| | | | | | |
|---|---|---|---|---|---|
| **D. Cerebellar Cortex to Cerebellar White Matter Volume Ratio** | Intercept | 5.38 | 5.22 – 5.53<br>[5.24 – 5.51] | < 2.00E-16***<br>[< 2.00E-16***] | |
| | Age (years) | -0.09 | -0.10 – -0.07<br>[-0.10 – -0.07] | < 2.00E-16***<br>[< 2.00E-16***] | 1.00E-04*** |
| | $Age^2$ | 0.001 | 0.001 – 0.002<br>[0.001 – 0.002] | 5.16E-13***<br>[1.87E-15***] | 1.00E-04*** |
| | Sex [Male] | 0.03 | -0.004 – 0.07<br>[-0.003 – 0.07] | 0.07†<br>[0.08†] | 0.09† |
| | Diagnostic Group [3q29Del] | -0.37 | -0.86 – 0.11<br>[-0.63 – -0.12] | 0.13<br>[3.96E-03**] | 6.40E-03** |
| | **Diagnostic Group x Sex** | **-0.19** | **-0.91 – 0.52**<br>**[-0.52 – 0.13]** | **0.60**<br>**[0.25]** | **0.25** |
| | $R^2$ / $R^2$ adjusted | 0.33 / 0.33 | | | |
| | Robust Wald test | 144.8 on 5 and 1625 DF, p-value < 2.20E-16*** | | | |
| | [F-statistic (OLS) | 162.9 on 5 and 1625 DF, p-value < 2.20E-16***] | | | |
| **E. Estimated Total Intracranial Volume (eICV) ($cm^3$)** | Intercept | 1416.29 | 1369.41 – 1463.16]<br>[1366.23 – 1466.34] | < 2.00E-16***<br>[< 2.00E-16***] | |
| | Age (years) | 9.66 | 4.64 – 14.69<br>[4.53 – 14.79] | 1.69E-04***<br>[2.29E-04***] | 3.00E-04*** |
| | $Age^2$ | -0.25 | -0.37 – -0.13<br>[-0.37 – -0.13] | 4.86E-05***<br>[3.92E-05***] | 1.00E-04*** |
| | Sex [Male] | 208.09 | 194.04 – 222.14<br>[194.05 – 222.13] | < 2.00E-16***<br>[< 2.00E-16***] | 1.00E-04*** |
| | Diagnostic Group [3q29Del] | -107.56 | -191.51 – -23.61<br>[-201.01 – -14.11] | 0.01**<br>[0.02*] | 0.03* |
| | **Diagnostic Group x Sex** | **-149.64** | **-249.96 – -49.33**<br>**[-269.37 – -29.91]** | **3.48E-03****<br>**[0.01**]** | **0.01**** |
| | $R^2$ / $R^2$ adjusted | 0.37 / 0.37 | | | |
| | Robust Wald test | 192.4 on 5 and 1625 DF, p-value < 2.20E-16*** | | | |
| | [F-statistic (OLS) | 191.3 on 5 and 1625 DF, p-value < 2.20E-16***] | | | |
| **F. eICV-Adjusted Total Cerebellum Volume ($cm^3$)** | Intercept | 144.46 | 142.99 – 145.94<br>[142.92 – 146.00] | < 2.00E-16***<br>[< 2.00E-16***] | |
| | Age (years) | -0.05 | -0.11 – 0.01<br>[-0.11 – 0.01] | 0.12<br>[0.12] | 0.12 |
| | Sex [Male] | 5.33 | 4.32 – 6.33<br>[4.33 – 6.32] | < 2.00E-16***<br>[< 2.00E-16***] | 1.00E-04*** |
| | Diagnostic Group [3q29Del] | -7.41 | -13.80 – -1.02<br>[-14.11 – -0.72] | 0.02*<br>[0.03*] | 0.03* |
| | **Group x Sex** | **3.94** | **-4.41 – 12.30**<br>**[-4.64 – 12.53]** | **0.35**<br>**[0.37]** | **0.37** |
| | $R^2$ / $R^2$ adjusted | 0.07 / 0.07 | | | |
| | Robust Wald test | 29.6 on 4 and 1626 DF, p-value < 2.20E-16*** | | | |
| | [F-statistic (OLS) | 30.3 on 4 and 1626 DF, p-value < 2.20E-16***] | | | |
| **G. eICV-Adjusted Cerebellar Cortex Volume ($cm^3$)** | Intercept | 119.89 | 118.60 – 121.17<br>[118.58 – 121.19] | < 2.00E-16***<br>[< 2.00E-16***] | |
| | Age (years) | -0.21 | -0.26 – -0.16<br>[-0.26 – -0.16] | 1.48E-15***<br>[1.50E-15***] | 1.00E-04*** |
| | Sex [Male] | 4.85 | 4.00 – 5.70<br>[4.01 – 5.70] | < 2.00E-16***<br>[< 2.00E-16***] | 1.00E-04*** |
| | Diagnostic Group [3q29Del] | -8.24 | -13.39 – -3.10<br>[-13.91 – -2.57] | 1.71E-03**<br>[4.42E-03**] | 4.30E-03** |
| | **Diagnostic Group x Sex** | **1.43** | **-5.62 – 8.48**<br>**[-5.84 – 8.70]** | **0.69**<br>**[0.70]** | **0.70** |
| | $R^2$ / $R^2$ adjusted | 0.11 / 0.11 | | | |
| | Robust Wald test | 51.0 on 4 and 1626 DF, p-value < 2.20E-16*** | | | |
| | [F-statistic (OLS) | 52.56 on 4 and 1626 DF, p-value < 2.20E-16***] | | | |

| | | | | | |
|---|---|---|---|---|---|
| **H. eICV-Adjusted Cerebellar White Matter Volume (cm$^3$)** | Intercept | 22.20 | 21.21 – 23.19 [21.19 – 23.21] | < 2.00E-16*** [< 2.00E-16***] | |
| | Age (years) | 0.43 | 0.32 – 0.53 [0.32 – 0.53] | 8.45E-15*** [1.32E-15***] | 1.00E-04*** |
| | Age$^2$ | -0.01 | -0.01 – -0.004 [-0.01 – -0.004] | 1.55E-06*** [4.10E-07***] | 1.00E-04*** |
| | Sex [Male] | 0.38 | 0.10 – 0.65 [0.09 – 0.66] | 8.15E-03** [9.49E-03**] | 9.30E-03** |
| | Diagnostic Group [3q29Del] | 1.04 | -2.56 – 4.64 [-0.85 – 2.93] | 0.57 [0.28] | 0.27 |
| | **Diagnostic Group x Sex** | **2.73** | **-2.29 – 7.75 [0.31 – 5.15]** | **0.29 [0.03*]** | **0.03*** |
| | $R^2$ / $R^2$ adjusted<br>Robust Wald test<br>[F-statistic (OLS) | 0.19 / 0.19<br>87.5 on 5 and 1625 DF, p-value < 2.20E-16***<br>78.16 on 5 and 1625 DF, p-value < 2.20E-16***] | | | |

Control *N* = 1,608 (Female *N* = 861, Male *N* = 747), 3q29Del *N* = 23 (Female *N* = 9, Male *N* = 14). p-value ≤ 0.001 '***', p-value ≤ 0.01 '**', p-value ≤ 0.05 '*', p-value ≤ 0.1 '†'. *Abbreviations*: 3q29 deletion syndrome, 3q29Del; VOI, volumetric measure of interest; eICV, estimated total intracranial volume; unstandardized coefficient estimate, *b*; confidence interval, CI; degrees of freedom, DF; permutation, perm; ordinary least squares, OLS.

**Fig. S8. Predictor effect plot showing a suggestive interaction effect between diagnostic group and sex on eICV-adjusted cerebellar white matter volumes.** Predicted values of eICV-adjusted cerebellar white matter volume across male versus female 3q29Del and control groups were computed from the exploratory interaction model reported in Table S5H, while covariates (age, age$^2$) were held fixed. Error bars indicate the 95% confidence interval. Non-robust OLS estimates, and permutation testing from Table S5H suggested a diagnostic group by sex interaction effect on eICV-adjusted cerebellar white matter volumes (non-robust $p \leq 0.05$, permutation $p \leq 0.05$), however this effect was not significant ($p > 0.05$) when robust standard error estimates were calculated to account for the heteroskedasticity in the data. We provide a graphic illustration of this finding for visual inspection of underlying trends, but we consider the evidence in favor of this sex-specific effect to be weak. Control *N* = 1,608 (Female *N* = 861, Male *N* = 747), 3q29Del *N* = 23 (Female *N* = 9, Male *N* = 14). p-value ≤ 0.001 '***', p-value ≤ 0.01 '**', p-value ≤ 0.05 '*', p-value ≤ 0.1 *'†'* *Abbreviations*: 3q29 deletion syndrome, 3q29Del; estimated total intracranial volume, eICV; ordinary least squares, OLS.

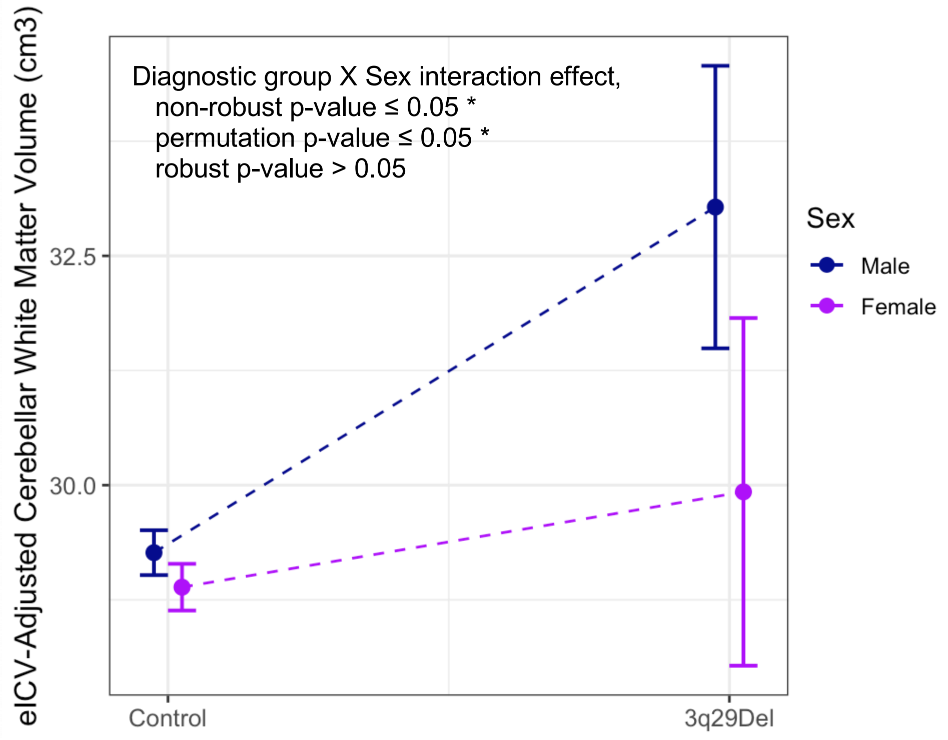

| Outcome variable | Explanatory variables | *b* | CI (95%) | p-value | perm. p-value |
|---|---|---|---|---|---|
| **eICV-Adjusted Cerebellar White Matter Volume ($cm^3$)**<br><br>**<u>Sex: Male</u>** | Intercept | 21.14 | 19.36 – 22.93<br>[19.41 – 22.88] | < 2.00E-16***<br>[< 2.00E-16***] | |
| | Age (years) | 0.56 | 0.36 – 0.75<br>[0.38 – 0.73] | 1.94E-08***<br>[8.15E-10***] | 1.00E-04*** |
| | $Age^2$ | -0.01 | -0.01 – -0.005<br>[-0.01 – -0.004] | 1.74E-04***<br>[2.48E-05***] | 1.00E-04*** |
| | **Diagnostic group [3q29Del]** | **4.09** | **0.50 – 7.68**<br>**[2.35 – 5.83]** | **0.03***<br>**[4.59E-06***]** | **1.00E-04*** |
| | $R^2$ / $R^2$ adjusted<br>Robust Wald test<br>[F-statistic (OLS) | 0.18 / 0.18<br>62.1 on 3 and 757 DF, p-value < 2.00E-16***<br>57.0 on 3 and 757 DF, p-value < 2.00E-16***] | | | |
| **eICV-Adjusted Cerebellar White Matter Volume ($cm^3$)**<br><br>**<u>Sex: Female</u>** | Intercept | 23.35 | 22.27 – 24.43<br>[22.16 – 24.54] | < 2.00E-16***<br>[< 2.00E-16***] | |
| | Age (years) | 0.32 | 0.20 – 0.44<br>[0.20 – 0.44] | 9.95E-08***<br>[3.16E-07***] | 1.00E-04*** |
| | $Age^2$ | -0.004 | -0.01 – -0.001<br>[-0.01 – -0.001] | 4.24E-03**<br>[4.83E-03**] | 4.90E-03** |
| | **Diagnostic group [3q29Del]** | **0.88** | **-2.64 – 4.39**<br>**[-0.79 – 2.54]** | **0.62**<br>**[0.30]** | **0.29** |
| | $R^2$ / $R^2$ adjusted<br>Robust Wald test<br>[F-statistic (OLS) | 0.21 / 0.20<br>89.4 on 3 and 866 DF, p-value < 2.00E-16***<br>75.5 on 3 and 866 DF, p-value < 2.00E-16***] | | | |

**Table S6. *Post hoc* analysis of the suggestive sex by diagnostic group interaction effect on eICV-adjusted cerebellar white matter volumes.** For further inspection of the suggestive diagnostic group by sex interaction effect described in Table S5H and Fig. S8, here we report results from a *post hoc* analysis of the effect of diagnostic group on eICV-adjusted cerebellar white matter volumes in males and females, separately. Sex-stratified multiple linear regression models include age and $age^2$ as covariates based on the best-fitting polynomial model for this VOI from Table S3H. Main effect of diagnostic group is reported in bold for clarity. Results indicate that male 3q29Del participants have larger eICV-adjusted cerebellar white matter volumes than male controls ($p \leq 0.05$), whereas this effect was not significant in female 3q29Del participants compared with female controls ($p > 0.05$). Inferences are based on heteroskedasticity-robust estimates in males versus females, which are provided above non-robust OLS estimates (in grey brackets). Robust Wald test statistics are reported to assess the overall significance of each model, along with exact p-values calculated by non-asymptotic permutation marginal tests. Since the heteroskedasticity-robust estimates for the corresponding sex by diagnostic group interaction failed to reach significance in Table S3H ($p > 0.05$), we consider the evidence in favor of this sex-specific effect to be weak. Male $N$ = 761 (Control $N$ = 747, 3q29Del $N$ = 14), Female $N$ = 870 (Control $N$ = 861, 3q29Del $N$ = 9). Contrast coding: reference level for the diagnostic group variable is healthy control. p-value ≤ 0.001 '***', p-value ≤ 0.01 '**', p-value ≤ 0.05 '*', p-value ≤ 0.1 '†'. *Abbreviations*: 3q29 deletion syndrome, 3q29Del; estimated total intracranial volume, eICV; VOI, volumetric measure of interest; unstandardized coefficient estimate, *b*; confidence interval, CI; degrees of freedom, DF; permutation, perm

| VOI | Summary of multiple linear regression findings | Diagnostic group | |
|---|---|---|---|
| | | **Control** Mean ± SD | **3q29Del** Mean ± SD |
| **Total Cerebellum Volume ($cm^3$)** | | | |
| **All participants** | | | |
| **Absolute Volume** | ✓ (3q29Del < control) | 145.86 ±15.17 | 132.74 ±12.81 |
| Diagnostic group X Sex | *ns* | — | — |
| **eICV-Adjusted Volume** | ✓ (3q29Del < control) | 145.86 ± 10.51 | 141.97 ± 11.31 |
| Diagnostic group X Sex | *ns* | — | — |
| **Male participants** | | | |
| **Absolute Volume** | ✓ (3q29Del < control) | 154.76 ± 13.47 | 137.58 ± 9.78 |
| **eICV-Adjusted Volume** | ✓ (3q29Del < control) | 148.72 ± 10.80 | 145.64 ± 10.48 |
| **Female participants** | | | |
| **Absolute Volume** | ✓ (3q29Del < control) | 138.13 ± 12.00 | 125.22 ± 13.83 |
| **eICV-Adjusted Volume** | ✓ (3q29Del < control) | 143.37 ± 9.59 | 136.26 ± 10.63 |
| **Cerebellar Cortex Volume ($cm^3$)** | | | |
| **All participants** | | | |
| **Absolute Volume** | ✓ (3q29Del < control) | 117.39 ± 12.52 | 105.34 ± 11.55 |
| Diagnostic group X Sex | *ns* | — | — |
| **eICV-Adjusted Volume** | ✓ (3q29Del < control) | 117.39 ± 9.12 | 112.31 ± 9.82 |
| Diagnostic group X Sex | *ns* | — | — |
| **Male participants** | | | |
| **Absolute Volume** | ✓ (3q29Del < control) | 124.61 ± 10.98 | 108.88 ± 9.35 |
| **eICV-Adjusted Volume** | ✓ (3q29Del < control) | 120.06 ± 9.21 | 114.96 ± 9.31 |
| **Female participants** | | | |
| **Absolute Volume** | ✓ (3q29Del < control) | 111.12 ± 10.18 | 99.84 ± 12.99 |
| **eICV-Adjusted Volume** | ✓ (3q29Del < control) | 115.07 ± 8.38 | 108.18 ± 9.63 |
| **Cerebellar White Matter Volume ($cm^3$)** | | | |
| **All participants** | | | |
| **Absolute Volume** | *ns* | 28.47 ± 3.89 | 27.40 ± 5.84 |
| Diagnostic group X Sex | *ns* | — | — |
| **eICV-Adjusted Volume** | ✓ (3q29Del > control) | 28.47 ± 3.13 | 29.66 ± 6.11 |
| Diagnostic group X Sex | ✓ (suggestive interaction effect) | — | — |
| **Male participants** | | | |
| **Absolute Volume** | *ns* | 30.15 ± 4.02 | 28.70 ± 6.35 |
| **eICV-Adjusted Volume** | ✓ (3q29Del > control; inspection of suggestive interaction effect) | 28.67 ± 3.46 | 30.68 ± 6.64 |
| **Female participants** | | | |
| **Absolute Volume** | *ns* | 27.01 ± 3.12 | 25.37 ± 4.54 |
| **eICV-Adjusted Volume** | *ns* (inspection of suggestive interaction effect) | 28.30 ± 2.80 | 28.08 ± 5.12 |

| **Cerebellar Cortex to White Matter Volume Ratio** | | | |
|---|---|---|---|
| **All participants** | | | |
| **Absolute Volume Ratio** | ✓ (3q29Del < control) | 4.16 ± 0.46 | 3.99 ± 0.83 |
| Diagnostic group X Sex | *ns* | — | — |
| **Male participants** | | | |
| **Absolute Volume Ratio** | ✓ (3q29Del < control) | 4.18 ± 0.48 | 3.97 ± 0.90 |
| **Female participants** | | | |
| **Absolute Volume Ratio** | ✓ (3q29Del < control) | 4.15 ± 0.45 | 4.02 ± 0.76 |
| **eICV ($cm^3$)** | | | |
| **All participants** | | | |
| **Absolute Volume** | ✓ (3q29Del < control) | 1587.49 ± 178.02 | 1413.33 ± 116.47 |
| Diagnostic group X Sex | ✓ (male effect > female effect) | — | — |
| **Male participants** | | | |
| **Absolute Volume** | ✓ (3q29Del < control) | 1701.33 ± 141.52 | 1435.33 ± 103.17 |
| **Female participants** | | | |
| **Absolute Volume** | ✓ (3q29Del < control) | 1488.71 ± 144.31 | 1379.12 ± 133.59 |

**Table S7. Summary of multiple linear regression findings and descriptive statistics for volumetric measures of interest in 3q29Del and control groups.** Diagnostic group differences and diagnostic group by sex interaction effects identified in the current study (*p's* ≤ 0.05) are marked by the "✓" symbol; the direction of the corresponding effect is specified by the ">" (greater than) or "<" (less than) symbols. Descriptive statistics (mean and SD) for each volumetric measure of interest are provided on the right for separate groups. Control *N* = 1,608 (Female *N* = 861, Male *N* = 747), 3q29Del *N* = 23 (Female *N* = 9, Male *N* = 14). *Abbreviations*: 3q29 deletion syndrome, 3q29Del; VOI, volumetric measure of interest; estimated total intracranial volume, eICV; standard deviation, SD; not significant, ns.

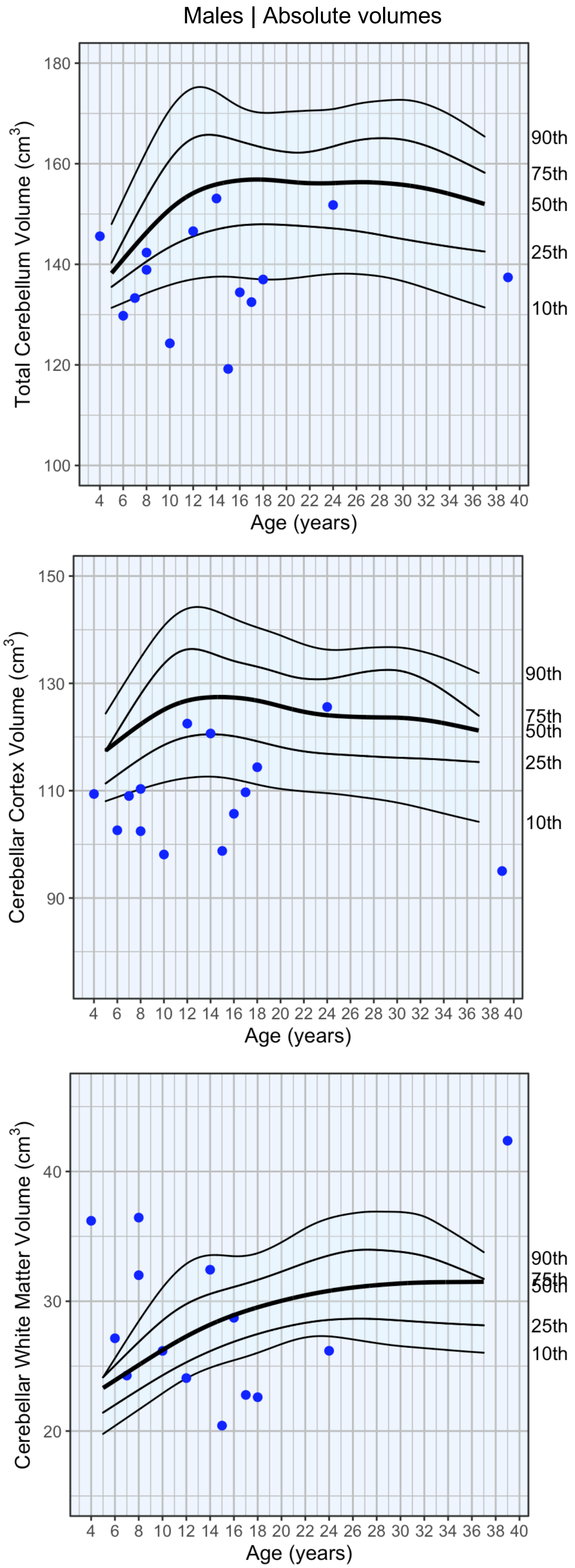

Males | Absolute volumes
Total Cerebellum Volume (cm$^3$)
90th
75th
50th
25th
10th
Age (years)
Cerebellar Cortex Volume (cm$^3$)
90th
75th
50th
25th
10th
Age (years)
Cerebellar White Matter Volume (cm$^3$)
90th
75th
50th
25th
10th
Age (years)

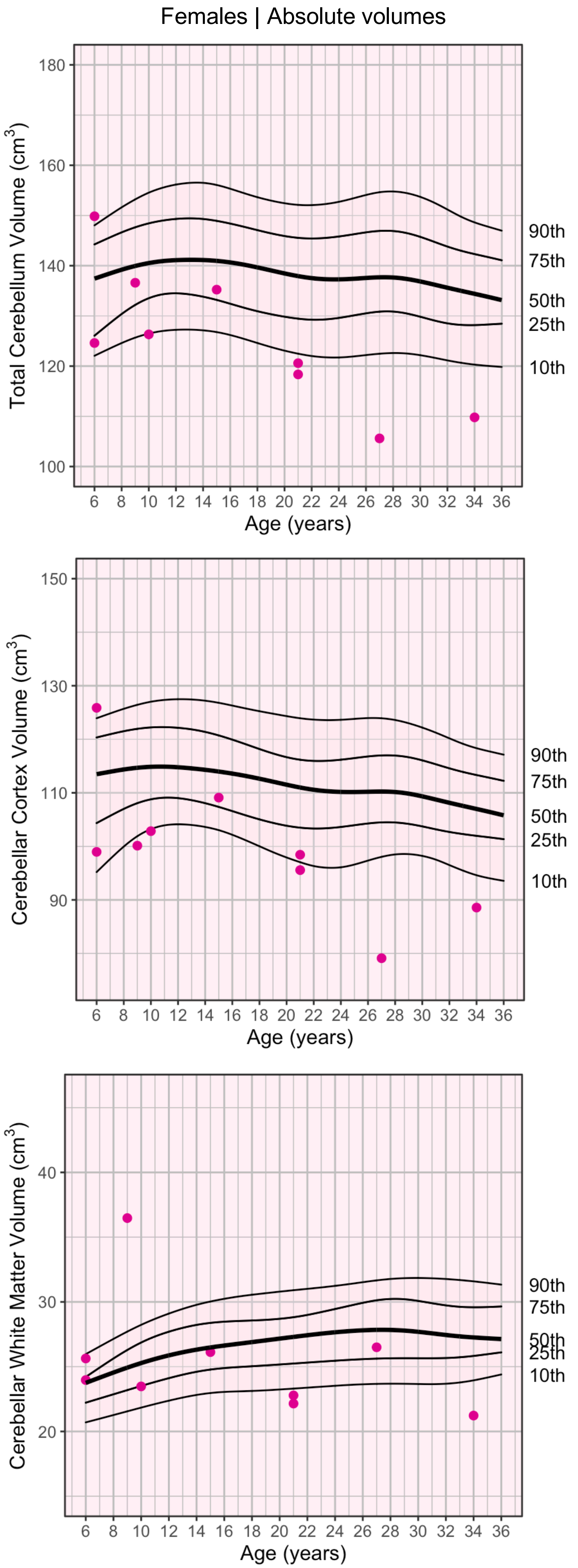

Females | Absolute volumes
Total Cerebellum Volume (cm$^3$)
90th
75th
50th
25th
10th
Age (years)
Cerebellar Cortex Volume (cm$^3$)
90th
75th
50th
25th
10th
Age (years)
Cerebellar White Matter Volume (cm$^3$)
90th
75th
50th
25th
10th
Age (years)

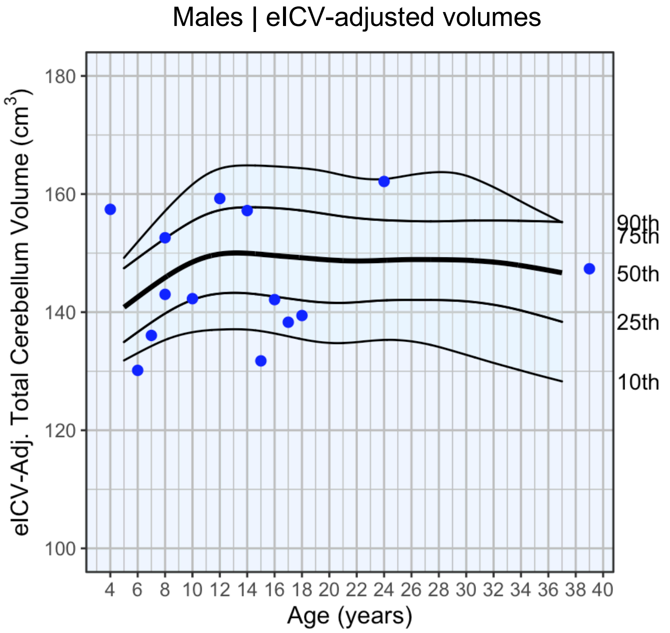
Males | eICV-adjusted volumes
eICV-Adj. Total Cerebellum Volume (cm$^3$)
Age (years)
90th
75th
50th
25th
10th

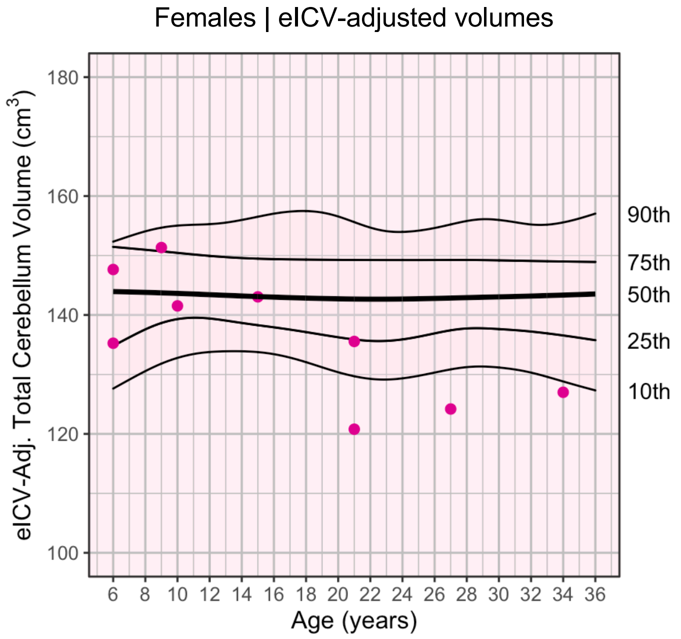
Females | eICV-adjusted volumes
eICV-Adj. Total Cerebellum Volume (cm$^3$)
Age (years)
90th
75th
50th
25th
10th

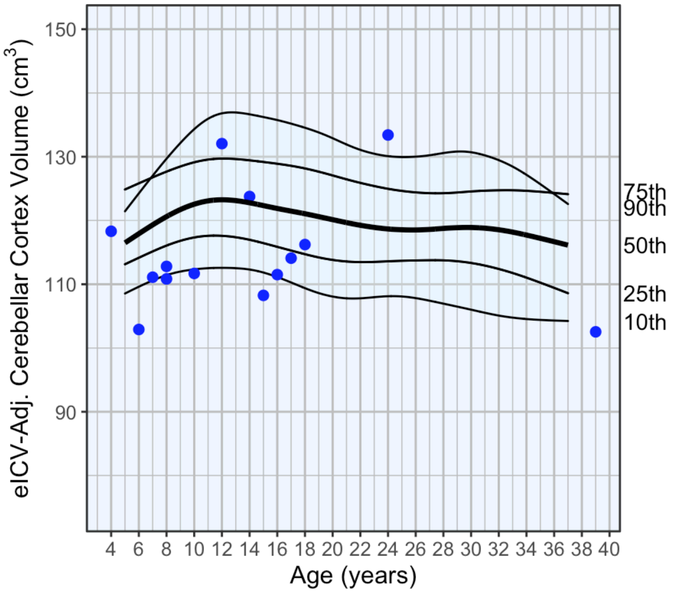
eICV-Adj. Cerebellar Cortex Volume (cm$^3$)
Age (years)
75th
90th
50th
25th
10th

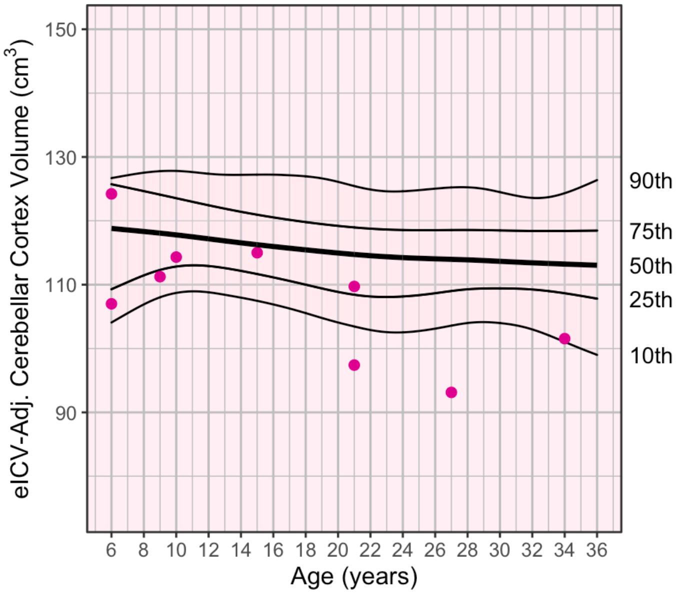
eICV-Adj. Cerebellar Cortex Volume (cm$^3$)
Age (years)
90th
75th
50th
25th
10th

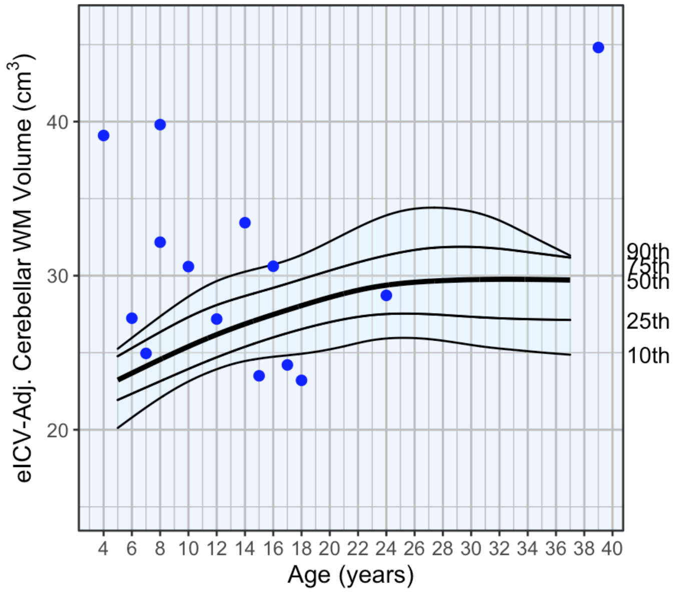
eICV-Adj. Cerebellar WM Volume (cm$^3$)
Age (years)
90th
75th
50th
25th
10th

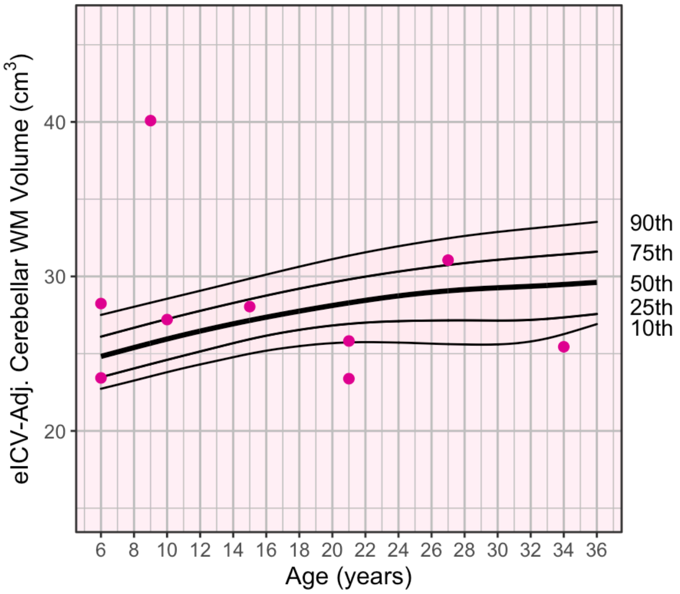
eICV-Adj. Cerebellar WM Volume (cm$^3$)
Age (years)
90th
75th
50th
25th
10th

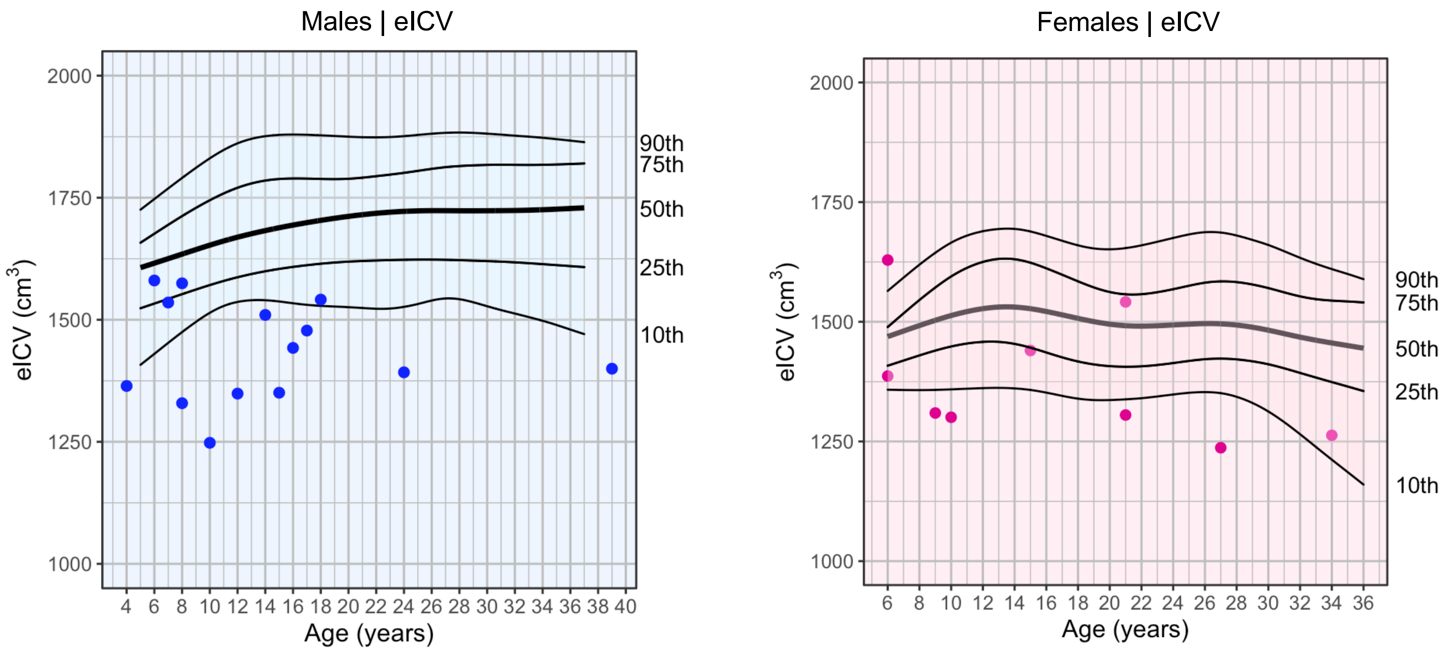

**Fig. S9. Estimated normative percentile curves for cerebellar volumes and eICV, stratified by sex.** Black solid lines represent the 10th, 25th, 50th (median, thicker), 75th, and 90th percentiles estimated by fitting quantile splines to volumetric data from *N* = 861 female healthy controls and *N* = 747 male healthy controls, separately. Pink dots represent data points for *N = 9* 3q29Del female participants and blue dots represent data points for *N = 14* 3q29Del male participants, whose volumetric measures are being compared to sex-stratified reference percentiles in healthy controls. A spline term of age was included as a covariate in each quantile spline model, as described by Oh et al. (2004). Due to the relative sparsity of volumetric data at the youngest and oldest endpoints of the age range covered in the present study, slight overlaps are observed in percentile bands. *Abbreviations*: 3q29 deletion syndrome, 3q29Del; estimated total intracranial volume, eICV; adjusted, adj.

Oh HS, Nychka D, Brown T, Charbonneau P (2004): Period analysis of variable stars by robust smoothing. *Journal of the Royal Statistical Society: Series C (Applied Statistics)*. *53*(1):15-30.

| | PFAC/MCM - (*N* = 11) | PFAC/MCM + (*N* = 13) | Test statistics |
|---|---|---|---|
| **Sex**, *n/N (%)* | | | |
| Male | 7/11 (63.64%) | 8/13 (61.54%) | OR = 0.92,<br>95% CI = 0.13 – 6.39,<br>p-value [a] = 1 |
| Female | 4/11 (36.36%) | 5/13 (38.46%) | |
| **Age** (in years) | | | |
| Mean ± SD | 17.64 ± 11.76 | 12.23 ± 5.72 | *t = -1.47,* DF = 22,<br>p-value [b] = 0.16[b] |
| Median [Range] | 15 [4 – 39] | 10 [6 – 21] | |
| **Ethnicity**, *n/N (%)* | | | |
| Non-Hispanic / Latino | 11/11 (100%) | 12/13 (92.31%) | OR = INF,<br>95% CI = 0.02 – INF,<br>*p-value* [a] = 1 |
| Hispanic / Latino | 0/11 (0%) | 1/13 (7.69%) | |
| **Race**, *n/N (%)* | | | |
| White | 10/11 (90.91%) | 12/13 (92.31%) | OR = 0.84,<br>95% CI = 0.01 – 71.84<br>p-value [a] = 1 |
| More than one race | 1/11 (9.09%) | 1/13 (7.69%) | |
| **History of head injury**[#], *n/N (%)* | | | |
| Negative | 6/8 (75.00%) | 11/13 (84.62%) | OR = 0.56,<br>95% CI = 0.03 – 9.62,<br>p-value [a] = 0.62 |
| Positive | 2/8 (25.00%) | 2/13 (15.38%) | |
| **History of neonatal complications during delivery**[#], *n/N (%)*<br>*"Did the baby have any trouble at birth?" (e.g., birth injuries, jaundice, fetal hypoxia, respiratory distress syndrome, hemorrhage); "Was the baby premature?"* | | | |
| Negative | 3/8 (37.50%) | 7/13 (53.85%) | OR = 0.53,<br>95% CI = 0.06 – 4.21,<br>p-value [a] = 0.66 |
| Positive | 5/8 (62.50%) | 6/13 (46.15%) | |
| **History of maternal complications during pregnancy**[#], *n/N (%)*<br>*"Did the mother have any illness or injury during pregnancy?" (e.g., preeclampsia, infection, physical trauma)* | | | |
| Negative | 4/8 (50.00%) | 9/13 (69.23%) | OR = 0.46,<br>95% CI = 0.05 – 3.90,<br>p-value [a] = 0.65 |
| Positive | 4/8 (50.00%) | 4/13 (30.77%) | |
| **Combined history of head injury, neonatal complications during delivery, and/or maternal complications during pregnancy**[#], *n/N (%)* | | | |
| Negative | 2/8 (25.00%) | 4/13 (30.77%) | OR= 0.76,<br>95% CI= 0.05 – 7,55,<br>p-value [a] = 1 |
| Positive | 6/8 (75.00%) | 9/13 (69.23%) | |

**Table S8. Demographic and relevant clinical characteristics of 3q29Del participants with versus without posterior fossa arachnoid cyst or mega cisterna magna findings.** There was no significant difference between PFAC/MCM positive (+) and negative (-) 3q29Del participants in demographic characteristics or in clinical characteristics that have been proposed to be associated with the etiology of secondary (acquired) cysts in previous literature (*p's* > 0.05). [a]Fisher's exact test, [b]Student's two sample t-test. [#]3q29Del *N* = 21 due to missing data. *Abbreviations:* 3q29 deletion syndrome, 3q29Del; posterior fossa arachnoid cyst, PFAC; mega cisterna magna, MCM; standard deviation, SD; odds ratio, OR; confidence interval, CI.

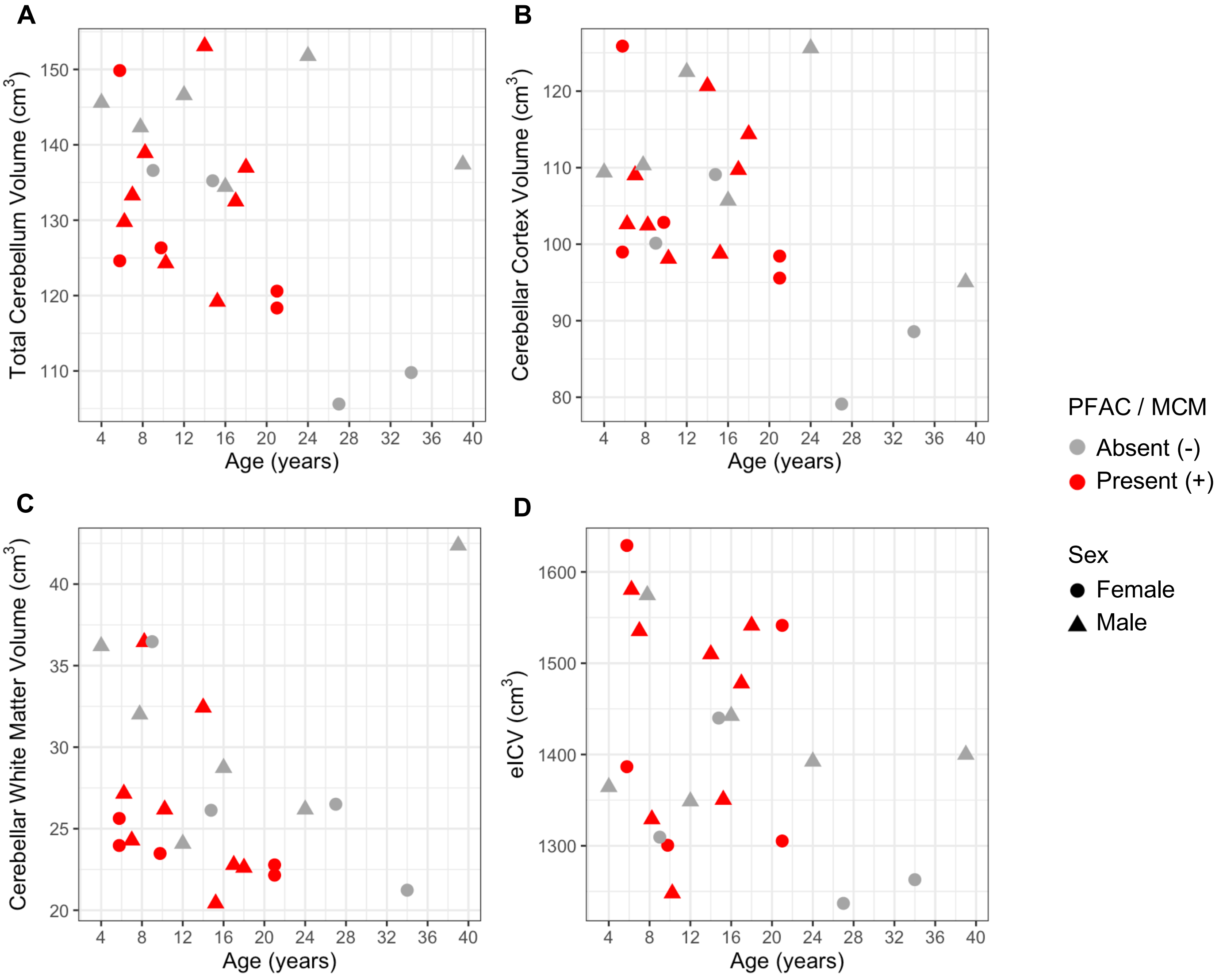

**Fig. S10. Scatter plots showing the distribution of A) total cerebellum volume, B) cerebellar cortex volume, C) cerebellar white matter volume, and D) eICV as a function of age among 3q29Del participants with versus without posterior fossa arachnoid cyst or mega cisterna magna findings.** A slight jitter was added systematically to all panels to minimize overplotting. 3q29Del *N* = 23 (Female *N* = 9, Male *N* = 14). *Abbreviations*: 3q29 deletion syndrome, 3q29Del; estimated total intracranial volume, eICV; posterior fossa arachnoid cyst, PFAC; mega cisterna magna, MCM.

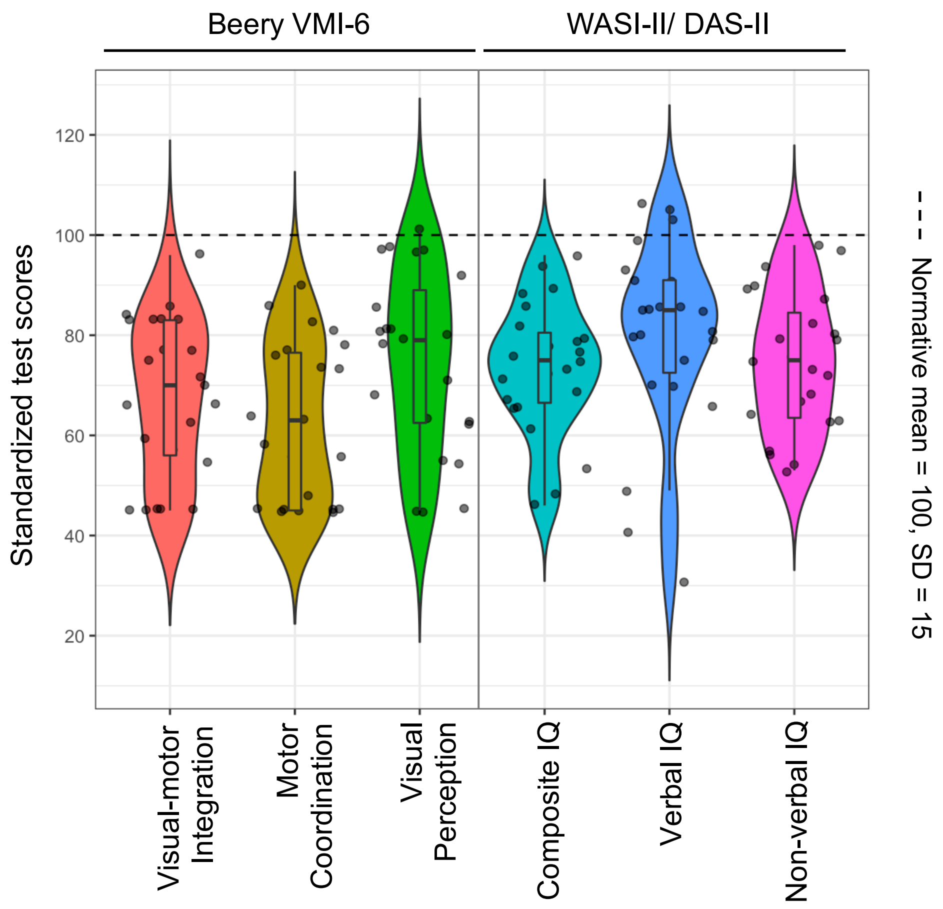

**Fig. S11. Violin plots with box plots visualizing the distribution of standardized test scores for sensorimotor and cognitive abilities among 3q29Del participants.** Violin plots represent the distribution of standardized test scores (normative mean = 100, SD = 15) for visual-motor integration, motor coordination and visual perception skills measured by the Beery-Buktenica Developmental Test of VMI, and for composite, verbal and non-verbal IQ scores measured by the WASI / DAS. Boxplots visualize the five-number summary statistics for each measure (minimum, lower quartile, median, upper quartile and maximum). Higher scores indicate better performance. 3q29Del *N* = 23. *Abbreviations*: 3q29 deletion syndrome, 3q29Del; intelligence quotient, IQ; Wechsler Abbreviated Scale of Intelligence, WASI; Differential Ability Scales, DAS; visual-motor integration, VMI; standard deviation, SD.

| **Standardized test scores** | **3q29Del** (*N* = 23) |
|---|---|
| **Visual-motor Integration** (*Beery VMI-6)* | |
| Mean ± SD | 67.83 ± 15.87 |
| Median [Range] | 70 [45 – 96] |
| **Supplemental test: Motor** | |
| **Coordination** (*Beery VMI-6)* | 62.65 ± 15.68 |
| Mean ± SD | 63 [45 – 90] |
| Median [Range] | |
| **Supplemental test: Visual** | |
| **Perception** (*Beery VMI-6)* | 74.74 ± 18.20 |
| Mean ± SD | 79 [45 – 101] |
| Median [Range] | |
| **Composite IQ** *(WASI-II/ DAS-II)* | |
| Mean ± SD | 73.48 ± 13.30 |
| Median [Range] | 75 [46 – 96] |
| **Supplemental test: Verbal IQ** | |
| *(WASI-II/ DAS-II)* | 79.87 ± 19.13 |
| Mean ± SD | 85 [31 – 106] |
| Median [Range] | |
| **Supplemental test: Non-verbal IQ** | |
| *(WASI-II/ DAS-II)* | 74.61 ± 13.80 |
| Mean ± SD | 75 [53 – 98] |
| Median [Range] | |

**Table. S9. Descriptive statistics for standardized test scores for sensorimotor and cognitive abilities among 3q29Del participants.** *Abbreviations*: 3q29 deletion syndrome, 3q29Del; intelligence quotient, IQ; Wechsler Abbreviated Scale of Intelligence, WASI; Differential Ability Scales, DAS; visual-motor integration, VMI; standard deviation, SD.

| Outcome variables | Explanatory variables | *b* | CI (95%) | p-value |
|---|---|---|---|---|
| **Visual-motor Integration** *(standardized score, Beery VMI-6)* | Intercept | 100.65 | 17.78 – 183.51 | 0.02* |
| | Age (years) | -0.63 | -1.72 – 0.46 | 0.24 |
| | Sex [Male] | -4.22 | -19.46 – 11.02 | 0.57 |
| | **Cerebellar Cortex Volume (cm$^3$)** | **-0.20** | **-0.95 – 0.56** | **0.59** |
| | $R^2$ / $R^2$ adjusted | 0.13 / -0.01 | | |
| | Robust Wald Test | 0.6 on 3 and 19 DF, p-value = 0.63 | | |
| | Intercept | 44.93 | 15.79 – 74.07 | 4.44E-03** |
| | Age (years) | -0.57 | -1.24 – 0.10 | 0.09† |
| | Sex [Male] | -10.47 | -24.59 – 3.65 | 0.14 |
| | **Cerebellar WM Volume (cm$^3$)** | **1.38** | **0.36 – 2.41** | **0.01**** |
| | $R^2$ / $R^2$ adjusted | 0.35 / 0.25 | | |
| | Robust Wald Test | 4.0 on 3 and 19 DF, p-value = 0.02* | | |
| **Supplemental test: Motor Coordination** *(standardized score, Beery VMI-6)* | Intercept | 44.27 | -44.76 – 133.29 | 0.31 |
| | Age (years) | 0.28 | -0.71 – 1.28 | 0.56 |
| | Sex [Male] | -5.78 | -20.33 – 8.78 | 0.42 |
| | **Cerebellar Cortex Volume (cm$^3$)** | **0.17** | **-0.60 – 0.94** | **0.65** |
| | $R^2$ / $R^2$ adjusted | 0.05 / -0.10 | | |
| | Robust Wald Test | 0.4 on 3 and 19 DF, p-value = 0.77 | | |
| | Intercept | 46.45 | 15.79 – 77.11 | 5.03E-03** |
| | Age (years) | 0.20 | -0.53 – 0.92 | 0.58 |
| | Sex [Male] | -6.58 | -21.59 – 8.42 | 0.37 |
| | **Cerebellar WM Volume (cm$^3$)** | **0.63** | **-0.45 – 1.71** | **0.24** |
| | $R^2$ / $R^2$ adjusted | 0.09 / -0.05 | | |
| | Robust Wald Test | 0.8 on 3 and 19 DF, p-value = 0.53 | | |
| **Supplemental test: Visual Perception** *(standardized score, Beery VMI-6)* | Intercept | 109.13 | 24.51 – 193.76 | 0.01** |
| | Age (years) | -0.60 | -1.68 – 0.48 | 0.26 |
| | Sex [Male] | -5.46 | -22.30 – 11.38 | 0.51 |
| | **Cerebellar Cortex Volume (cm$^3$)** | **-0.21** | **-0.96 – 0.54** | **0.57** |
| | $R^2$ / $R^2$ adjusted | 0.10 / -0.04 | | |
| | Robust Wald Test | 0.6 on 3 and 19 DF, p-value = 0.62 | | |
| | Intercept | 65.04 | 30.90 – 99.18 | 7.88E-04*** |
| | Age (years) | -0.52 | -1.34 – 0.29 | 0.19 |
| | Sex [Male] | -10.06 | -30.39 – 10.28 | 0.31 |
| | **Cerebellar WM Volume (cm$^3$)** | **0.87** | **-0.83 – 2.56** | **0.30** |
| | $R^2$ / $R^2$ adjusted | 0.16 / 0.03 | | |
| | Robust Wald Test | 0.8 on 3 and 19 DF, p-value = 0.52 | | |
| **Composite IQ** *(standardized score, WASI-II/ DAS-II)* | Intercept | 62.45 | -4.14 – 129.04 | 0.06† |
| | Age (years) | 0.06 | -0.86 – 0.97 | 0.90 |
| | Sex [Male] | -5.15 | -18.74 – 8.44 | 0.44 |
| | **Cerebellar Cortex Volume (cm$^3$)** | **0.13** | **-0.47 – 0.72** | **0.66** |
| | $R^2$ / $R^2$ adjusted | 0.03 / -0.12 | | |
| | Robust Wald Test | 0.2 on 3 and 19 DF, p-value = 0.88 | | |
| | Intercept | 42.69 | 19.34 – 66.03 | 1.14E-03** |
| | Age (years) | -0.03 | -0.56 – 0.50 | 0.90 |
| | Sex [Male] | -8.66 | -20.51 – 3.18 | 0.14 |
| | **Cerebellar WM Volume (cm$^3$)** | **1.33** | **0.36 – 2.31** | **9.89E-03**** |
| | $R^2$ / $R^2$ adjusted | 0.34 / 0.23 | | |
| | Robust Wald Test | 3.1 on 3 and 19 DF, p-value = 0.05* | | |

| | | | | |
|---|---|---|---|---|
| | Intercept | 46.24 | -32.13 – 124.61 | 0.23 |
| | Age (years) | 0.28 | -0.72 – 1.29 | 0.56 |
| | Sex [Male] | -9.09 | -25.92 – 7.74 | 0.27 |
| | **Cerebellar Cortex Volume ($cm^3$)** | **0.33** | **-0.36 – 1.02** | **0.33** |
| **Supplemental test: Verbal IQ** *(standardized score, WASI-II/ DAS-II)* | $R^2$ / $R^2$ adjusted | 0.06 / -0.08 | | |
| | Robust Wald Test | 0.5 on 3 and 19 DF, p-value = 0.66 | | |
| | Intercept | 43.93 | 12.21 – 75.64 | 9.20E-03** |
| | Age (years) | 0.10 | -0.65 – 0.85 | 0.78 |
| | Sex [Male] | -11.58 | -28.35 – 5.18 | 0.16 |
| | **Cerebellar WM Volume ($cm^3$)** | **1.51** | **0.13 – 2.89** | **0.03*** |
| | $R^2$ / $R^2$ adjusted | 0.23 / 0.11 | | |
| | Robust Wald Test | 2.9 on 3 and 19 DF, p-value = 0.06† | | |
| | Intercept | 94.11 | 33.85 – 154.37 | 4.04E-03** |
| | Age (years) | -0.52 | -1.55 – 0.50 | 0.30 |
| | Sex [Male] | -5.40 | -19.42 – 8.63 | 0.43 |
| | **Cerebellar Cortex Volume ($cm^3$)** | **-0.08** | **-0.64 – 0.48** | **0.77** |
| **Supplemental test: Non-verbal IQ** *(standardized score, WASI-II/ DAS-II)* | $R^2$ / $R^2$ adjusted | 0.14 / 0.003 | | |
| | Robust Wald Test | 0.5 on 3 and 19 DF, p-value = 0.67 | | |
| | Intercept | 53.59 | 23.85 – 83.33 | 1.29E-03** |
| | Age (years) | -0.52 | -1.11 – 0.07 | 0.08† |
| | Sex [Male] | -10.37 | -22.86 – 2.12 | 0.10† |
| | **Cerebellar WM Volume ($cm^3$)** | **1.28** | **0.17 – 2.39** | **0.03*** |
| | $R^2$ / $R^2$ adjusted | 0.41 / 0.31 | | |
| | Robust Wald Test | 3.0 on 3 and 19 DF, p-value = 0.6† | | |

**Table S10. Extended multiple linear regression results testing the relationships between tissue-specific cerebellar volumes and sensorimotor and cognitive abilities among 3q29Del participants.** Sex and age were included as covariates in each regression model. Main effects of cerebellar cortex and white matter volumes are reported in bold for clarity. Regression parameters reflect heteroskedasticity-robust estimates, which were computed to address observed violations of statistical assumptions. Robust Wald test statistics are reported to assess the overall significance of each model. Regression results indicate significant relationships between cerebellar white matter volume and standardized test scores for visual-motor integration skills ($p \leq 0.01$) and composite IQ ($p \leq 0.01$) among 3q29Del participants, while correcting for age and sex. In supplemental analyses, cerebellar white matter volume was found to have a significant relationship with both verbal ($p \leq 0.05$), and non-verbal IQ ($p \leq 0.05$). See Table 5 for results from secondary models including estimated total intracranial volume as an additional covariate to account for global variability in head size. 3q29Del $N$ = 23. Contrast coding: reference level for the sex variable is female. p-value ≤ 0.001 '***', p-value ≤ 0.01 '**', p-value ≤ 0.05 '*', p-value ≤ 0.1 '†'. *Abbreviations*: 3q29 deletion syndrome, 3q29Del; volumetric measure of interest, VOI; intelligence quotient, IQ; Wechsler Abbreviated Scale of Intelligence, WASI; Differential Ability Scales, DAS; visual-motor integration, VMI; white matter, WM; unstandardized coefficient estimate, *b*; confidence interval, CI; degrees of freedom, DF.

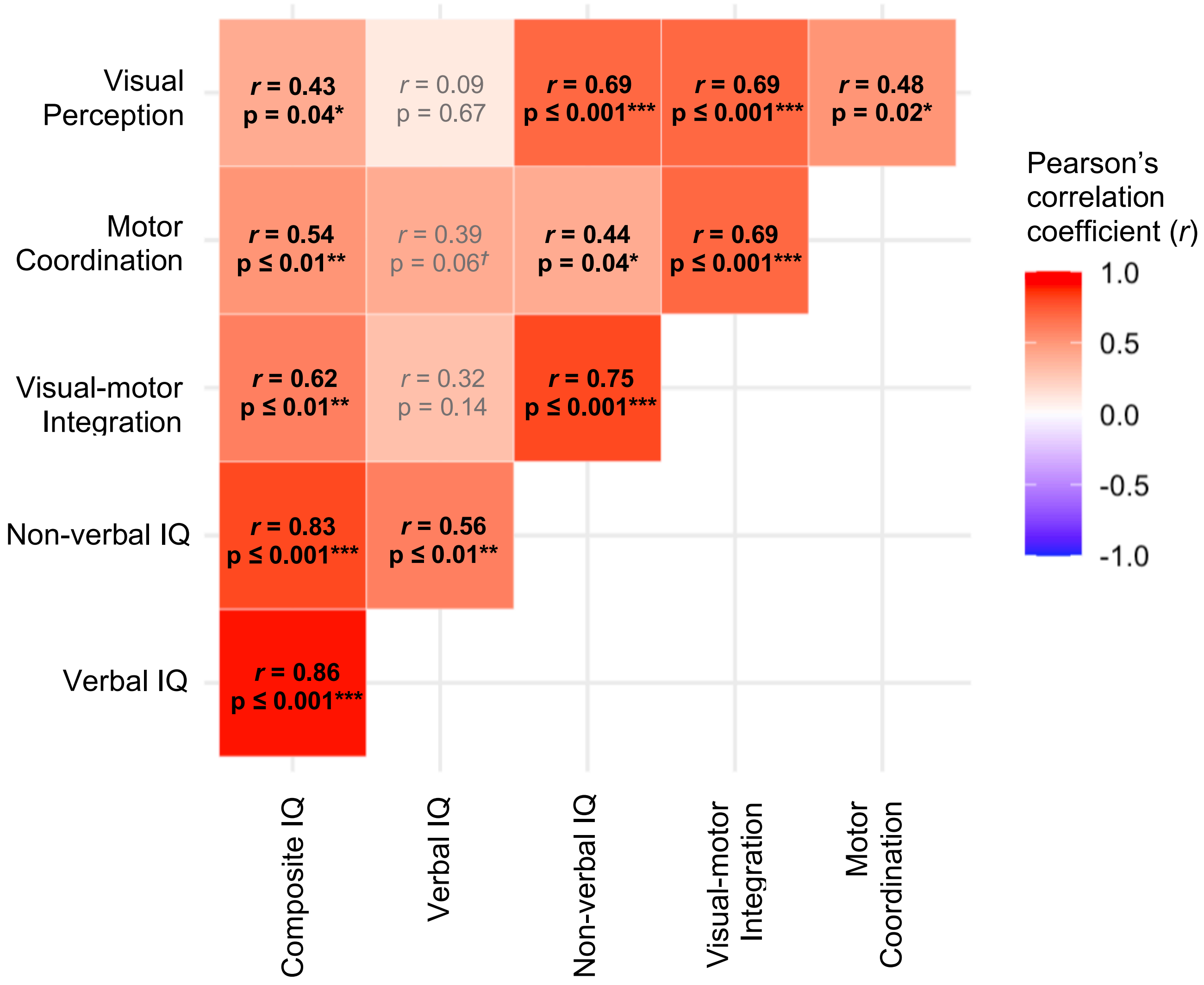

**Fig. S12. Heatmap visualization of pairwise Pearson's correlations between standardized test scores for sensorimotor and cognitive abilities among 3q29Del participants.** We assessed the pairwise Pearson's correlations among all behavioral measures to determine the extent to which our findings may be interrelated. In the 3q29Del sample, composite IQ, verbal IQ and non-verbal IQ scores were significantly correlated with one another (*p's* ≤ 0.01, moderate-very strong). Similarly, visual-motor integration, motor coordination, and visual perception scores showed significant pairwise correlations with each other (*p's* ≤ 0.05, moderate-strong). Composite IQ scores had significant correlations with visual-motor integration (*p* ≤ 0.01, strong), motor coordination (*p* ≤ 0.01, moderate), and visual perception scores (*p* ≤ 0.05, moderate), which were largely driven by significant correlations observed between non-verbal IQ and motor coordination scores (*p* ≤ 0.05, moderate) and between non-verbal IQ and visual perception scores (*p* ≤ 0.001, strong). Verbal IQ scores did not correlate significantly with visual-motor integration, motor coordination, or visual perception scores (*p's* > 0.05, very weak-weak). All correlation coefficients (*r*) were positive. Significant test results (*p's* ≤ 0.05) are reported in bold for clarity. The strength of the computed correlation coefficients was evaluated based on the following criteria: |*r*| = 0 – 0.19, very weak; |*r*| = 0.20 – 0.39, weak; |*r*| = 0.40 – 0.59, moderate; |*r*| = 0.60 – 0.79, strong; |*r*| = 0.80 – 1, very strong. 3q29Del *N* = 23. p-value ≤ 0.001 '***', p-value ≤ 0.01 '**', p-value ≤ 0.05 '*', p-value ≤ 0.1 '†'. *Abbreviations*: 3q29 deletion syndrome, 3q29Del; IQ, intelligence quotient.

| Standardized test scores | PFAC/MCM - (*N* = 11) | PFAC/MCM + (*N* = 13) | Test statistics |
|---|---|---|---|
| **Visual-motor Integration** (*Beery VMI-6)* | | | |
| Mean ± SD | 66.64 ± 16.45 | 68.85 ± 15.30 | *t* = 0.34, DF = 22, |
| Median [Range] | 68 [45 – 86] | 70 [45 – 96] | p-value = 0.74[a] |
| **Supplemental test: Motor Coordination** (*Beery VMI-6)* | | | |
| Mean ± SD | 64.00 ± 15.95 | 60.38 ± 15.79 | *W* = 60.00, |
| Median [Range] | 63 [45 – 90] | 58 [45 – 83] | p-value = 0.52[b] |
| **Supplemental test: Visual Perception** (*Beery VMI-6)* | | | |
| Mean ± SD | 69.09 ± 19.36 | 78.38 ± 16.41 | *t* = 1.27, DF = 22, |
| Median [Range] | 63 [45 – 98] | 79 [45 – 101] | p-value = 0.22[a] |
| **Composite IQ** *(WASI-II/ DAS-II)* | | | |
| Mean ± SD | 74.91 ± 12.83 | 70.77 ± 14.45 | *t* = -0.74, DF = 22, |
| Median [Range] | 75 [54 – 96] | 72 [46 – 89] | p-value = 0.47[a] |
| **Supplemental test: Verbal IQ** *(WASI-II/ DAS-II)* | | | |
| Mean ± SD | 84.73 ± 17.19 | 74.00 ± 20.24 | *W* = 52.50, |
| Median [Range] | 85 [57 – 106] | 80 [31 – 93] | p-value = 0.28[b] |
| **Supplemental test: Non-verbal IQ** *(WASI-II/ DAS-II)* | | | |
| Mean ± SD | 73.09 ± 12.71 | 75.54 ± 14.60 | *t* = 0.43, DF = 22, |
| Median [Range] | 72 [54 – 98] | 79 [53 – 97] | p-value = 0.67[a] |

**Table S11. Standardized test scores for sensorimotor and cognitive abilities in 3q29Del participants with versus without posterior fossa arachnoid cyst or mega cisterna magna findings.** There was no significant difference between PFAC/MCM positive (+) versus negative (-) 3q29Del participants in visual-motor integration, motor coordination or visual perception scores measured by the Beery-Buktenica Developmental Test of VMI, or in composite, verbal or non-verbal IQ scores measured by the WASI / DAS (*p's* > 0.05). [a]Student's two sample t-test, [b]Wilcoxon rank sum test with continuity correction. Non-parametric statistics are reported in cases where the data do not meet parametric assumptions. *Abbreviations*: 3q29 deletion syndrome, 3q29Del; posterior fossa arachnoid cyst, PFAC; mega cisterna magna, MCM; intelligence quotient, IQ; WASI, Wechsler Abbreviated Scale of Intelligence; DAS, Differential Ability Scales; VMI, visual-motor integration; standard deviation, SD.

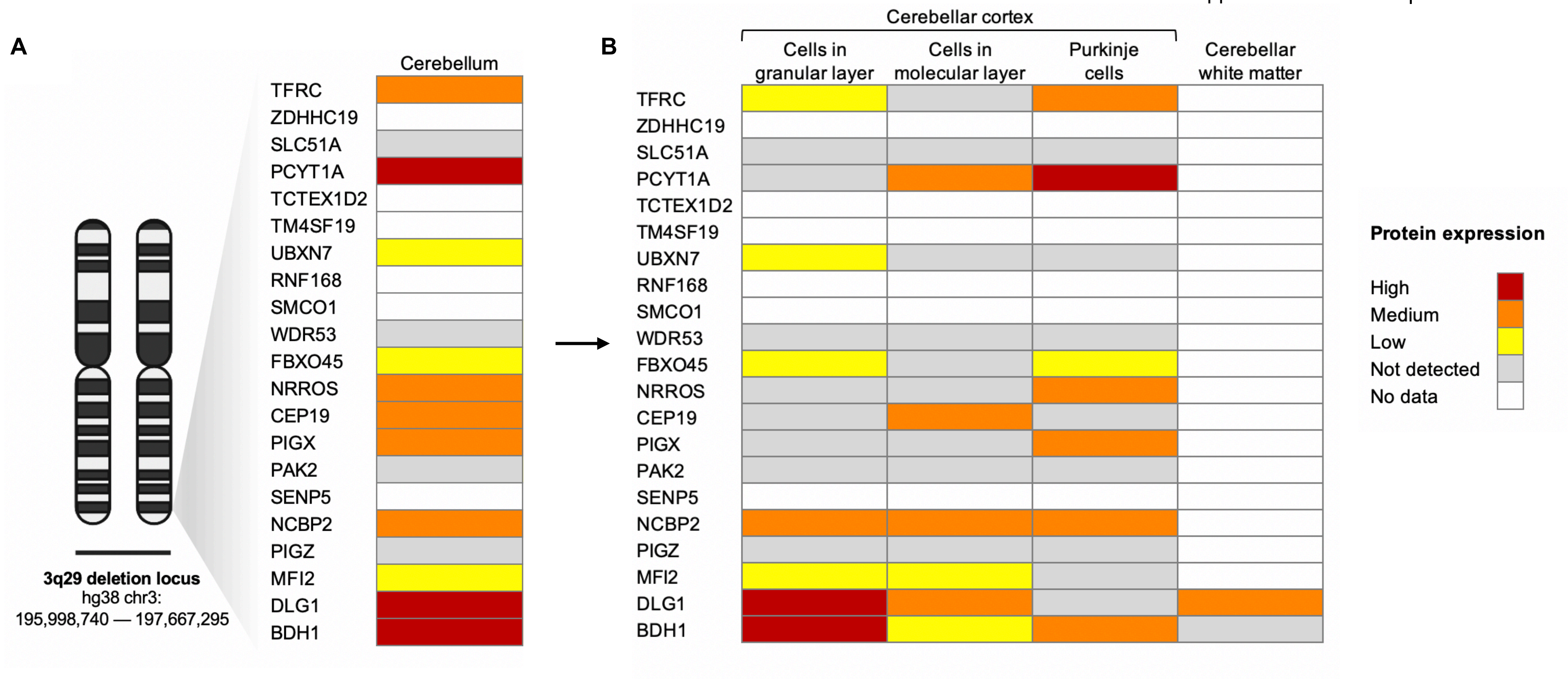

**Fig. S13. Cerebellar protein expression profiles of 3q29 interval genes annotated by the Human Protein Atlas**. In **A)** protein expression profiles of 21 protein coding genes located in the 3q29 interval are provided for the human cerebellum. In **B)** protein expression profiles of 3q29 interval genes are provided for annotated cerebellar subregions, using the same units. All annotations were obtained from the Human Protein Atlas (version 20.1) and were established by evaluation of immunohistochemical staining patterns, RNA-sequencing data, and available protein/gene characterization data, as described by Uhlen et al. (2010 & 2015). Protein expression profiles were characterized in normal human tissues as “High”, “Medium”, “Low” or “Not detected”. The “No data” category indicates that there was no available protein expression information on the Human Protein Atlas for a given query. The cerebellar cortex includes three layers: 1) the granular layer, which contains granule cells, golgi cells and synaptic glomeruli, 2) the molecular layer, which contains axons from the cells of the granular layer, Purkinje dendrites, astrocytes and neuronal cell bodies (e.g., stellate and basket cells), and 3) the Purkinje layer, which contains Purkinje cells (sole output of cerebellar cortex), Purkinje dendrites, interneurons, and glial cells (e.g., Bergman glia). Cerebellum is also rich in white matter, which contains myelinated axon bundles, and glial cells (e.g., oligodendrocytes). Note that, for FBXO45, RNA-based expert annotation could not be performed by the Human Protein Atlas due to inconclusive results (query date: 10.02.2021); hence the protein expression profile provided for this gene relies on immunohistochemistry findings only. The data visualized in this figure can be accessed via http://www.proteinatlas.org.

Uhlen M, Oksvold P, Fagerberg L, et al. (2010): Towards a knowledge-based human protein atlas. *Nature Biotechnology*. *28*(12):1248-1250.
Uhlen M, Fagerberg L, Hallström BM, et al. (2015): Tissue-based map of the human proteome. *Science*. 347(6220).

### Supplemental methods

#### *Extended methods for processing and quality control of structural MRI data*

FreeSurfer software (http://surfer.nmr.mgh.harvard.edu/) was used for automated segmentation of all structural MR images, based on probabilistic information estimated from a manually labeled training set (Fischl et al., 2002). This approach has been shown to yield well-defined cerebellar boundaries comparable in accuracy to manual labeling (Fischl et al., 2002; Lee et al., 2015). Prior findings also indicate that the FreeSurfer algorithm is especially suitable for multi-center data acquired on different scanners, as it demonstrates relatively low sensitivity to noise and variable image quality (Mayer et al., 2016; Dewey et al., 2010). Note that a mock scanner training protocol was used in the 3q29Del project to minimize motion artifacts and attrition; participants with a contraindication for MRI were excluded.

Since it is currently unfeasible to accurately reconstruct and segment the cortical surface of the cerebellum using this harmonized framework, we treated the cerebellum as a volumetric structure. For quality control (QC), two trained evaluators (ES, LL) inspected all cerebellar segmentations obtained from 3q29Del participants in axial, sagittal, and coronal reformats to determine whether technical problems (e.g., motion artifacts) or notable pathology (e.g., arachnoid cysts) interfered with registration or segmentation quality. One 3q29Del participant failed to pass QC due to a motion artifact and skull deformity interfering with the extraction of reliable volumetric measures (Fig. S1O). Similarly, the outputs of the structural pipeline for control participants were inspected by the HCP, as described by Marcus et al. (2013) and Elam et al. (2021); no major errors were identified. Detailed QC information for the HCP dataset is available at https://wiki.humanconnectome.org/. To facilitate joint analysis, the two trained evaluators additionally cross-compared the image quality, tissue contrast and cerebellar segmentation masks of 23 age- and sex-matched case-control pairs randomly selected from the entire dataset (see Fig. S1A-N for representative images). No systematic irregularities were identified between the two diagnostic groups and no manual intervention was performed in either group to avoid adding subjectivity to volumetric measures.

Since visual QC of eICV segmentation masks is not attainable in the atlas-based head size normalization approach used in the present study, we assessed the quality of our eICV measures by testing the correlations between eICV, total brain volume and head circumference among 3q29Del participants using Pearson product moment analysis. As expected from previous literature (McKinney et al., 2017; Koyabu et al., 2014; Buckner et al., 2004; Sanfilipo et al., 2004; Kollias et al., 1993), these correlations were positive and significant with moderate to strong effect sizes ($p$'s $\leq 0.05$) (Fig. S2), providing an indirect means of quality assurance for the eICV data.

#### *Extended methods for radiological evaluation of structural MRI data*

T1- and T2-weighted images were reviewed qualitatively by a board-certified neuroradiologist (AEGY) in axial, sagittal, and coronal reformats using Horos (https://horosproject.org). The conditions that were evaluated for differential diagnosis of enlarged retrocerebellar cerebrospinal fluid (CSF) space includes Dandy-Walker malformation (DWM), Blake's pouch cyst, mega cisterna magna (MCM), posterior fossa arachnoid cyst (PFAC), and isolated inferior cerebellar vermian hypoplasia. DWM, the most common posterior fossa malformation in the general population, can occur in isolation or as part of chromosomal anomalies or Mendelian disorders (Doherty et al., 2013). Diagnostic features of DWM on neuroimaging include hypoplasia of the cerebellar vermis, which is elevated and upwardly rotated, and dilation of the fourth ventricle, which fills and enlarges the posterior fossa (Bosemani et al., 2015). DWM is often associated with additional malformations, including callosal dysgenesis, occipital encephaloceles, polymicrogyria, and grey matter heterotropia (Parisi et al., 2003). Hydrocephalus may be present. Blake's pouch cyst occurs sporadically due to a lack of fenestration of the Blake pouch, a normal developmental structure, resulting in absence of communication between the fourth ventricle and the subarachnoid space, and leading to hydrocephalus (Tortori-Donati et al., 1996; Cornips et al., 2010). The cerebellum has a normal size and shape. Imaging demonstrates an enlarged fourth ventricle that communicates with an infravermian cyst, often resulting in hydrocephalus. MCM is an enlarged cisterna magna (≥10 mm on midsagittal images) with an intact vermis and a normal fourth ventricle. The posterior fossa may be enlarged, with scalloping of the occipital bone, without hydrocephalus. MCM may be caused by delayed fenestration of the Blake's pouch, whereas the absence of fenestration leads to a Blake's pouch cyst (Nelson et al., 2004). Arachnoid cysts result from duplication of the arachnoid membrane, with approximately 10% of arachnoid cysts in children occurring in the posterior fossa (Ali et al., 2014). Although PFACs may be asymptomatic and identified incidentally, they may present with macrocephaly, increased intracranial pressure, and developmental delay, particularly if CSF flow is obstructed (Marin-Sanabria et al., 2007). PFACs are isointense relative to CSF, well defined, can also result in scalloping of the occipital bone, and can exert mass effect on the cerebellum, with a normal appearance of the fourth ventricle and vermis. Isolated inferior vermian hypoplasia is characterized by partial absence of the inferior portion of the cerebellar vermis. More than 75% of patients with isolated inferior vermian hypoplasia have a favorable outcome, although in some patients, mild functional deficits in fine motor activity and receptive language may be present (Limperopoulos et al., 2006; Tarui et al., 2014).

***Details on standardized behavioral measures***

Participants were administered the *Beery-Buktenica Developmental Test of Visual-Motor Integration* (VMI, 6th edition) to assess the extent to which they can integrate their visual and fine motor abilities in a geometric design-copying task. To measure VMI, participants were asked to copy a set of geometric forms that increase in difficulty; the drawings were scored based on standard criteria outlined in the test manual according to how accurately each design was copied compared with the original.

The two supplemental tests included in the Beery VMI were additionally administered to assess visual and motor skills, separately. The visual perception supplemental test was used to assess visual analysis skills in a format requiring minimal motor input. In this task, participants were shown a series of reference geometric designs with multiple similar shapes provided below them; the task required participants to choose/match the shape that was identical to the reference design. The motor coordination supplemental test was used to assess fine motor coordination skills in a format requiring minimal visual analysis skills. In this task, participants were asked to trace a set of geometric forms with a pencil without going outside a double-lined path. Raw scores were converted to age-appropriate standard scores based on the standardization sample reported for the corresponding tests (normative mean = 100, SD = 15). Higher scores indicate better performance.

Two measures of intellectual functioning were utilized in the present study given the age range of the participants. The general conceptual ability (GCA) and full-scale intelligence quotient (FSIQ) scores from the *Differential Ability Scales* (DAS, 2nd edition) and the *Wechsler Abbreviated Scale of Intelligence* (WASI, 2nd edition) were used as composite IQ scores. The verbal reasoning and verbal comprehension domains of the two scales were used to index verbal IQ, while the non-verbal reasoning and perceptual reasoning domains indexed non-verbal IQ. The contents of these tests are sufficiently similar for combination across scales in cross-sectional analyses. Raw scores were transformed into age-appropriate standard scores based on the standardization sample reported for the corresponding tests (normative mean = 100, SD = 15). Higher scores indicate better performance.

***Extended statistical methods for penalized cubic spline and quantile spline models***

The following analyses were performed to 1) build developmental trajectories for our volumetric measures of interest (VOI) and 2) to estimate normative percentile curves for our VOIs (akin to a growth chart) as an alternative to the polynomial linear regression models presented in the main manuscript for case-control comparisons. In these supplemental analyses, penalized cubic splines were fitted to volumetric data using the *mgcv* package in *R* (version 1.8-33) (Wood, 2011; Wood, 2017) and quantile splines were fitted to volumetric data using the *fields* package in *R* (version 12.5) (Nychka et al., 2021). Given the local nature of splines, these methods are capable of capturing a wide range of nonlinear neurodevelopmental trends in our data and were incorporated into the present study to improve the statistical rigor of our central analyses. We provide a detailed summary of each approach below.

1) <u>Building developmental trajectories for VOIs:</u>

As a supplemental method, the penalized spline approach (Wahba, 1980; Eilers et al., 1996) was adopted to model volumetric changes in our brain regions of interest across age, since this method offers increased mathematical flexibility compared with parametric models that rely on delicate assumptions. As in regression splines (Eilers et al., 1996), penalized splines make use of piecewise polynomial approximation with a flexible selection of knots to effectively capture the underlying changes in a dataset, when given a fixed basis-dimension and appropriate positioning of the knots that provides fair coverage of the covariate values (Wood, 2017). This creates computational efficiency compared with smoothing splines, which place a knot at every unique sample point. Moreover, penalized splines add a roughness (or "wiggliness") penalty to the model, as in smoothing splines (Reinsch, 1967), to control the smoothness of the fit while avoiding the problem of overfitting. In summary, penalized splines offer an effective compromise between regression splines and smoothing splines (two popular tools) for more closely approximating age-associated changes in our outcome variables of interest.

Below, we will gradually build our way towards defining the penalized cubic spline approach that was used to characterize the developmental trajectories of our VOIs.

The definition of a natural spline is as follows (given a set of knots at $k_1, k_2, \ldots, k_K$):

1. $f(x)$ is a polynomial of degree $p$ on each of the intervals $[k_1, k_2]$, $[k_2, k_3]$, ..., $[k_{K-1}, k_K]$.
2. $f(x)$ has continuous $(p-1)_{th}$ derivatives at knots $k_1, k_2, \ldots, k_K$.
3. $f(x)$is a polynomial of degree $\frac{(p-1)}{2}$ on $(-\infty, k_1]$ and $[k_K, \infty)$.

The last requirement forces a lower degree to the left (or right) of the leftmost (or rightmost) knot, to reduce the variance at the boundaries of the observations. In the present study, we used natural cubic splines $(p = 3)$, which are linear beyond the boundary knots.

The unique solution to the optimization problem described in *equation (1)* is a natural cubic spline with knots at every unique observation $x_1, x_2, \ldots, x_n$. The solution $f(x)$ is called a smoothing spline (Reinsch, 1967).

$$\min_f \{\frac{1}{n}\sum_{i=1}^{n}(y_i - f(x_i))^2 + \lambda \int (f''(x))^2 dx\} \qquad \text{Equation (1)}$$

where $(x_i, y_i), i = 1, \ldots, n$ are a set of observations, $f(x)$ includes all that have continuous second derivatives and $\lambda > 0$ is a smoothing parameter that controls the trade-off between fidelity to the data and roughness of the function estimate.

Although smoothing splines enjoy the theoretical property of an optimizing solution (as described above), the degrees of freedom (i.e., number of knots) are comparable to the data to be smoothed and can be redundant, as the knots are placed at every unique observation. If the penalty term in *equation (1)* is removed and $f(x)$ is restricted to a natural spline with user-defined knots (as in the definition of a natural spline where the knot positions at $k_1, k_2, \ldots, k_K$ can be completely decided by users), the solution is reduced to a regression spline. Although regression splines ease the computational redundancy of smoothing splines by removing any restrictions in knot selection, they have the potential to be over-fitted. This can be handled by a regularization term, such as the integrated square second derivative in smoothing splines. Therefore, it is natural to combine the freely chosen degrees of freedom of a regression spline and the roughness penalty of a smoothing spline, which in turn gives penalized splines (Wahba, 1980). Specifically, we use penalized cubic splines in the present study, which is the solution to the optimization problem in *equation (2)*, where the number and positions of the knots are freely chosen in advance and the number of knots is usually much smaller than the sample size:

$$\begin{cases} \min_f \{\frac{1}{n}\sum_{i=1}^{n}(y_i - f(x_i))^2 + \lambda \int (f''(x))^2 dx\} \\ f \text{ is a natural cubic spline with predefined knots at } k_1, \dots, k_K \end{cases} \quad \text{Equation (2)}$$

To obtain a satisfying fit, the knots should be arranged nicely to cover the distribution of the covariate in the original data set (Wood, 2017).

For each VOI (i.e., total cerebellum volume, cerebellar cortex volume, cerebellar white matter volume, estimated total intracranial volume (eICV), and the eICV-adjusted versions of cerebellar volumes), we denote $y_i$ as the corresponding volume of the $i_{th}$ participant in the pooled dataset (i.e., data aggregated across the two diagnostic groups). It is assumed that $y_i$ is represented by a function of the participant's age, with sex and diagnostic group added as covariates, as expressed in *equation (3).*

$$y_i = f(Age_i) + \alpha \cdot Sex_i + \beta \cdot Group_i + \epsilon_i,\ \epsilon_i \overset{\text{iid}}{\sim} N(0, \sigma^2) \quad \text{Equation (3)}$$

$Sex_i$ is a sex indicator taking 1 for males and 0 for females. $Group_i$ is a diagnostic group indicator taking 1 for 3q29Del participants and 0 for healthy control participants. The residual random error $\epsilon_i$ is assumed to follow a Gaussian distribution with mean 0 and a common variance $\sigma^2$ across participants (homoscedasticity). $f(Age_i)$ is a natural cubic spline term of age. The number of knots is chosen to be 10 and they evenly cover the quantiles of the age distribution (i.e., the knots are placed at the 0%, 11.11%, 22.22%, 33.33%, 44.44%, 55.56%, 66.67%, 77.78%, 88.89%, and 100% quantiles of age in the pooled data).

Following the optimization framework of a penalized cubic spline described in *equation (2)*, the parameter estimates are obtained via the objective function:

$$\begin{cases} \min_f \{\frac{1}{n}\sum_{i=1}^{n}(y_i - f(Age_i) - \alpha \cdot Sex_i - \beta \cdot Group_i)^2 + \lambda \int (f''(x))^2 dx\} \\ f \text{ is a natural cubic spline with with 10 knots spread evenly across the age quantiles} \end{cases} \quad \text{Equation (4)}$$

where the smoothing parameter $\lambda$ is selected by the restricted maximum likelihood (REML) method (Wood, 2011).

From the fitted penalized cubic splines, mean developmental trajectories for each VOI were estimated and laid onto the scatter plots of the original volumetric data points in Fig. S7. The effective degrees of freedom (EDF) reported in Table S4 represent how complex the neurodevelopmental pattern of the estimated volumetric trajectory of each VOI is across age. EDF = 1 is equivalent to a straight line, EDF = 2 is equivalent to a quadratic curve, etc., with higher EDFs describing increased wiggliness (Wood, 2017). The EDF of the smooth term is defined as the trace of the influence matrix. The significance of the smooth term $f(Age)$, the sex indicator and the diagnostic group indicator are also reported in Table S4 in the form of p-values. Note that the significance of the smooth term is derived from an approximate F-test of whether the smooth term of age is significant in the penalized cubic spline models for each VOI ($f(Age) = 0$). The p-values are approximate in the sense that the components of the test statistic are weighted by the iterative fitting weights (Wood, 2017). A comprehensive description of how to obtain theoretical p-values is detailed by Wood (2013). The p-values for sex and diagnostic group are derived from standard t-tests of whether the indicator of sex/diagnostic group is significant in determining each VOI.

All of the aforementioned parameter estimates and p-values were obtained using the *gam()* function from the *mgcv* package in R (Wood, 2011; Wood, 2017). Using the function *gam.check()* from the same package, model diagnostics were performed, which confirmed that the required assumptions of the penalized cubic spline model in *equation (3)* were met and our basis dimension (10 knots of 9 basis dimension) was adequate to cover the age distribution of the present study sample. More specifically, QQ-plots were explored to justify the Gaussian assumption of the residuals; residuals versus linear predictors/fitted values were plotted to justify the homoscedasticity assumption; the histogram of residuals was plotted to check the normality of the residuals; and the p-value for the residual randomization test was explored to check the adequacy of the basis dimension.

2) *Estimating normative percentile curves for VOIs:*

Besides estimating mean volumetric changes across age in the entire study sample (i.e., data pooled across the two diagnostic groups), another interest was in estimating sex-specific normative percentiles for volumetric changes observed across age in our healthy control participants only. Here, our goal was to establish well-founded references (or "nomograms") for comparison against the VOIs of individual 3q29Del participants to further characterize the degrees of neuroanatomical deviance observed within the 3q29Del sample. A secondary goal was to explore the relative distribution of 3q29Del data points in these normative charts to gain insights into potential age-windows of heightened structural vulnerability. However, the limited sample size of the 3q29Del group (especially when stratified by sex), and the paucity of data on control participants younger than age 5 precluded our ability to reach any meaningful inferences in this regard. We foresee that these estimates may become particularly useful for future studies investigating longitudinal neuroimaging outcomes in 3q29Del participants.

In the supplemental analyses described below, we characterized the 10$^{th}$, 25$^{th}$, 50$^{th}$, 75$^{th}$, and 90$^{th}$ percentiles of normative volumetric growth curves for each VOI, stratified by sex. Similar to modelling mean volume changes, our goal here was to model normative percentiles under a penalized spline framework, hence we used the quantile smoothing splines approach for this objective.

In quantile regression, the check loss shown in *equation (5)* is used as the objective of model fitting, where $\alpha$ represents the $\alpha$ quantile of the dependent variable $y$, $x$ is the independent variable influencing $y$, and $1_{\{u<0\}}$ is an indicator taking 1 when $u < 0$ and 0 otherwise.

$$\rho_\alpha\big(y - g(x)\big) = (y - g(x))(\alpha - 1_{\{(y-g(x))<0\}}) \qquad \text{Equation (5)}$$

To improve computation, Nychka et al. (1995) proposed a modified check loss which rounds out the corner of the check loss in a small interval around zero by piecing in a quadratic function, in order to make the loss function differentiable at zero. The modified check loss is shown in the following equation:

$$\rho_\alpha, C\big(y - g(x)\big) = \begin{cases} \rho_\alpha\big(y - g(x)\big), & if\ |y - g(x)| > C \\ \dfrac{1-\alpha}{C}(y - g(x))^2, & if\ 0 \le y - g(x) \le C \\ \dfrac{\alpha}{C}\big(y - g(x)\big)^2, & if\ -C \le y - g(x) < 0 \end{cases} \qquad \text{Equation (6)}$$

where $C$ is a scale factor for rounding out the absolute value function at zero to a quadratic (Nychka et al., 2021). According to Nychka et al. (1995), $C$ should be chosen to be effectively zero relative to the magnitude of the data values.

After presenting the form of the modified check loss, we can adopt the optimization framework of smoothing splines shown in *equation (1)* by simply replacing the squared error loss to the modified check loss in *equation (6)*:

$$\min_{g}\{\frac{1}{n}\sum_{i=1}^{n}\rho_{\alpha},C(y_i - g(x_i)) + \lambda\int(g''(x))^2dx\} \qquad \text{Equation (7)}$$

Similar to least square smoothing splines, the quantile smoothing splines that we adopt use the L2 roughness penalty on the integral of the squared second derivative.

For each VOI (i.e., total cerebellum volume, cerebellar cortex volume, cerebellar white matter volume, estimated total intracranial volume (eICV), we denote $y_i$ as the corresponding volume of the $i_{th}$ healthy control participant, stratified by sex. It is assumed that the $\alpha$ quantile of $y_i$ is represented by a function of the participant's age $g(Age_i)$ (i.e., $P(y_i \leq g(Age_i) = \alpha)$. Following the optimization framework of a quantile spline described in *equation (7)*, the parameter estimates are obtained via the following objective over all $g$, such that the roughness penalty is finite:

$$\min_{g}\{\frac{1}{n}\sum_{i=1}^{n}\rho_{\alpha},C(y_i - g(Age_i)) + \lambda\int(g''(x))^2dx\} \qquad \text{Equation (8)}$$

where the smoothing parameter $\lambda$ is selected by the generalized cross-validation (GCV) method (Graven, 1989), and $C$ is set to $10^{-5}$ of the variance of the $y$'s.

From the fitted quantile splines for $\alpha$ = 0.10, 0.25, 0.50, 0.75, 0.90, normative developmental trajectories for the $10^{th}$, $25^{th}$, $50^{th}$, $75^{th}$, and $90^{th}$ percentiles of the VOIs were estimated, respectively. The computation was performed using the *qsreg()* function from the *fields* package in R (Douglas Nychka et al., 2021). In Fig. S9, the original VOIs of individual 3q29Del participants are plotted against the normative percentile curves derived from male and female controls, separately. This case-by-case comparison provides a general idea of which age- and sex-specific normative percentile each 3q29Del participant's cerebellar and eICV volumes correspond to.

***List of R packages used for statistical analyses and diagnostics***

Multiple linear regression analyses were performed via the standard R *lm()* function. For model comparisons, analyses of variance (ANOVA) were performed by the standard R *anova()* function. Wald statistics were calculated by the *waldtest()* function from the *lmtest* package (https://CRAN.R-project.org/package=lmtest). Heteroscedasticity-robust estimates were calculated using the *vcovHC()* function from the *sandwich* package (https://CRAN.R-project.org/package=sandwich). Permutation tests were performed using the *lmperm()* function of the *permuco* package https://CRAN.R-project.org/packages=permuco). Diagnostics plots for linear regression were created using the R base function *plot()*. Shapiro-Wilk tests were performed using the standard R *shapiro.test()* function to check for assumptions of normality. Breusch-Pagan and Levene's test were performed using the *bptest()* function from the *lmtest* package (https://CRAN.R-project.org/package=lmtest) and the *levene_test()* function from the *rstatix* package (https://CRAN.R-project.org/package=rstatix) to check for homogeneity of variances, respectively. Fisher's exact tests, Pearson's chi-squared tests, Student's two sample t-tests and Wilcoxon signed-rank tests were performed using the standard R *fisher.test(), chisq.test(), t.test()* and *wilcox.test()* functions. Effect sizes for Wilcoxon signed-rank tests were calculated using the *wilcox_effsize()* function from the *rstatix* package (https://CRAN.R-project.org/package=rstatix). The standard R *cor.test()* function was used to perform Pearson's correlations. The *pcor.test()* function from the *ppcor* package (https://CRAN.R-project.org/package=ppcor) was used to calculate partial correlations. For spline modeling, the *gam()* and *gam.check()* functions from the *mgcv* package (https://cran.r-project.org/package=mgcv) and the *qsreg()* function from the *fields* package (https://cran.r-project.org/package=fields) were used. Graphics were generated by the *ggplot2* (https://CRAN.R-project.org/package=ggplot2), *ggpubr (https://cran.r-project.org/package=ggpubr)* and *jtools* (https://cran.r-project.org/package=jtools) packages.

***References for supplemental methods***

Ali ZS, Lang SS, Bakar D, Storm PB, Stein SC (2014): Pediatric intracranial arachnoid cysts: comparative effectiveness of surgical treatment options. *Childs Nerv Syst*. 30:461-469.

Bosemani T, Orman G, Boltshauser E, Tekes A, Huisman TA, Poretti A (2015): Congenital abnormalities of the posterior fossa. *Radiographics*. 35:200-220.

Buckner RL, Head D, Parker J, Fotenos AF, Marcus D, Morris JC, et al. (2004): A unified approach for morphometric and functional data analysis in young, old, and demented adults using automated atlas-based head size normalization: reliability and validation against manual measurement of total intracranial volume. *Neuroimage*. 23:724-738.

Cornips EM, Overvliet GM, Weber JW, Postma AA, Hoeberigs CM, Baldewijns MM, et al. (2010): The clinical spectrum of Blake's pouch cyst: report of six illustrative cases. *Childs Nerv Syst*. 26:1057-1064.

Dewey J, Hana G, Russell T, Price J, McCaffrey D, Harezlak J, et al. (2010): Reliability and validity of MRI-based automated volumetry software relative to auto-assisted manual measurement of subcortical structures in HIV-infected patients from a multisite study. *Neuroimage*. 51:1334-1344.

Doherty D, Millen KJ, Barkovich AJ (2013): Midbrain and hindbrain malformations: advances in clinical diagnosis, imaging, and genetics. *Lancet Neurol*. 12:381-393.

Eilers PH, Marx BD (1996): Flexible smoothing with B-splines and penalties. *Statistical science*. 11(2):89-121.

Elam JS, Glasser MF, Harms MP, Sotiropoulos SN, Andersson JLR, Burgess GC, et al. (2021): The Human Connectome Project: A retrospective. *Neuroimage*. 244:118543.

Fischl B, Salat DH, Busa E, Albert M, Dieterich M, Haselgrove C, et al. (2002): Whole brain segmentation: automated labeling of neuroanatomical structures in the human brain. *Neuron*. 33:341-355.

Graven P (1989): Smoothing noisy data with spline function: estimating the correct degree of smoothing by the method of generalized cross-validatIon. *Number. Math.* 31:377-403.

Kollias SS, Ball WS, Jr., Prenger EC (1993): Cystic malformations of the posterior fossa: differential diagnosis clarified through embryologic analysis. *Radiographics*. 13:1211-1231.

Koyabu D, Werneburg I, Morimoto N, Zollikofer CP, Forasiepi AM, Endo H, et al. (2014): Mammalian skull heterochrony reveals modular evolution and a link between cranial development and brain size. *Nature communications*. 5:1-9.

Lee DK, Yoon U, Kwak K, Lee JM (2015): Automated Segmentation of Cerebellum Using Brain Mask and Partial Volume Estimation Map. *Comput Math Methods Med*. 2015:167489.

Limperopoulos C, Robertson RL, Estroff JA, Barnewolt C, Levine D, Bassan H, et al. (2006): Diagnosis of inferior vermian hypoplasia by fetal magnetic resonance imaging: potential pitfalls and neurodevelopmental outcome. *Am J Obstet Gynecol*. 194:1070-1076.

Marcus DS, Harms MP, Snyder AZ, Jenkinson M, Wilson JA, Glasser MF, et al. (2013): Human Connectome Project informatics: quality control, database services, and data visualization. *Neuroimage*. 80:202-219.

Marin-Sanabria EA, Yamamoto H, Nagashima T, Kohmura E (2007): Evaluation of the management of arachnoid cyst of the posterior fossa in pediatric population: experience over 27 years. *Childs Nerv Syst*. 23:535-542.

Mayer KN, Latal B, Knirsch W, Scheer I, von Rhein M, Reich B, et al. (2016): Comparison of automated brain volumetry methods with stereology in children aged 2 to 3 years. *Neuroradiology*. 58:901-910.

McKinney AM (2017): Mega Cisterna Magna and Retrocerebellar Arachnoid Cysts. *Atlas of Normal Imaging Variations of the Brain, Skull, and Craniocervical Vasculature*: Springer, pp 19-41.

Nelson MD, Jr., Maher K, Gilles FH (2004): A different approach to cysts of the posterior fossa. *Pediatr Radiol*. 34:720-732.

Nychka D, Gray G, Haaland P, Martin D, O'connell M (1995): A nonparametric regression approach to syringe grading for quality improvement. *Journal of the American Statistical Association*. 90(432):1171–1178.

Nychka D, Furrer R, Paige J, Sain S (2021): "fields: Tools for spatial data". R package version 12.5, https://github.com/dnychka/fieldsRPackage

Parisi MA, Dobyns WB (2003): Human malformations of the midbrain and hindbrain: review and proposed classification scheme. *Mol Genet Metab*. 80:36-53.

Reinsch CH (1967): Smoothing by spline functions. *Numerische mathematik*. 10(3):177-183.

R Development Core Team (2021): R: A Language and Environment for Statistical Computing. Vienna, Austria: R Foundation for Statistical Computing.

Sanfilipo MP, Benedict RH, Zivadinov R, Bakshi R (2004): Correction for intracranial volume in analysis of whole brain atrophy in multiple sclerosis: the proportion vs. residual method. *Neuroimage*. 22:1732-1743.

Tarui T, Limperopoulos C, Sullivan NR, Robertson RL, du Plessis AJ (2014): Long-term developmental outcome of children with a fetal diagnosis of isolated inferior vermian hypoplasia. *Arch Dis Child Fetal Neonatal Ed*. 99:F54-58.

Tortori-Donati P, Fondelli MP, Rossi A, Carini S (1996): Cystic malformations of the posterior cranial fossa originating from a defect of the posterior membranous area. Mega cisterna magna and persisting Blake's pouch: two separate entities. *Childs Nerv Syst*. 12:303-308.

Wahba G (1980): Spline bases, regularization, and generalized cross validation for solving approximation problems with large quantities of noisy data. University of WISCONSIN.

Wood SN (2011): Fast stable restricted maximum likelihood and marginal likelihood estimation of semiparametric generalized linear models. *Journal of the Royal Statistical Society: Series B (Statistical Methodology).* 73(1):3-36.

Wood SN (2013): On p-values for smooth components of an extended generalized additive model. *Biometrika*. 100(1), 221-228.

Wood SN (2017): Generalized additive models: an introduction with R. CRC press.
